# Revolutionizing Lung Cancer Detection: Evaluating AI Models for VOC Analysis and Unveiling Key Exhaled Biomarkers

**DOI:** 10.1101/2025.02.03.25321431

**Authors:** Francesco Hamilton

## Abstract

Volatile Organic Compounds (VOCs) are organic chemicals that readily vaporize at room temperature and are emitted from diverse sources, including paints, building materials, and biological processes^1^. While the understanding of VOC production within the human body remains limited, it is well-established that VOCs arising from cellular metabolic activities accumulate in significant concentrations in the bloodstream^2^. As cellular processes are disrupted by bacterial or viral infections or the onset of cancer, these metabolic pathways undergo substantial alterations, leading to distinct changes in the VOC profiles generated by lymphocytes and other cell types. These alterations manifest in the VOC composition present in the blood and exhaled breath^3^. This study seeks to advance the field by identifying optimal AI-based analytical methods for lung cancer diagnosis, assessing the accuracy of non-invasive VOC-based diagnostic approaches, and characterizing the key VOCs influenced by the metabolic shifts associated with cancer cells.

This analysis was conducted using a dataset comprising 427 anonymized cases, including patients with benign lung nodules, malignant lung cancers, and healthy controls. Using this dataset we trained a neural network, support vector machine (SVM), Convolutional Neural Network (CNN), Recurrent Neural Network (RNN), and XGBoost model, allowing for the accuracy of predictions provided by each model to be compared. To generate multiple comparisons and metrics of evaluation, the dataset was split into single “case” comparisons comparing patients without tumors and those with benign/malignant tumors individually, and one case comparing all three classes together. This, combined with train-test splits of 70/30 (where 70% of the dataset is randomly used for training the model and 30% to evaluate the model’s effectiveness), 80/20, and 90/10, allows for a thorough evaluation of the ROC score, precision, and F1 score of each model across multiple conditions.

In addition to evaluating the viability of VOC-based diagnosis using artificial intelligence models, this study aimed to identify the key VOCs in lung cancer that contribute to a successful diagnosis. To determine important VOCs for diagnosis, the dataset was analyzed using a Random Forest Model, a multiple decision tree-based model, and an XGBoost model (using SHAP values), independently ranking and outputting the 5 most important VOCs used in distinguishing lung cancer, benign lung nodules, and healthy patients. Using the Random Forest (RF) model, we identified the VOCs most critical for distinguishing among all three classes—benign lung nodules, malignant lung cancers, and controls. The top VOCs included C13H22O, C4H8O2, C4H8O, C7H11O, and C5H10O, followed by C2H4O2, C6H10O2, and C6H12O. When differentiating between cancer and control classes, the most important VOCs were C4H8O, C4H8O2, C13H22O, C11H22O, and C7H6O. For distinguishing benign and control classes, the key VOCs identified were C13H22O, C7H11O, C4H8O2, C6H10O2, and C2H4O2.

When comparing VOC profiles between patients with benign and cancerous nodules and in comparison of those with benign nodules against control patients, XGBoost was the most accurate and effective for successful diagnosis. However, when used for distinguishing cancerous nodules and control patients, neural networks outperformed the XGBoost models with a weighted F1 score of 0.96 and an ROC score of 0.96 compared to the F1 score of 0.94 and ROC score of 0.94 in the XGBoost model. CNNs and RNNs also provided ample performance (lower than their decision tree and hyperplane-based counterparts) but each with its strengths. CNNs provided higher “top end” performance with accuracies across control and cancer classes being disproportionately higher, while RNNs had a higher average with all three classes having similar accuracies (i.e. performed better across benign classes). In their best comparisons (using case 4 and a 75/25 split) the CNN had an overall accuracy of 0.77 while the RNN had an accuracy of 0.79 (using a multiclass dataset and an 80/20 split).

The VOCs identified in this study provide a foundation for future research aiming to accurately diagnose lung cancer using AI-analysis methods and elucidate the metabolic changes that enable effective classification (Table 1.3). The machine learning methods evaluated in this paper each provide their strengths, and this study attempts to quantify the benefits of each of these methods through various comparisons mirroring what these models would have to analyze in a screening environment. By highlighting these advantages, this research aims to guide future efforts in synthesizing models to further enhance the accuracy and reliability of VOC-based lung cancer diagnostic methods.

## Introduction

With one in sixteen people affected by lung cancer in their lifetime and accounting for one in five cancer deaths, lung cancer is the leading cause of cancer death^4^. Despite the availability of lung cancer screenings, the average diagnosis for early-stage lung cancer occurs 141 days after symptoms first emerge^5^, with 47% of diagnoses being late-stage^4^. While screening and surveys have had widespread success, they have been limited by the risks of overdiagnosis and false positives in patients with low-risk cancers^6^.

Breathomics, the analysis of exhaled breath metabolites, seeks to shorten this time frame by taking advantage of the strong correlation of certain exhaled compounds with Non-Small Cell Lung Cancers (NSCLC). Specifically, they target Volatile Organic Compounds (VOCs), organic compounds naturally released into the bloodstream by human cells as a byproduct of their metabolic processes^2^. Researchers can diagnose cancers using VOCs due to cancer’s erratic upregulation of metabolic pathways/cellular processes and the subsequent change in VOC “profiles”. AI models can sift through this data, taking note of these key differences, and generate a diagnosis using the concentrations of key aldehydes and ethanol derivatives in the patient’s blood and breath samples^7^.

Despite the promise of Breathomics for lung cancer diagnosis, standardized methods of non-invasive exhaled VOC analysis have been hindered by high cost, sample variability, and the lack of standardization in statistical analysis methods. Important VOCs in cancer diagnosis have varied between studies due to the high variability in breath sampling procedures, different analysis methods, and environmental conditions. In addition, the common sample collection method: gas chromatography combined with mass spectroscopy, proves costly and difficult to perform analyses, as it requires specially trained technicians and facilities to allow for interpretation. This study seeks to lay the groundwork for further implementation of AI-guided VOC analysis by establishing the best artificial intelligence algorithms for generating reliable diagnoses, testing potential algorithms for future VOC analysis using complex multi-institutional datasets, and evaluating the individual strengths of the most common models used in VOC analysis. In addition, it seeks to identify the most important VOCs and associated pathways, allowing for diagnosis of benign growths and lung tumors alike to facilitate future research evaluating breathomics as a viable NSCLC screening method.

To perform these analyses, this study utilized the molecular concentration data of the VOCs from 427 anonymized patients generated by the Dr. Fu lab at the University of Louisville^10^. The dataset includes benign, cancerous, and control cases and the corresponding molecular concentrations for all major VOCs associated with lung cancer as indicated by past studies. To compare these cases, 4 datasets were created with 3 One-vs-One cases: Case 1, which contained 156 cancer patients compared against the 193 control patients, Case 2 comparing the cancer patients and the 65 benign patients, Case 3 using benign and control patients, and 1

One-vs-Rest (multiclass comparison) case named Case 4. Case 4 incorporated all patient classes with each class being compared against the rest of the classes individually and compiled to a final result(e.g. benign and cancer classes being compared against the control patients). These values were then implemented into an SVM, XGBoost, and Neural Network, and each model was evaluated based on its performance^13^ ^14^ ^15^. To explain which VOCs were crucial in identifying lung cancer, this study also utilized a random forest model, an ensemble machine learning model based on decision trees, to discover important “features” (VOCs) in each case analysis and cross-referenced that with the SHAP (PPV)^17^ values for each compound generated using an XGBoost model.

Convolutional Neural Networks, typically used for spatial pattern analysis problems, are a subset of neural networks commonly integrated with other model types to provide more accurate analysis of intricate trends. To examine the viability of CNNs for VOC analysis, this study implemented a CNN and trained it on 4 different train-test splits in a multiclass comparison (70/30, 75/25, 80/20, and 90/10) to evaluate its accuracy and AUC score across cancer, benign, and control cases. To provide further analysis, a Receiver Operating Characteristic (ROC) and Precision Recall (PR) curve were generated using the sklearn metrics tool. In addition to a CNN, this study also examined the viability of Recurrent Neural Networks (denoted RNN) using similar splits and plotted ROC and PR curves to determine its performance.

## Methods

To source a dataset for model training, this study utilized the Dr. Fu lab group’s dataset of 427 cases measuring the Volatile Organic Compound Levels in patients with benign and cancerous tumors in addition to control cases^10^. The dataset comprised 193 control cases, 65 benign cases, and 157 cancer cases, further divided into single comparisons between each class and one whole group comparison (malignant, control, and benign). To confirm group homogeneity and ensure the presence of significant group-based differences, 2D PCA plot analyses in the multiclass and single comparisons were performed. Using sklearn’s PCA and StandardScaler the data was standardized, and the PCA results were generated, allowing for further analysis. The PCA plots displayed each case as a point aligned on a graph based on its VOC values, allowing for the identification of clear class separation and distinct traits between classes. To load and preprocess the dataset, VOC (Volatile Organic Compound) values and class labels were extracted from the dataset’s .csv file, with VOCs forming the feature matrix *X* and the class labels, indicating cancer status, forming the target array *y*. For preprocessing, the VOC values in *X* were scaled using StandardScaler, which standardized features by removing the mean and scaling to unit variance. The class labels were encoded as categorical variables, where each label (e.g., Cancer, Benign, Control) was transformed into one-hot encoding using LabelEncoder and then to categorical values.

The dataset was then implemented into three different deep learning analysis models: a Support Vector Machine^14^, an XGBoost model^15^, and a Forward Neural Network^13^. SVMs are supervised learning models designed to classify data by finding the optimal hyperplane that separates classes in high-dimensional space^14^. Their ability to effectively handle both linear and non-linear classification tasks makes them particularly valuable in distinguishing complex VOC signatures, especially in smaller datasets when spectral overlaps occur. XGBoost, an advanced gradient boosting algorithm, builds decision trees iteratively by minimizing prediction errors through weighted adjustments, enabling it to capture intricate feature relationships at the cost of requiring more training data^15^. This makes XGBoost especially effective in scenarios where VOC datasets exhibit high complexity or feature interdependence, such as environmental monitoring or industrial diagnostics^15^. Feedforward neural networks (denoted NNs in this paper), composed of interconnected layers of neurons, are adept at modeling non-linear relationships within input features, but also may require larger datasets^13^. They excel in complex pattern recognition tasks, such as time-series VOC data analysis or high-dimensional molecular fingerprint interpretation^13^.

The Neural Network utilized three (dense) layers and 100 epochs, with layer 1 using a ReLu activation function and 64 neurons, layer 2 containing 32 neurons, and layer 3 serving as the output layer with three outputs: control, cancer, and benign. This specific architecture was chosen because there were 400 patients in this dataset, and a smaller NN was required. The SVM utilized the radial basis function (RBF) kernel with probability estimates enabled, allowing for non-linear classification due to the highly variable nature of VOC values. The XGBoost model, a decision tree-based algorithm, used the log-loss evaluation metric to classify VOC data with fine-grained feature selection.

To evaluate the performance of Convolutional Neural Networks (CNNs) and Recurrent Neural Networks (RNNs) in analyzing Volatile Organic Compound (VOC) data, two distinct architectures were designed to handle the sequential and feature-rich nature of PPM-level measurements. The CNN was tailored to capture spatial patterns in the data, consisting of two convolutional layers with 64 and 128 filters, each using a kernel size of 3 and ReLU activation. MaxPooling layers with a pool size of 2 followed each convolution to downsample the data. The extracted features were flattened and passed through a fully connected layer with 128 neurons, activated by ReLU, before reaching the output layer, which used a softmax activation function to predict class probabilities. The RNN architecture, by contrast, focused on modeling temporal dependencies in the data through two stacked LSTM layers. The first LSTM layer had 64 units with sequences returned to facilitate stacking, while the second layer had 128 units and produced a single feature vector for classification. This was followed by a dense layer with 128 ReLU-activated neurons and an output layer similar to that in the CNN.

Both models were implemented using TensorFlow/Keras, with input data standardized using Scikit-learn’s StandardScaler and reshaped into a format suitable for sequential analysis. The architectures were trained for 1000 epochs with a batch size of 32, using Adam as the optimizer and categorical cross-entropy as the loss function. Data splits for training and validation varied across experiments, including configurations such as 70/30, 75/25, 90/10, and 80/20 splits. The CNN was designed to excel at identifying localized features within the input, while the RNN emphasized capturing sequential patterns and long-term dependencies. Performance evaluation included accuracy, precision, recall, confusion matrices, and both Receiver Operating Characteristic (ROC) and Precision-Recall (PR) curves, providing a comprehensive basis for comparing their suitability in VOC classification.

The performance of each model was then plotted to ROC (receiver operating characteristic) and precision-recall curves in sklearn.metrics to determine model performance across threshold values using a One-vs-Rest approach for case 4 and a One-vs-one approach for cases 1, 2, and 3.

These models were then trained on the dataset, and utilizing a randomized portion of the dataset allowed for testing the accuracy of each model and assessing the best model for VOC interpretation. To generate the most important VOCs for NSCLC diagnosis, this study utilized feature importances generated by a Random Forest Model in combination with SHAP (Shapley Additive Explanations) values created using an XGBoost model. Using both models allowed us to compare whether important features varied across decision tree-based models, and ensure accurate identification of key VOCS. To source the biological implications and common names of the top 5 VOCs (identified by the random forest model) we utilized the PubChem database to find genes and pathways associated with 1-Tridecanol, Butyric acid, Butyraldehyde, cyclohexylmethanone, and 2-Pentanone, which alone contributed to the majority of the decision in case 4 (multiclass) for XGBoost and Random Forest Models.

To display model performance, we utilized the sklearn classification_report and confusion_matrix functions with each model across train/test splits, which provided specific details about their accuracy and predictions.

Due to Dr. Fu’s dataset already being anonymized, no further work was needed in terms of ethical considerations, and due to the data being open access, no further permission was required. The data used in this study were obtained from the publicly available open-source dataset sourced from the Dr. Fu lab group at the University of Louisville (https://doi.org/10.1371/journal.pone.0277431). We are grateful for the availability of this comprehensive resource, which facilitated our investigation into the role of volatile organic compounds (VOCs) in lung cancer detection. The manuscript and its supplementary materials include all relevant data for this study.

## Results

In the multiclass comparison, XGBoost had the highest accuracy at 86% in the 80/20 data split using the full multiclass dataset, compared to the SVM (80%) and Neural Network (77%) models, as demonstrated in Figure 3.9. This also consisted of an F1 score of 0.85 in the XGBoost model, 0.78 in the SVM, and 0.77 in the Neural Network. Similar patterns emerged in 70/30 and 90/10 splits, with XGBoost consistently outperforming or matching the accuracy of the other methods; however, accuracy suffered due to smaller training sets, with the SVM marginally outperforming the XGBoost (0.84 compared to 0.82) in the 70/30 comparison. These results are corroborated across Figures 3.4, 3.5, 3.7, and 3.7 with XGB having a higher ROC AUC in both splits when compared to NN, with an average ROC AUC of 0.71 (70/30) and 0.87 (80/20). While SVM outperforms XGB slightly in the 80/20 split (0.89 vs. 0.87), XGB shows strong and consistent ROC performance in both splits, particularly in distinguishing the control class with an AUC of 0.98 and 0.95 when identifying cancer patients in the 80/20 split. While XGB has a lower PR AUC for the benign class in the 80/20 split (0.45), its strong performance in cancer (0.80) and control (0.98) contributes to maintaining the highest overall average AUC. XGBoost’s performance in cancer varies between Precision-Recall and ROC methods, increasing from 0.8 to 0.86 from the 80/20 split to the 70/30 split in PR, but decreasing from 0.95 to 0.69 in ROC. XGB has a higher ROC AUC in both splits than NN, with an average ROC AUC of 0.71 (70/30) and 0.87 (80/20).

**Table 1.2:**
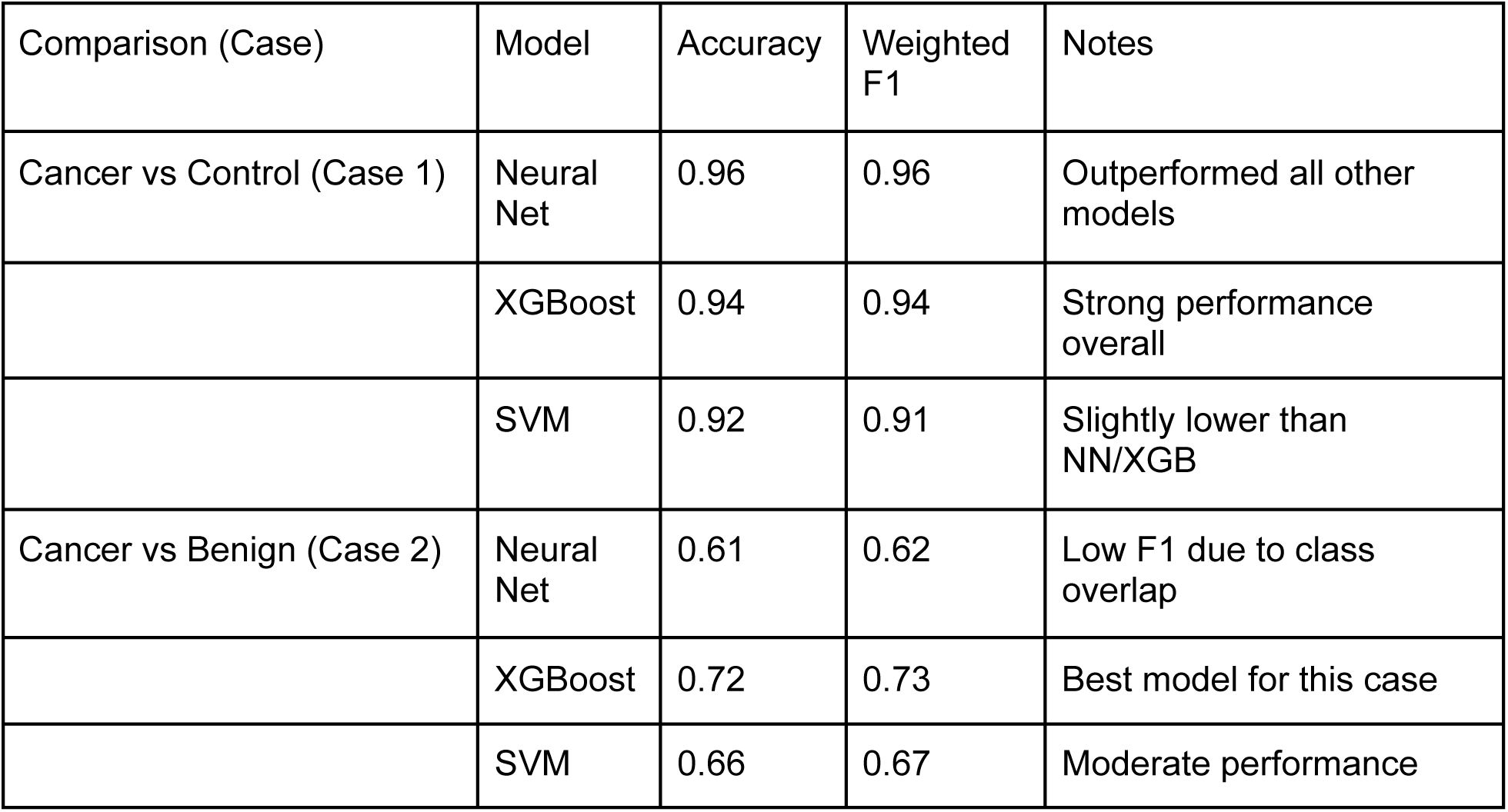
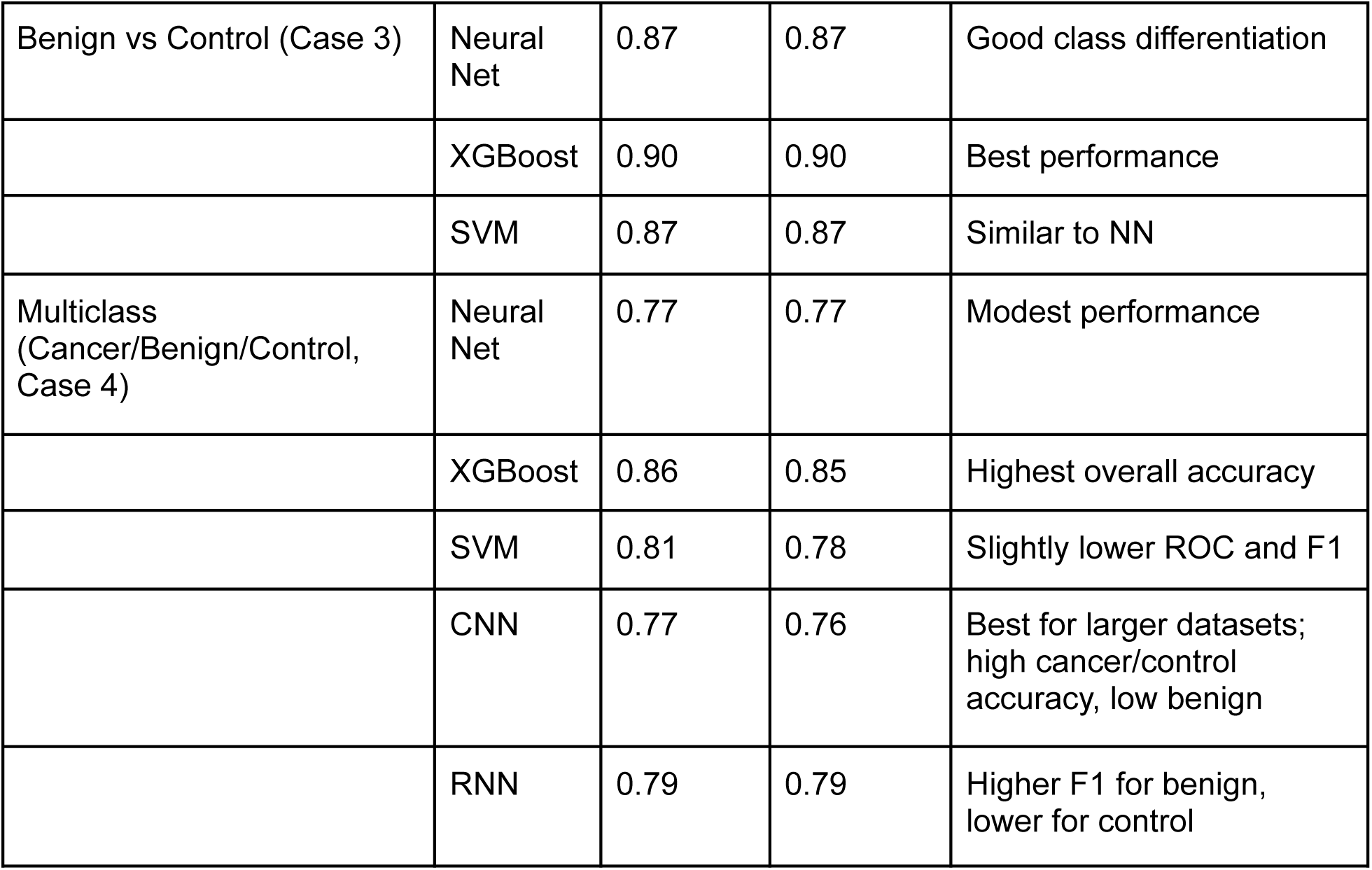

| Comparison (Case) | Model | Accuracy | Weighted F1 | Notes |
| --- | --- | --- | --- | --- |
| Cancer vs Control (Case 1) | Neural Net | 0.96 | 0.96 | Outperformed all other models |
|  | XGBoost | 0.94 | 0.94 | Strong performance overall |
|  | SVM | 0.92 | 0.91 | Slightly lower than NN/XGB |
| Cancer vs Benign (Case 2) | Neural Net | 0.61 | 0.62 | Low F1 due to class overlap |
|  | XGBoost | 0.72 | 0.73 | Best model for this case |
|  | SVM | 0.66 | 0.67 | Moderate performance |
| Benign vs Control (Case 3) | Neural Net | 0.87 | 0.87 | Good class differentiation |
|  | XGBoost | 0.90 | 0.90 | Best performance |
|  | SVM | 0.87 | 0.87 | Similar to NN |
| Multiclass (Cancer/Benign/Control, Case 4) | Neural Net | 0.77 | 0.77 | Modest performance |
|  | XGBoost | 0.86 | 0.85 | Highest overall accuracy |
|  | SVM | 0.81 | 0.78 | Slightly lower ROC and F1 |
|  | CNN | 0.77 | 0.76 | Best for larger datasets; high cancer/control accuracy, low benign |
|  | RNN | 0.79 | 0.79 | Higher F1 for benign, lower for control |

When comparing cancer and control classes (case 1), the models demonstrated varying levels of performance, with Neural Networks outperforming SVM models and even XGBoost. The neural network achieved high precision, scoring 0.96 for both macro and weighted averages, with a weighted F1 score of 0.96, indicating excellent class differentiation. Similarly, XGBoost performed comparably well, with a macro precision average of 0.94, a weighted precision average of 0.94, and a weighted F1 score of 0.94, underscoring its robust classification capability. By contrast, the SVM model showed slightly lower performance. It attained a precision of 0.92 for both the macro and weighted averages and a weighted F1 score of 0.91, suggesting it struggled slightly more in differentiating between the two classes compared to the neural network and XGBoost. Despite these differences, all models demonstrated a solid ability to distinguish between cancer and control samples, most likely due to the large difference in VOC concentration between classes, with the Neural Network outperforming all other models.

In case 2, comparing benign and cancer classes, (shown in Figure 5.1) the models had more difficulty differentiating between the two classes. The neural network had a precision of 0.58 when using a macro average and 0.68 when using the weighted average, with a weighted F1 score of 0.62. Using the same dataset, the SVM had a precision of 0.60 when using the macro average and 0.68 when using the weighted average, with a higher weighted F1 score of 0.67. The XGBoost, however, was most accurate with a macro precision average of 0.69, a weighted precision average of 0.77, and a F1 score of 0.73. When comparing these results with the PCA plot (Figure 2.3) it’s evident that there is confusion in distinguishing cancer and benign classes, with some benign patients having larger VOC readings than some cancer patients. This theory is corroborated in the 80/20 split confusion matrix (Figure 5.2) with a higher proportion of cancer patients being misidentified as benign across all models, regardless of model architecture. With 7 misidentified as cancerous (as compared to 19 benign) in the Neural Network, 10 in the SVM (as compared to 14 benign), and only 5 misidentified in XGBoost models (with 15 misidentified benign) it’s clear that while all models had difficulty predicting cancer patients, due to the Neural Network’s complexity it performed worst in diagnosing cancer patients correctly in this single comparison. To confirm these low readings were not due to the small testing set, a 70/30 split was also performed, available below under Figures 5.4-5.6, with similar accuracies of 0.72 (XGB), 0.63 (NN), and 0.66 (SVM).

During case 3, the comparison of benign tumor nodules and control patients, the accuracy of the models was slightly lower in discriminating between the two classes compared to the cancer versus control comparison. As shown in Figure 4.1, the forward neural network had a precision of 0.87 when using a macro average and 0.87 when using the weighted average, with a weighted F1 score of 0.87. Using the same dataset, the SVM had a precision of 0.88 when using the macro average and 0.88 when using the weighted average, with a higher weighted F1 score of 0.87. The XGBoost, however, performed the best with a macro precision average of 0.89 and a weighted precision average of 0.90 with an F1 score of 0.90. Figures 4.2 and 4.3, generated using an 80/20 split, demonstrate similar themes, with XGBoost having the greatest AUC and a relatively small number of incorrectly identified cases. Despite the overlap between benign and control classes as demonstrated in PC2 (Figure 2.3), the models adjusted well and were able to achieve high accuracies, reinforcing XGBoost’s suitability for VOC-based lung cancer diagnosis, particularly when VOC features are complex and highly variable. In comparisons where the difference between classes is much larger (e.g. control versus cancer), classification becomes easier for all models, and this difference shrinks with SVM performing at or slightly below the level of XGBoost.

The PCA (principal component analysis) plots (Figure 2.1 - 2.3) add further evidence to the viability of using similar VOCs to distinguish between control patients and those with benign and cancerous tumors as there is a clear gradient in PC2 (principal component 2), relatively little overlap between control and cancer classes, and a clear linear trend in PC1 and PC2. However, the multiclass comparison PCA (Figure 2.3) also demonstrates the potential difficulty in distinguishing benign nodules (also seen in the low F1 scores in the train/test evaluations) where the benign class lacks any clear trend and overlaps significantly with control and cancer classes. While this problem is less significant in Figure 2.1, which represents the single comparison between control and cancer classes, there is still overlap with the average expression of PC2 between the two classes being close. However, while there are significant outliers between the two classes (Figure 2.3), the average expression of principal component 2 is higher in Cancer samples, indicating the upregulation of certain “key” VOCs as the distinguishing factor between classes. Between control and cancer classes, there is a clear difference between the two classes in principal component 1, indicating a distinct and easily identifiable difference in VOC levels despite donor variability. Comparing Figures 2.1 and 2.2, it’s also evident that VOCs in patients with cancer and those with benign nodules overlap significantly, while there is clear separation between cancer and control classes. While this may be due to the usage of early-stage cancers where the VOC production mirrors that of benign nodules, it’s most likely due to the inflammation and increase in metabolic processes that benign nodules tend to promote. As seen in Figures 1.3-1.5, Benign and Cancer nodules produce different VOCS, but since they are both “higher” than control classes, the PCA may be grouping these VOCs together without accounting for whether these VOCs are shared.

Taking feature importance values from a Random Forest Model, the most important VOCs used in multiclass differentiation (Case 4) were: 1-Tridecanol, Butyric acid, butyraldehyde, cyclohexylmethanone, and 2-pentanone. Butryaldehyde, a derivative of butane, and 1-Tridecanol, a derivative of acetone, represent derivatives of 2 commonly upregulated VOCs in SW1116 cell lines^2^, ranked 1 and 3 in terms of importance highlighting the importance of VOCs related to membrane synthesis, as they are long-chain alcohols that promote the production of long chain fatty alcohols, which are an essential part of membrane lipid synthesis.

In the multiclass comparison (case 4), the Random Forest Model achieved an accuracy of 0.68 when using a split of 70/30, 0.7 in 80/20, and 0.625 in 90/10. Displaying modest accuracy, while the RF model is not being analyzed for its analysis capacity due to its difficulties in analyzing non-linear complexities, it represents a viable option for VOC analysis in smaller datasets and can potentially be used for generating diagnoses. The difference between XGB and RF models, which both use decision trees, is that XGBoost is a sequential model, while Random Forest is a bagging model. These different architectures allowed for greater accuracy in XGBoost models, as each tree is generated sequentially rather than being generated in parallel.

Despite the unorthodox nature of using CNNs for VOC analysis, it achieved a high accuracy of 0.77 in the 75/25 split, with a F1 score of 0.09 for benign cases, 0.77 for malignant cases, and 0.90 for control cases. CNNs perform best with larger datasets, which is made clear when comparing the F1 score of 0.07 in the relatively small dataset of benign cases and the F1 score of 0.9 and 0.77 in the larger control and cancer datasets. Similar results appeared in the 80/20 split, with an F1 and a recall of 0.09 and 0.11 for benign cases with only 9 support cases. However, despite this lower performance in some categories, the precision for cancer patients hovered around ∼0.7 and ∼0.9 for control cases with overall accuracies of 72% in the 90/10, 77% in the 75/25, 74% in the 70/30, and 71% in the 80/20 split (Figures 7.1-7.9). CNNs provide a viable option for future research aiming to analyze complex nonlinear relationships in large multi-institution datasets, as their model architecture allows for capturing spatial hierarchies and local patterns in data through convolutional layers. This makes them well-suited for processing structured data, such as medical imaging or time-series data, as well as unstructured datasets (as indicated by these results), like VOC profiles from breath analysis. Their ability to handle high-dimensional inputs and integrate domain-specific features can be leveraged to identify subtle patterns associated with distinct classes, improving the relatively high diagnostic accuracy already achieved and building predictive models in diverse clinical settings.

In addition to a CNN, a recurrent neural network or RNN, was implemented using the 3 aforementioned pairwise and multiclass comparison datasets. Unlike the CNN (75/25), the RNN performed best in the 80/20 split (Figure 8.9) with an accuracy of 0.79, a weighted average F1 score of 0.79, and a higher F1 score of 0.33 in benign cases. Despite only having 9 support cases for the benign condition (Figure 8.6) the RNN demonstrates far less confusion between benign and cancer classes than seen in CNN models (Figures 7.2, 7.6, 7.10, 7.14), but suffers from a worse F1 score in control and cancer classes than seen in CNNs(0.84 and 0.67 as opposed to 0.9 and 0.77 respectively). Indicating greater baseline performance, at the cost of higher accuracy in cases with larger training/testing sets, as was observed in CNNs.

## Discussion

For a brief overview of all findings, Tables 1.1 through 1.3 demonstrate key VOCs identified and model accuracies per comparison.

While acetone and ethanol have long been associated with lung cancer, specific compounds associated with lung cancer have been a topic for debate. This study outlines key VOCs in lung cancer utilizing a Random Forest Model identified and ranked several aldehydes as being key markers of cancerous lung tumors, including: Butanal (C4H8O), Heptanal (C7H14O), Pentanal (C5H10O), Decanal (C10H20O), Nonanal (C9H18O), Octanal (C8H16O), and Tridecanal (C13H22O). Key diagnostic targets identified also include 1-Tridecanol, Butyric acid, butyraldehyde, cyclohexylmethanone, and 2-pentanone, with those top 5 alone contributing to roughly 50% of the final decision (in the multiclass comparison) as highlighted in the feature importance value graph in Figure 1.1 – generated by the random forest model. While the XGBoost also produced a feature importance graph based on SHAP values, the most important VOCs were largely the same (except for C6H4O [Crotonaldehyde], which was much higher on SHAP), indicating the importance of these VOCs across model types. Specific VOCs were also identified as specifically important in single comparisons (Figures 1.3-1.6) with C13H22O (an important VOC in the multiclass comparison as well) contributing more than 5 times the positive predictive value of the second most important VOC in the benign and control comparison (2.6 PPV), but only 18th in the benign and cancer comparison (0.17). This indicates that Tridecanal (C13H22O) is an important marker in differentiating patients with benign growths.

C4H8O (Butyraldehyde), on the other hand, is important in identifying cancer classes ranked #1 in cancer versus benign (PPV >1.4) and cancer versus control cases (PPV > 2), but only ranked #9 in terms of PPV in the benign versus control comparison (PPV <0.5). Indicating C4H8O’s importance as a cancer marker, but setting it apart from important VOCs, distinguishing benign patients. C4H8O2 also served as an important cancer marker with a PPV value ranked #2 in the cancer versus benign and cancer versus control cases. C2H4O, interestingly, was only ranked highly in the benign versus cancer comparison, indicating its importance purely being related to its capacity to distinguish cancer patients and benign patients, and causing it to be ranked mid-range in the multiclass comparison.

The upregulation of VOCs identified in this study is likely due to metabolic shifts and oxidative stress characteristic of tumor growth. Using the Human Metabolite Database, we analyzed the top 5 VOCs in the multiclass comparison, identifying pathways associated with these VOCs and their connection to diagnosing malignant and benign growths.

1-Tridecanol, the most important VOC in this comparison, is a long-chain fatty acid made of tridecane substituted by a hydroxy group at position ^12^. It’s been associated with fatty acid metabolism with alterations in lipid metabolism, including those related to long-chain alcohols like C₁₃H₂₂O, which have been implicated in cancer. These metabolic changes can support cancer cell proliferation by influencing membrane composition and signaling pathways that enhance growth and survival. As demonstrated in the multiclass comparison in Figures 1.1 through 1.5, reflecting the inflammation and potential need for detoxification across benign and cancerous growths, and subsequent upregulation of fatty alcohols such as 1-tridecanol^16^.

Butyric acid (C₄H₈O₂) is a short-chain fatty acid involved in histone deacetylase inhibition and regulation of gene expression^18^. It played a larger role in identifying cancer classes when compared to benign and control classes, with it only playing a minor role in differentiating benign and control classes (Figures 1.3-1.5). Due to the Warburg effect, where cancer cells prioritize glycolysis over all other energy production methods, short-chain fatty acids like Butyric acid are consumed in larger quantities, potentially driving up their concentration in cancer cells. Butyric acid can influence cancer progression by blocking cancer stem cell proliferation, slowing cancer development, and activating pathways that suppress tumorigenesis through tumor suppressors like PTEN and P53 ^20^. Its role in regulating immune responses and inflammation also plays a part in cancer biology, potentially contributing to its importance.

Butyraldehyde (C_₄_H_₈_O) is a volatile organic compound involved in metabolic pathways related to fatty acid biosynthesis and oxidative stress. In cancer cells, the dysregulation of oxidative stress is a hallmark feature, and the metabolism of aldehydes like butyraldehyde plays a role in altering the cellular redox environment. Specifically, butyraldehyde is metabolized by aldehyde dehydrogenase (ALDH)^21^, which converts it into butyric acid (another key VOC identified) thereby mitigating oxidative/electrophilic stress in prokaryotic and eukaryotic cells. This process can influence the tumor microenvironment by promoting cell survival and resistance to oxidative damage^21^. Its involvement in metabolic reprogramming and oxidative stress suggests its role in cancer progression and its potential utility as a biomarker in early detection ^16^.

Cyclohexylmethanone (C_₇_H_₁₁_O) may be associated with aromatic compound metabolism, which is relevant in cancer due to the effects of metabolites on cell signaling^16^. Aromatic hydrocarbons and their derivatives are involved in various cellular processes, including apoptosis and angiogenesis, which can be upregulated in cancer cells^19^.

2-Pentanone (C_₅_H_₁₀_O) is a ketone with possible roles in metabolic reprogramming, a hallmark of cancer^16^. Cancer cells often exhibit altered metabolism to support rapid growth and survival, and ketones like 2-pentanone can influence pathways related to energy production, mitochondrial function, and cellular homeostasis^16^. These VOCs are linked to metabolic changes that are crucial for cancer progression, affecting everything from lipid and energy metabolism to oxidative stress and epigenetic modifications.

**Table 1.3:**

| Comparison | Key VOCs (Top 5) | Brief Biological Significance |
| --- | --- | --- |
| Multiclass (Case 4) | 1-Tridecanol, Butyric acid, Butyraldehyde, Cyclohexylmethanone, 2-Pentanone | 1-Tridecanol: long-chain fatty alcohol, membrane lipid metabolism; Butyric acid: short-chain fatty acid, epigenetic regulation, tumor suppression; Butyraldehyde: aldehyde, oxidative stress marker; Cyclohexylmethanone: aromatic metabolism, apoptosis signaling; 2-Pentanone: ketone, altered cancer metabolism |
| Cancer vs Control (Case 1) | Butyraldehyde, Butyric acid, 1-Tridecanol, Undecylaldehyde, Benzaldehyde | Butyraldehyde & Butyric acid: cancer-associated metabolic upregulation; 1-Tridecanol: lipid metabolism; Undecylaldehyde: aldehyde marker of oxidative stress; Benzaldehyde: aromatic aldehyde, metabolic reprogramming |
| Cancer vs Benign (Case 2) | Butyraldehyde, Butyric acid, 1-Tridecanol, 2-Pentanone, Cyclohexylmethanone | Highlights cancer-specific VOCs, overlaps with multiclass; Butyraldehyde and Butyric acid are top differentiators for malignant cells |
| Benign vs Control (Case 3) | 1-Tridecanol, Cyclohexylmethanone, Butyric acid, Acetonylacetone, Acetic acid | Primarily benign growth markers; 1-Tridecanol and Cyclohexylmethanone reflect membrane remodeling and altered cellular metabolism |

Future studies investigating the critical pathways involved in lung cancer differentiation can provide valuable insights into how VOC-based analysis can be refined. By understanding what sets these pathways (and VOCs) apart, researchers can better identify and target VOCs that serve as biomarkers, potentially enhancing the accuracy and effectiveness of VOC analysis for early lung cancer detection and diagnosis.

In addition to identifying and understanding key VOCs in lung cancer using the HMDB^16^, this study sought to establish the most accurate model for diagnosis using various deep-learning methods. In the multiclass comparison, which mirrors the clinical environment where clinicians must differentiate between healthy patients and benign/malignant growths, the XGBoost model had an overall accuracy of 0.86 compared to the SVM and Neural Network’s 0.81 and 0.77, respectively. XGBoost outperformed all other models when utilizing an 80/20 split, with similar themes observed in the 70/30 (NN: 0.77, SVM: 0.77, and XGBoost: 0.82 [accuracy]) and 90/10 ( NN: 0.72, SVM: 0.77, and XGBoost: 0.84) splits. XGB generally performed better or equally compared to other models in both PR AUC and ROC AUC metrics across both train/test splits. It maintained a high average PR AUC (0.74 in both splits), outperforming NN and SVM, and it also showed a strong ROC AUC, especially with a notable improvement between the 70/30 split (0.71) and the 80/20 split (0.87). Even though SVM outperforms XGB slightly in ROC AUC for the 80/20 split, XGB’s robust performance in terms of recall, coupled with its overall high precision and ROC AUC, positions it as the best all-around model for both cancer detection and classification tasks.

In case 1 (Cancer vs Control), all models demonstrated exceptional performance, achieving accuracy rates exceeding 90%. Notably, the Neural Network outperformed the other models, achieving an accuracy of 96%, compared to 94% for the XGBoost model (and 92% in the SVM). This superior performance can likely be attributed to the Neural Network’s intricate architecture, which is better equipped to detect subtle patterns that the relatively simpler XGBoost model may overlook.

Most models suffered in case 2 (Benign versus Cancerous, with substantially lower F1 scores across all analysis methods. The XGBoost model outperformed the Neural Network and SVM at the 80/20 split with an accuracy of 72% as compared to 66% (SVM) and 61% (NN) (Figure 5.1). This echoes themes found in case 1 and case 4 (multiclass), where the relatively small sample size of the dataset and complex architecture hurt the NN’s ability to identify trends and giving it an F1 score of only 0.46 in the benign classes compared to the XGB’s 0.6. The performance drop regardless of model architecture may also be due to the overlap in upregulated VOCs between benign nodules and lung cancers; it’s more likely due to donor variability (unaccounted for in an anonymized dataset), and the highly variable nature of benign nodule and lung cancer VOC production. Age, fitness, site inflammation, and cancer stage all have a bearing on VOC production, and lower readings in the cancer class are most likely the result of an earlier-stage NSCLC. Future studies could incorporate these factors into analysis and use a larger benign nodule dataset to ensure that more complex models, such as NNs, are better able to identify key trends.

In addition to assessing the performance of a standard Neural Network (NN), this study sought to determine whether its limitations could be addressed through the implementation of Recurrent Neural Network (RNN) and Convolutional Neural Network (CNN) architectures. The CNN performed marginally worse than the neural network, but demonstrated a higher. When evaluating based on the multiclass comparison (case 4), the CNN (convolutional neural network) achieved a weighted F1 and recall score of 0.71 in the 70/30 split, with an F1 score of 0.07 for benign cases and 0.74 and 0.85 for malignant and control cases, respectively (Figure 7.5). While possibly a result of only 17 benign cases being used in the test set, similar trends emerged in the 75/25 split, with a weighted F1 and recall score of roughly 0.76 and only 0.09 for the benign cases. However, the model performed far better in cancer and control classes, with an F1 score of 0.77 and 0.9, and an accuracy of 0.77. These results in the 75/25 split are mirrored in figures 7.11 and 7.12 with a ROC AUC of 0.54, 0.84, and 0.96 in benign, cancer, and control classes, respectively, and a PR AUC of 0.12 in benign cases, 0.81 in cancer cases, and 0.96 in control cases. Demonstrating the high accuracy between cancer and control cases, and underscoring an even greater potential in a larger dataset.

The RNN (recurrent neural network) performed marginally better than the CNN across the board, with an average precision and accuracy of 0.8 and 0.79 in the 80/20, and 0.74 and 0.73 in the 70/30. While the CNN had higher F1 scores (using the 75/25 split) of 0.9 and 0.77 in control and cancer cases when compared to the RNN’s 0.82 and 0.72, the RNN outperformed the CNN in the 90/10 split (0.77 compared to 0.72) and 80/20 split (0.79 compared to 0.71). In addition, the RNN had a far higher F1 score for the Benign cases (around 0.3), demonstrating how it compensates better for the overlap between cancer and benign cases observed and the smaller number of cases observed in the benign class. The RNN outperformed the NN in the 80/20 split, achieving an accuracy of 0.79 and F1 scores of 0.33 for benign cases and 0.78 for cancer cases, compared to the NN’s corresponding values of 0.77, 0.11, and 0.74. However, a trade-off was observed, as the NN performed better in control cases, achieving an F1 score of 0.92 compared to the RNN’s 0.89. Despite this, the RNN maintained a higher overall accuracy.

Although CNNs and RNNs demonstrated relatively low accuracy in this case, they show promise for future applications in VOC-based diagnostics, especially for conditions like lung cancer. CNNs, typically associated with image recognition, can learn complex patterns from sequential data like VOC values, making them adaptable to subtle variations in exhaled compounds linked to different disease states. RNNs provided a greater baseline performance, allowing for nuanced and accurate analysis and behaving similarly to CNNs, providing a highly viable alternative to the NNs evaluated in this paper. As datasets grow in size and diversity, CNN/RNNs may capture nuanced features and correlations more effectively than their less complex counterparts, potentially increasing their predictive power and preventing negative results as seen with the patients with benign tumors in this study. Moreover, advancements in model architecture and data preprocessing, along with integration with other machine learning techniques, could improve the reliability of CNN/RNNs for VOC analysis by compensating for their propensity to overfit and low accuracy at small sample sizes.

## Conclusion

This study successfully identified key VOCs in identifying lung cancer and evaluated different models’ accuracy when used in the diagnosis and prediction of lung cancer (Tables 1.2, 1.3). Future research can further investigate the biological pathways that contribute to the importance of specific aldehydes in lung cancer and the biochemical reactions involved in secreting the VOCs associated with lung malignancies. In addition, more work can be done to discover VOCs present in other respiratory conditions, including infections and smoking-related diseases, to broaden the scope of breath analysis as a method for disease diagnosis. Future research can target the VOCs identified in this paper to generate rapid diagnoses from common breath analysis machines using fewer chemical signatures and build future AI-guided diagnostic methods on the ones outlined by this paper and the code provided. With cigarette smoking being reported in 75-90% of lung cancer cases diagnosed and 90% of cancers detected in those with a family history of the condition being early stage, more research should be done to establish methods encouraging greater acceptance of widespread non-invasive screening in high-risk populations^11^.

This study has several limitations that should be considered when interpreting the results. The dataset was fairly limited, with 427 samples obtained from patients at the University of Louisville, with only 65 benign cases compared to roughly 200 control and 150 malignant cases. By using multi-institutional data and performing longitudinal studies on patients diagnosed with lung cancer, performance could be significantly improved, accounting for variability across NSCLC stages and benign cases. Furthermore, due to the anonymization of the dataset, demographic factors were not considered when building the diagnostic models^10^, and since only the chemical formulas were supplied by the Dr. Fu Lab, the true names of the chemicals were determined using existing literature.

Future research should involve a larger sample from multiple institutions and geographic locations to include a more diverse study population and ensure that models perform similarly across all patients and geographic locations.

Future studies should perform hyperparameter tuning and assess the viability of hybrid learning models for interpretation to generate more accurate readings and ensure clinical efficacy. Models such as SVM and XGBoost provided excellent baseline performance, bridging efficiency with high accuracy, while more complex models suffered from lower baseline performance but were better suited for non-linear and complex relationships. RNNs provided a great compromise, achieving similar ∼0.3 F1 scores in the difficult benign class while performing similarly to SVM and XGBoost models.

Work should also be performed evaluating whether certain models, such as XGBoost and CNN/RNNs, can be combined to compensate for their weaknesses, with CNN/RNNs providing analysis of nuanced themes while taking advantage of the high baseline performance of XGBoost models. CNNs can also be integrated for a more holistic approach, using imaging and clinical notes to be analyzed in concert with VOC values and bridging structured and unstructured data, allowing for greater accuracy and a broader breadth of data to be analyzed.

AI models show great promise for diagnosing lung cancer using volatile organic compounds (VOCs), each model type bringing unique advantages (Table 1.2). While machine learning models like decision tree-based models and support vector machines offer interpretability and high baseline performance, deep learning models using Neural Networks excel at capturing nuanced patterns in complex VOC data. The path forward likely lies in innovative hybrid approaches that combine these strengths, creating robust, accurate, and adaptable diagnostic tools. By minimizing the number of features and targeting only “key” VOCs identified in this paper (Figures 1.1-1.5), researchers can streamline future analysis and simplify collection methods, catering only to compounds identified as significant in their comparison. Starting from potentially hundreds of VOCs, researchers can target pathways associated with significant VOCs identified in this paper and dive deeper into the specific reasons for these compounds’ upregulation (Table 1.3, Figures 1.1-1.5).

In advancing interdisciplinary collaboration and optimizing AI techniques for VOC analysis, we edge closer to a future where non-invasive, AI-driven VOC diagnostics transform lung cancer detection and patient outcomes.

## Data Availability

All data produced in the present study are available upon reasonable request to the authors

https://github.com/sam-uofl/Dr.FuLab

## Acknowledgements

Dr. Fu Lab University of Louisville

## Supplementary Materials

**Table 1.1 (sourced from HMDB):**
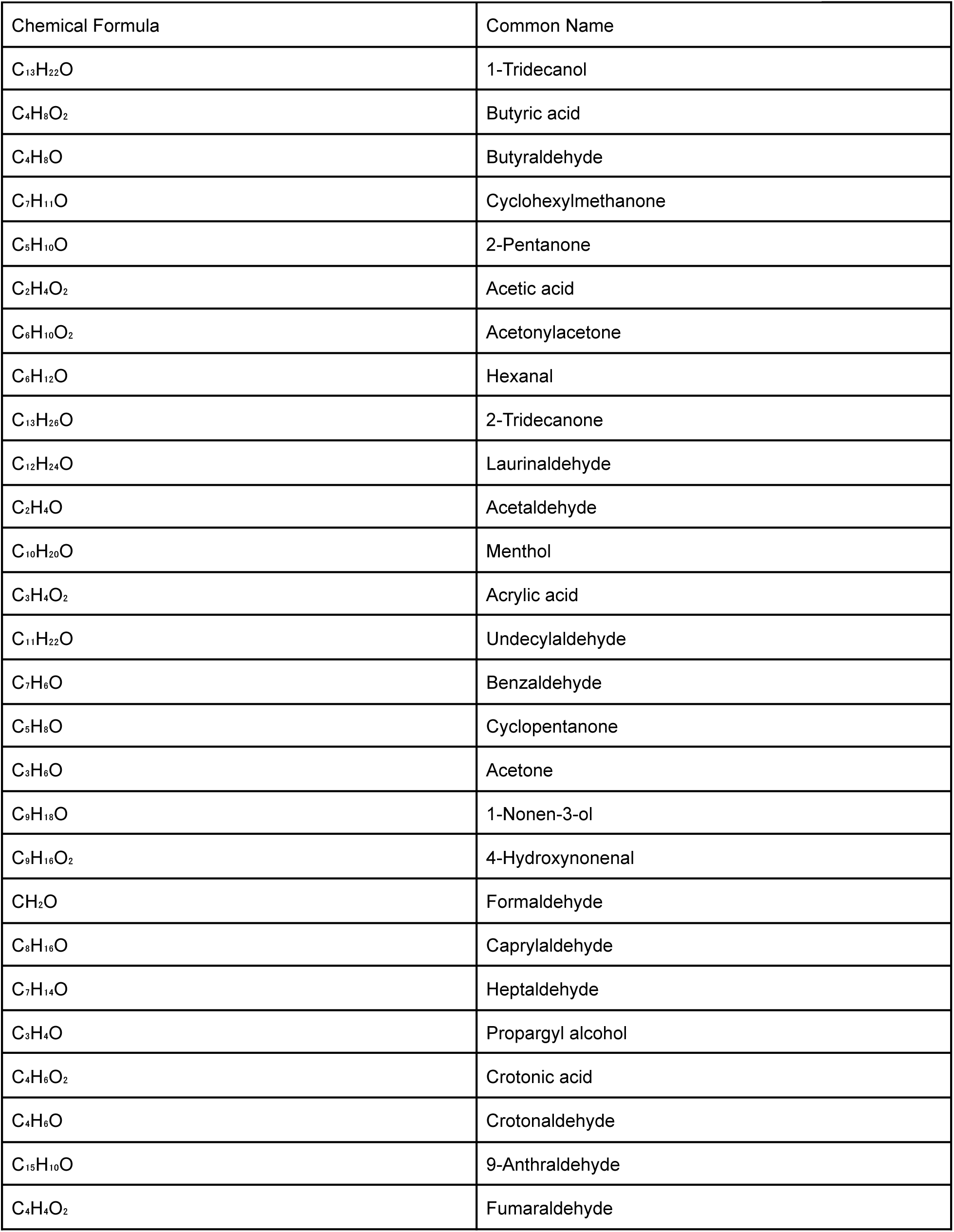
Note that some compounds may have multiple common names or synonyms:

**Table 1.2:**

| Comparison (Case) | Model | Accuracy | Weighted F1 | Notes |
| --- | --- | --- | --- | --- |
| Cancer vs Control (Case 1) | Neural Net | 0.96 | 0.96 | Outperformed all other models |
|  | XGBoost | 0.94 | 0.94 | Strong performance overall |
|  | SVM | 0.92 | 0.91 | Slightly lower than NN/XGB |
| Cancer vs Benign (Case 2) | Neural Net | 0.61 | 0.62 | Low F1 due to class overlap |
|  | XGBoost | 0.72 | 0.73 | Best model for this case |
|  | SVM | 0.66 | 0.67 | Moderate performance |
| Benign vs Control (Case 3) | Neural Net | 0.87 | 0.87 | Good class differentiation |
|  | XGBoost | 0.90 | 0.90 | Best performance |
|  | SVM | 0.87 | 0.87 | Similar to NN |
| Multiclass (Cancer/Benign/Control, Case 4) | Neural Net | 0.77 | 0.77 | Modest performance |
|  | XGBoost | 0.86 | 0.85 | Highest overall accuracy |
|  | SVM | 0.81 | 0.78 | Slightly lower ROC and F1 |
|  | CNN | 0.77 | 0.76 | Best for larger datasets; high cancer/control accuracy, low benign |
|  | RNN | 0.79 | 0.79 | Higher F1 for benign, lower for control |

**Table 1.3:**
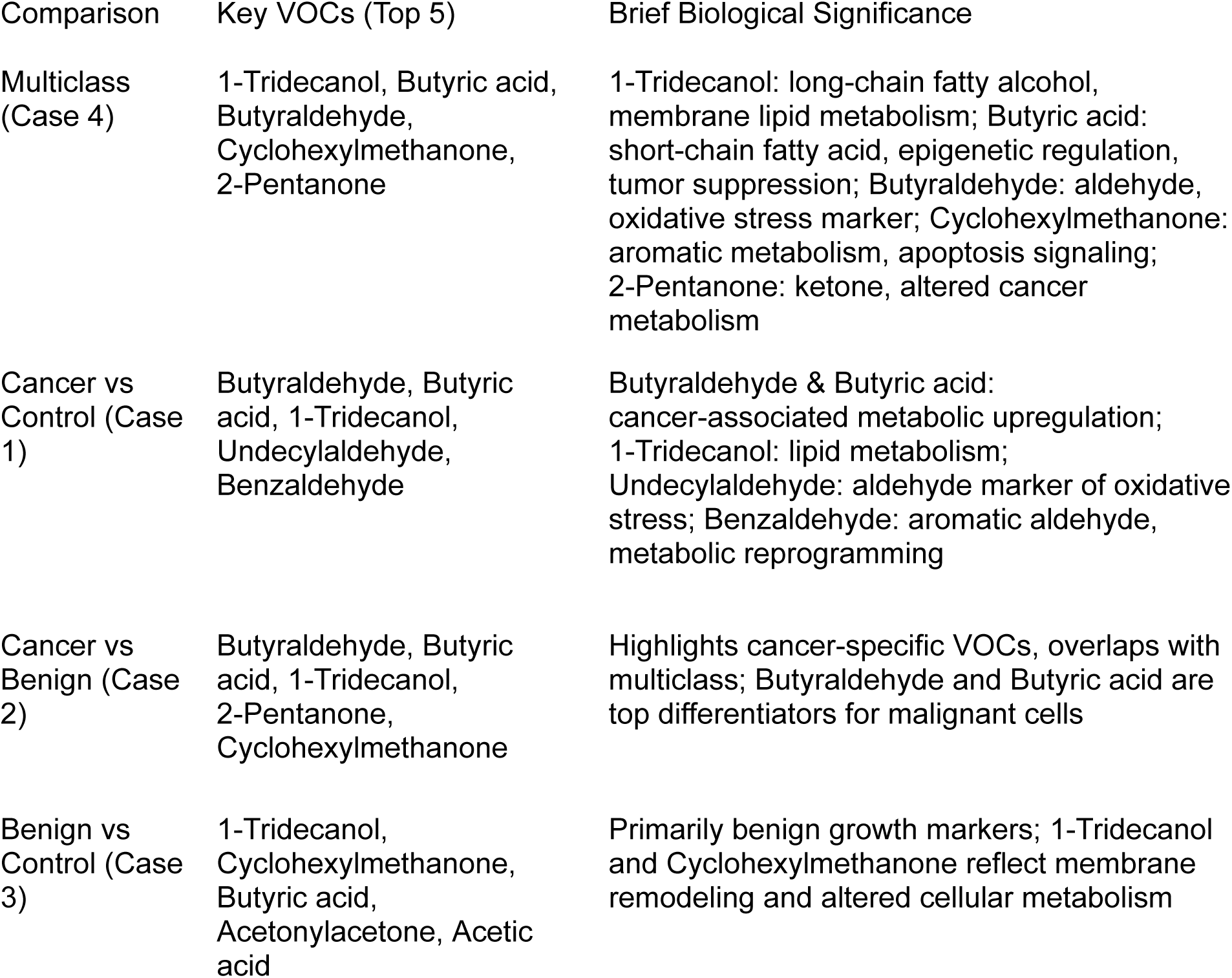

**Figure 1.1.**
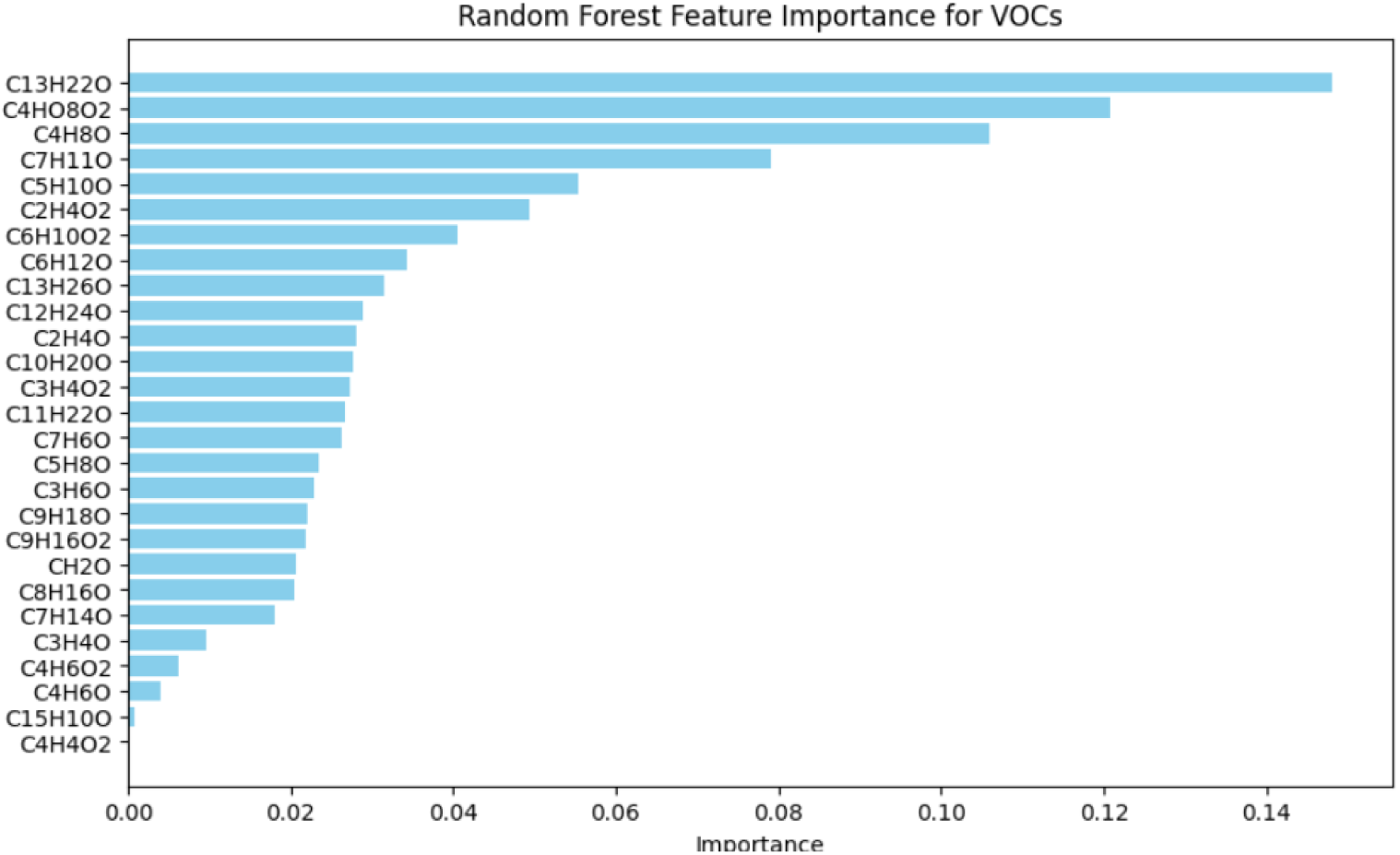
Random Forest Generated Feature Importance Values in Case 4 (Multiclass).

**Figure 1.2.**
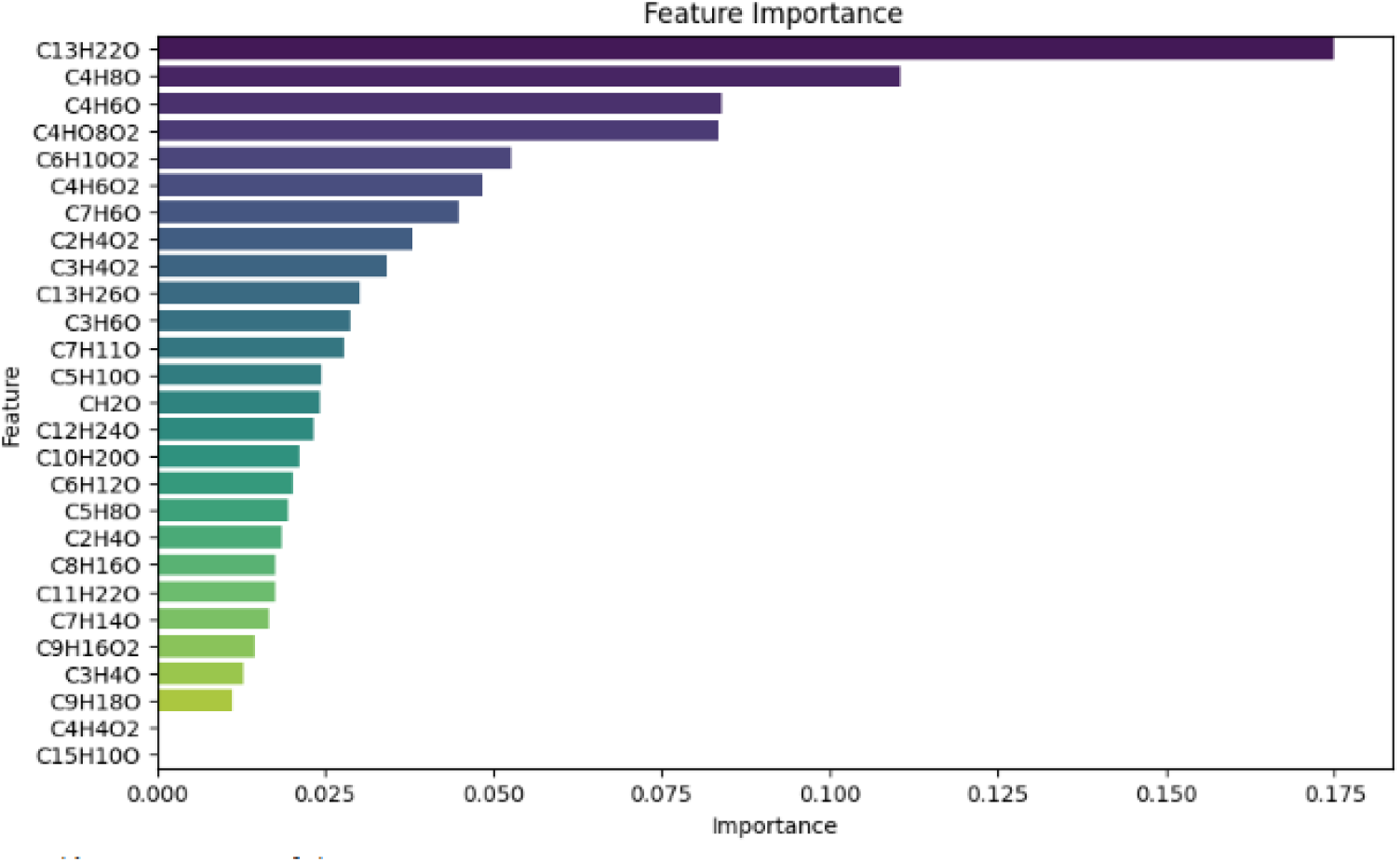
XGB SHAP-based Feature importance values in Case 4 (Multiclass).

**Figure 1.3.**
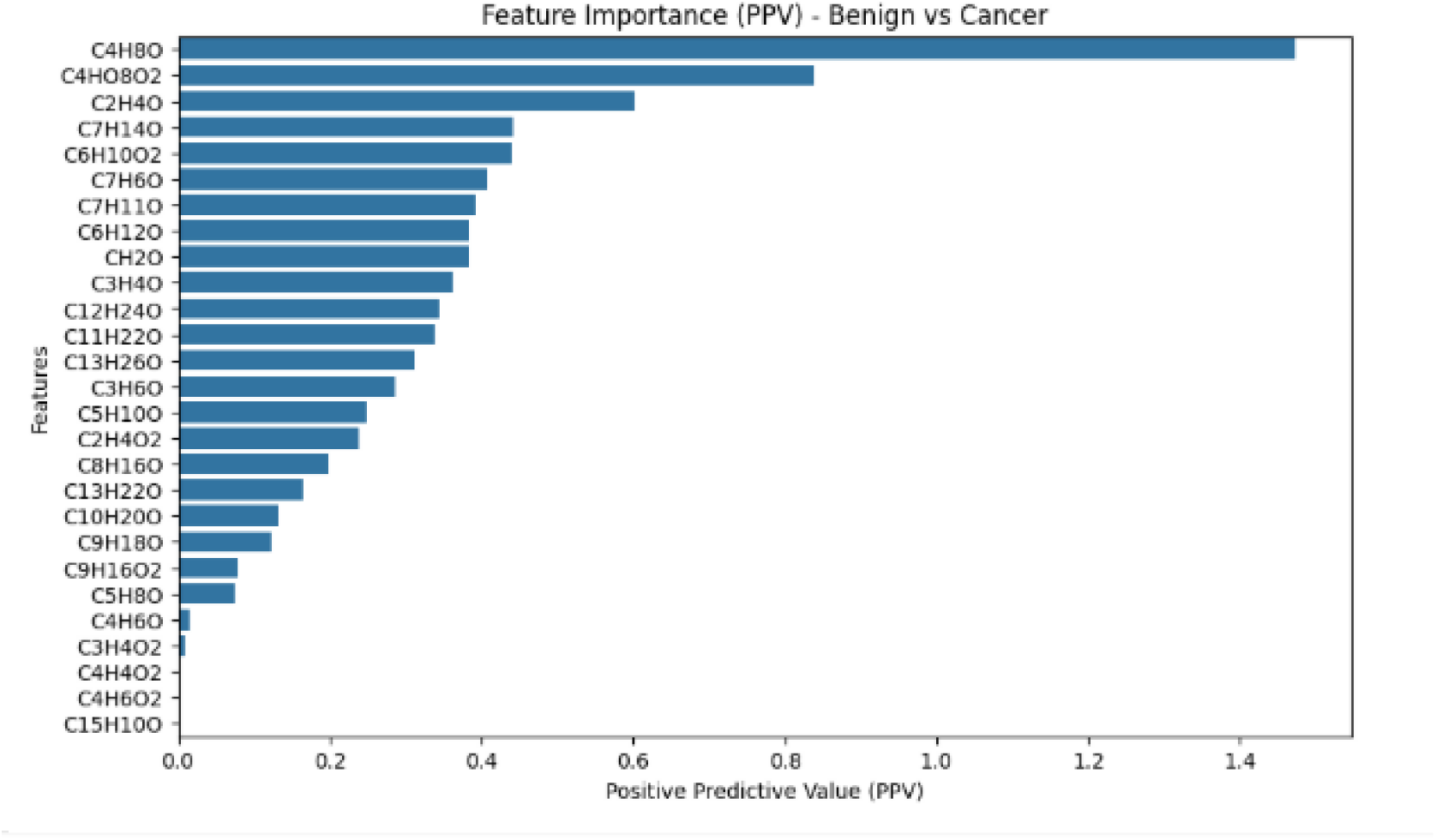
PPV Benign and Cancer.

**Figure 1.4.**
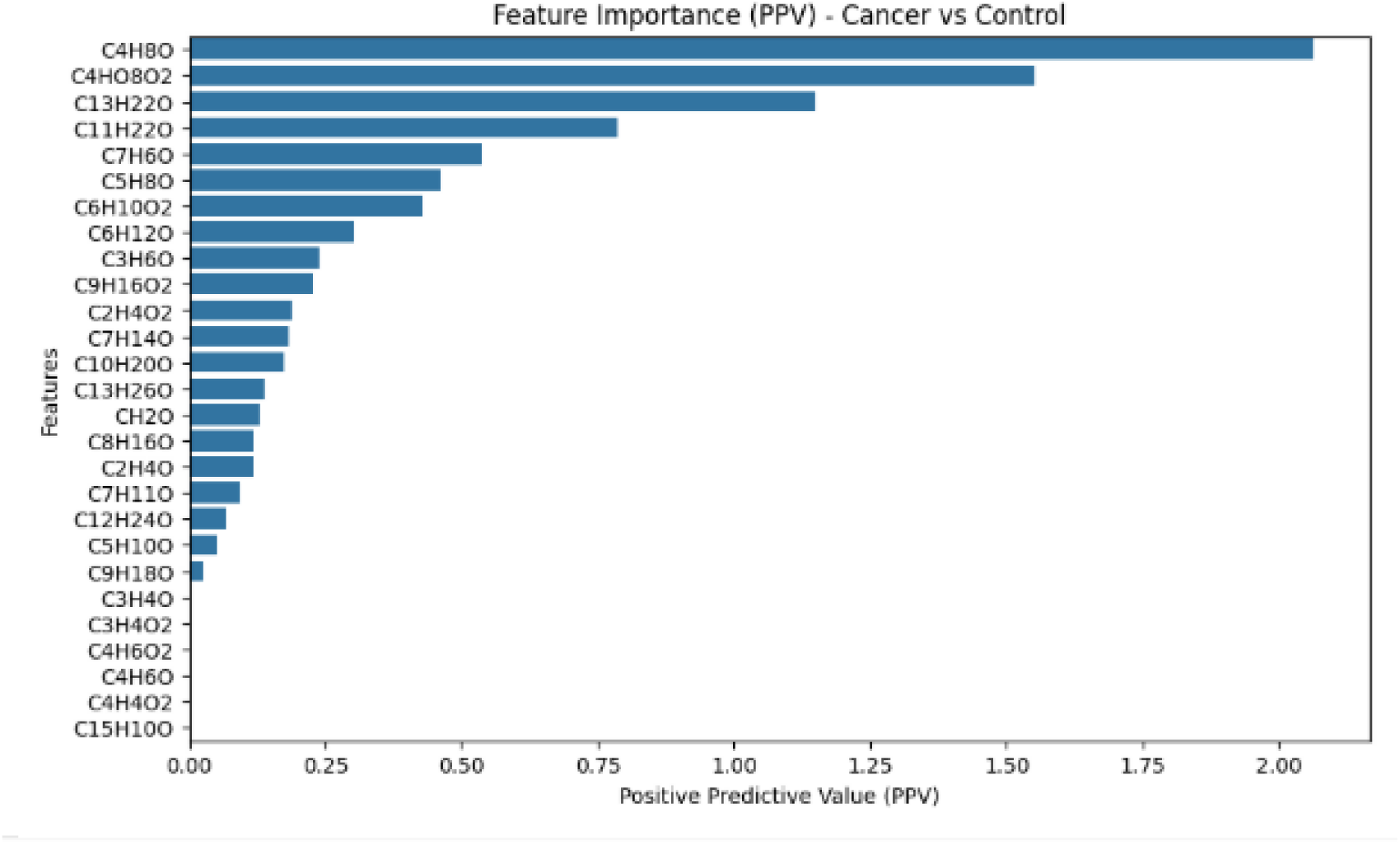
PPV Cancer and Control.

**Figure 1.5.**
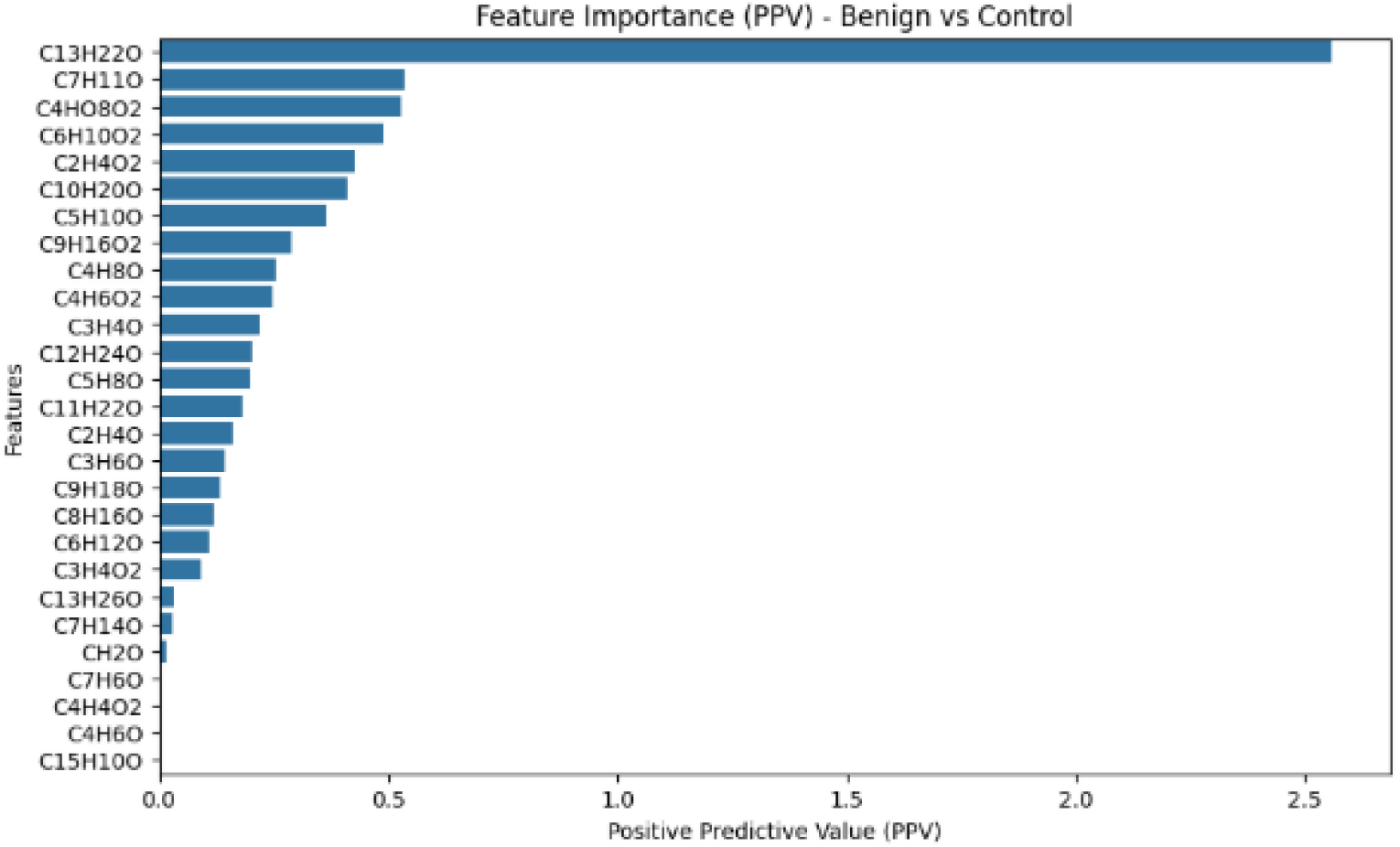
PPV Benign and Control.

**Figure 2.1.**
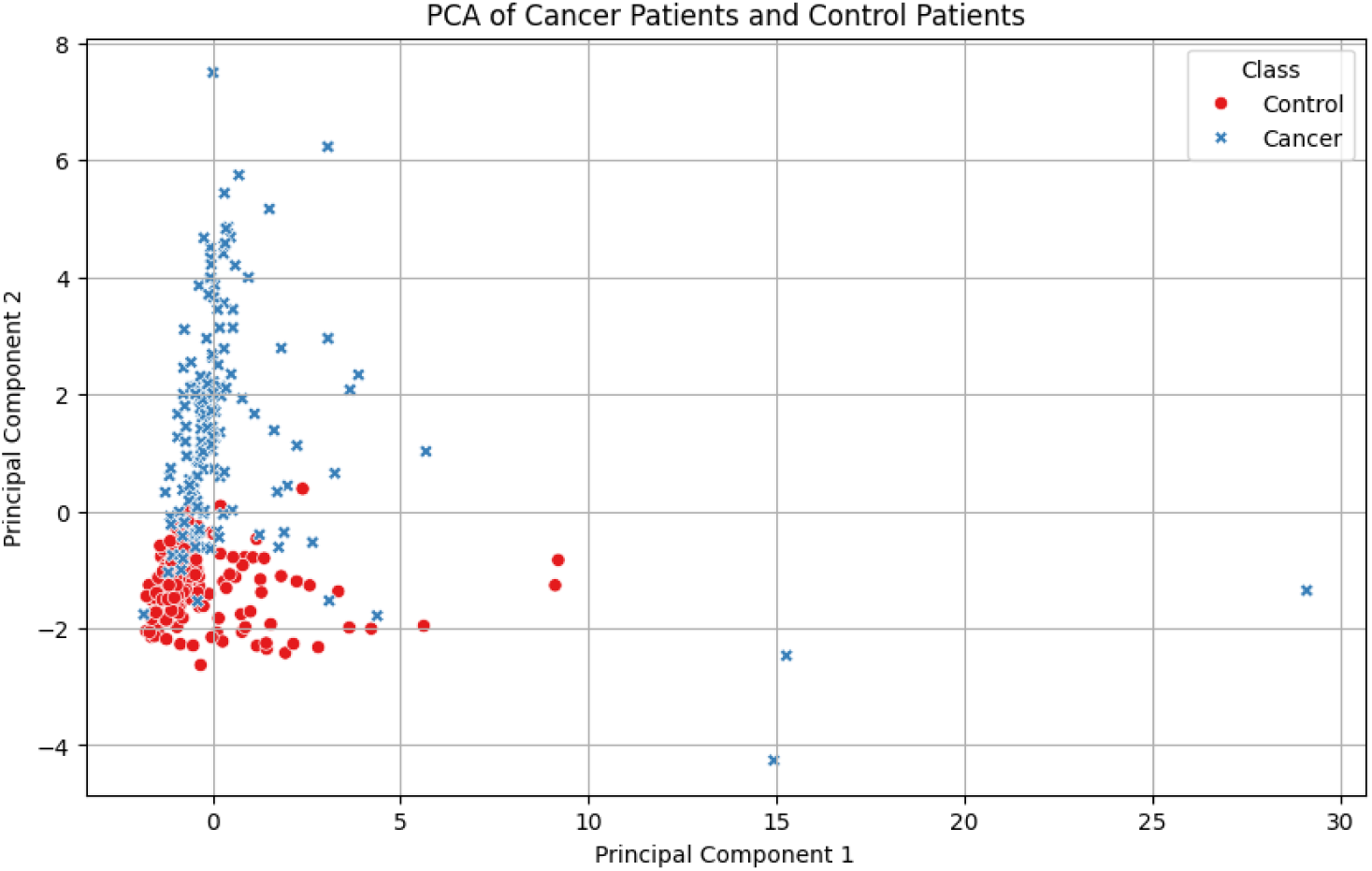
PCA Between Cancer and Control Patients.

**Figure 2.2.**
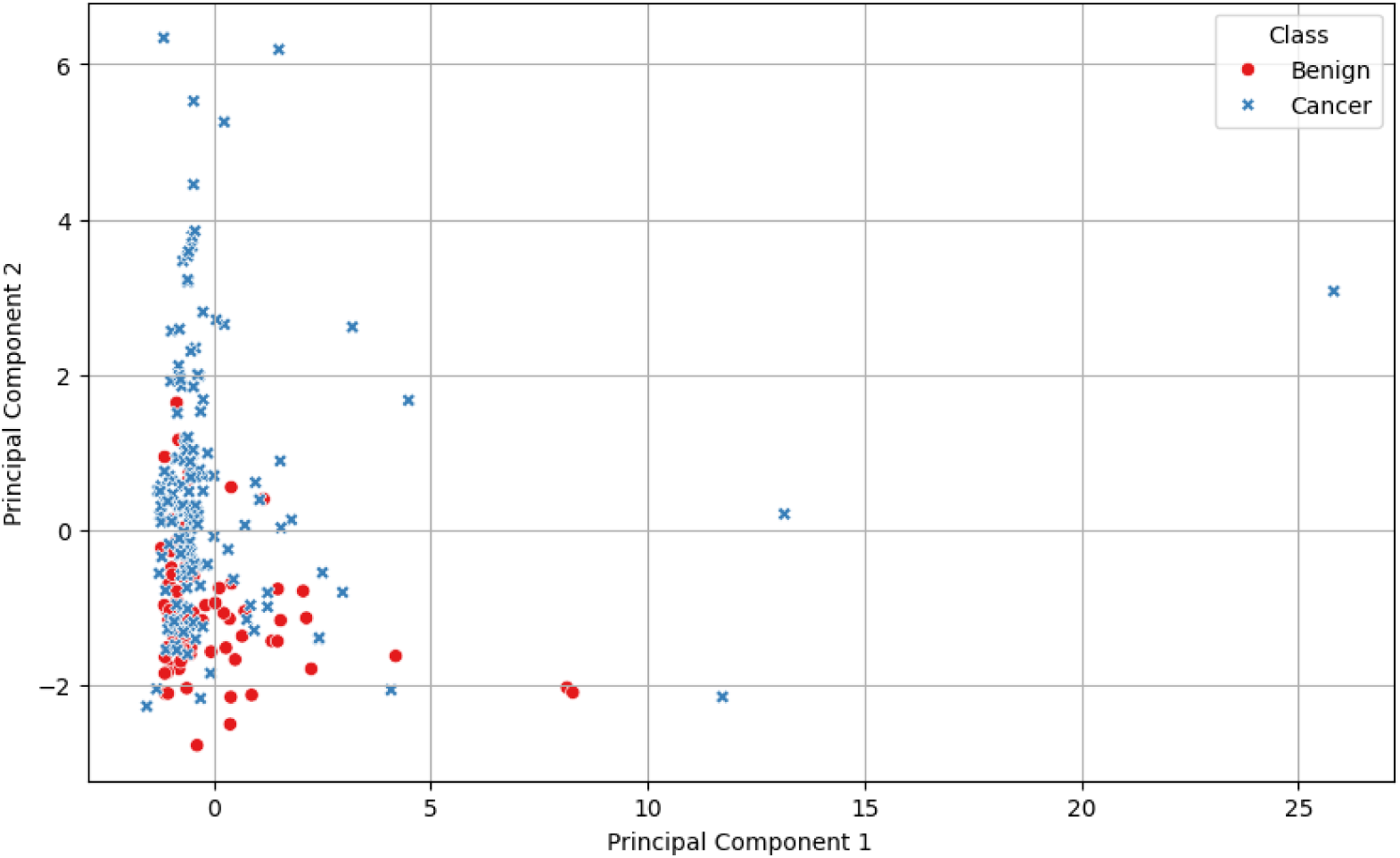
PCA Between Cancer and Benign Patients.

**Figure 2.3.**
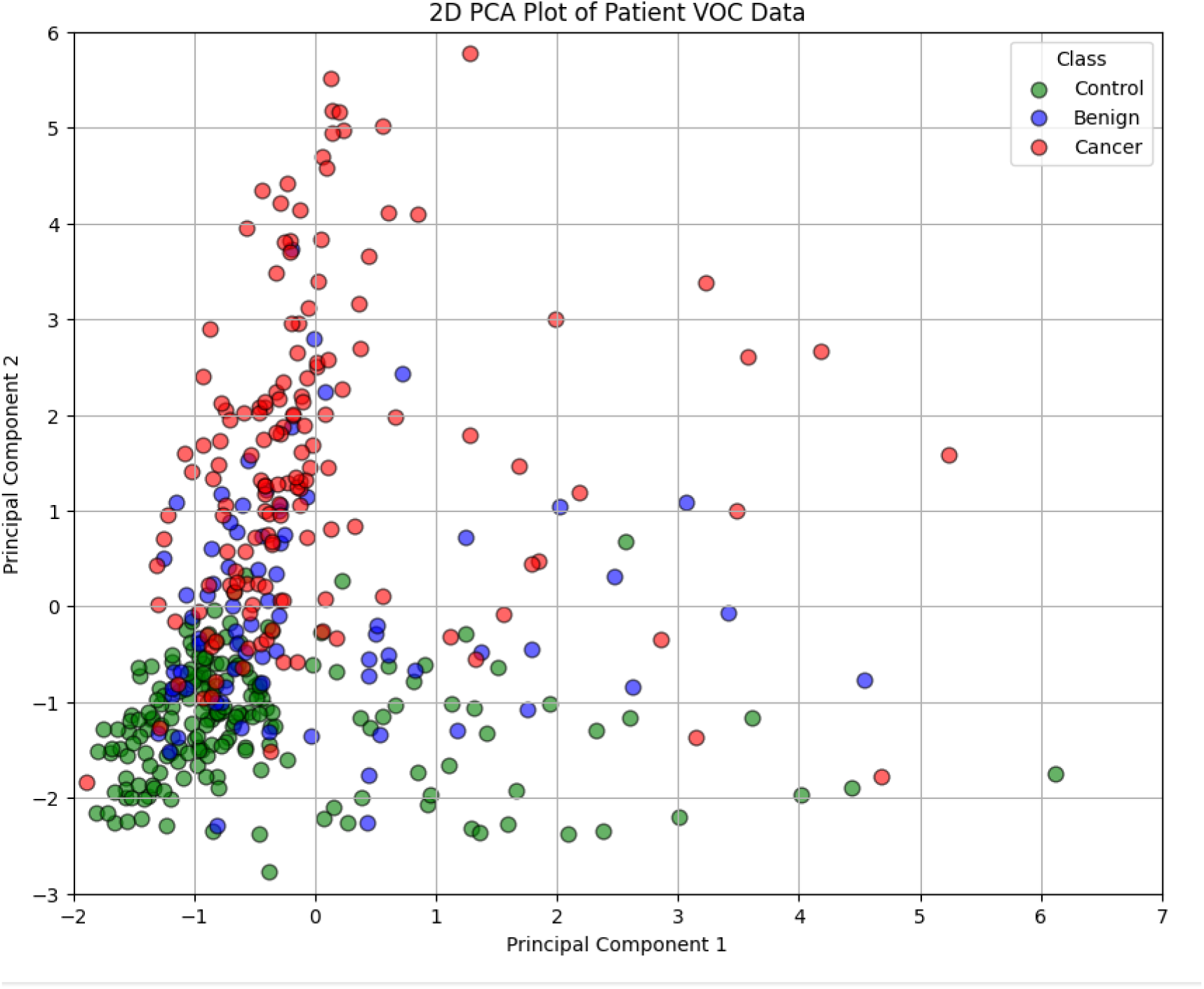
Multiclass PCA (Case 4).

**Figure 3.1.**
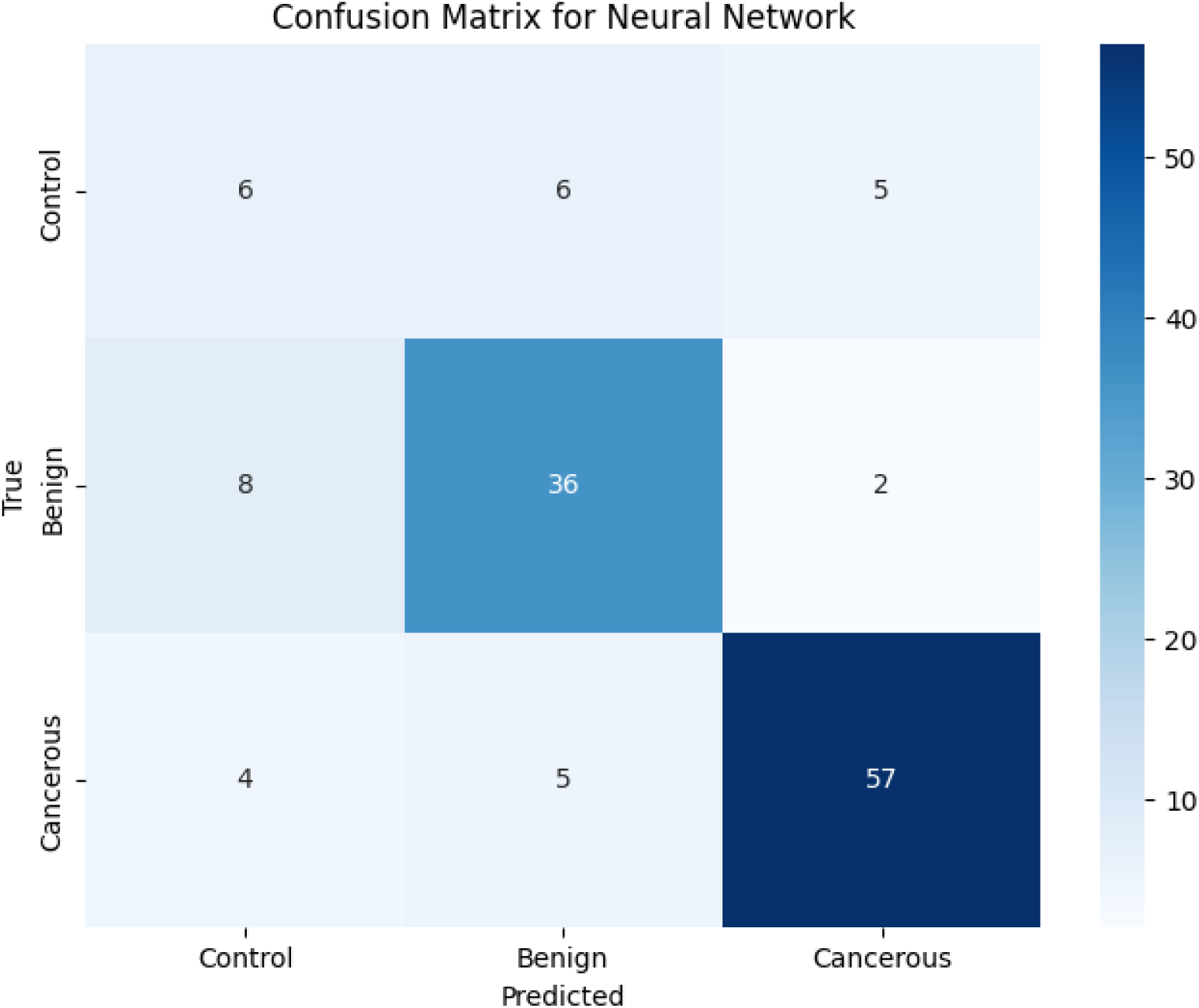
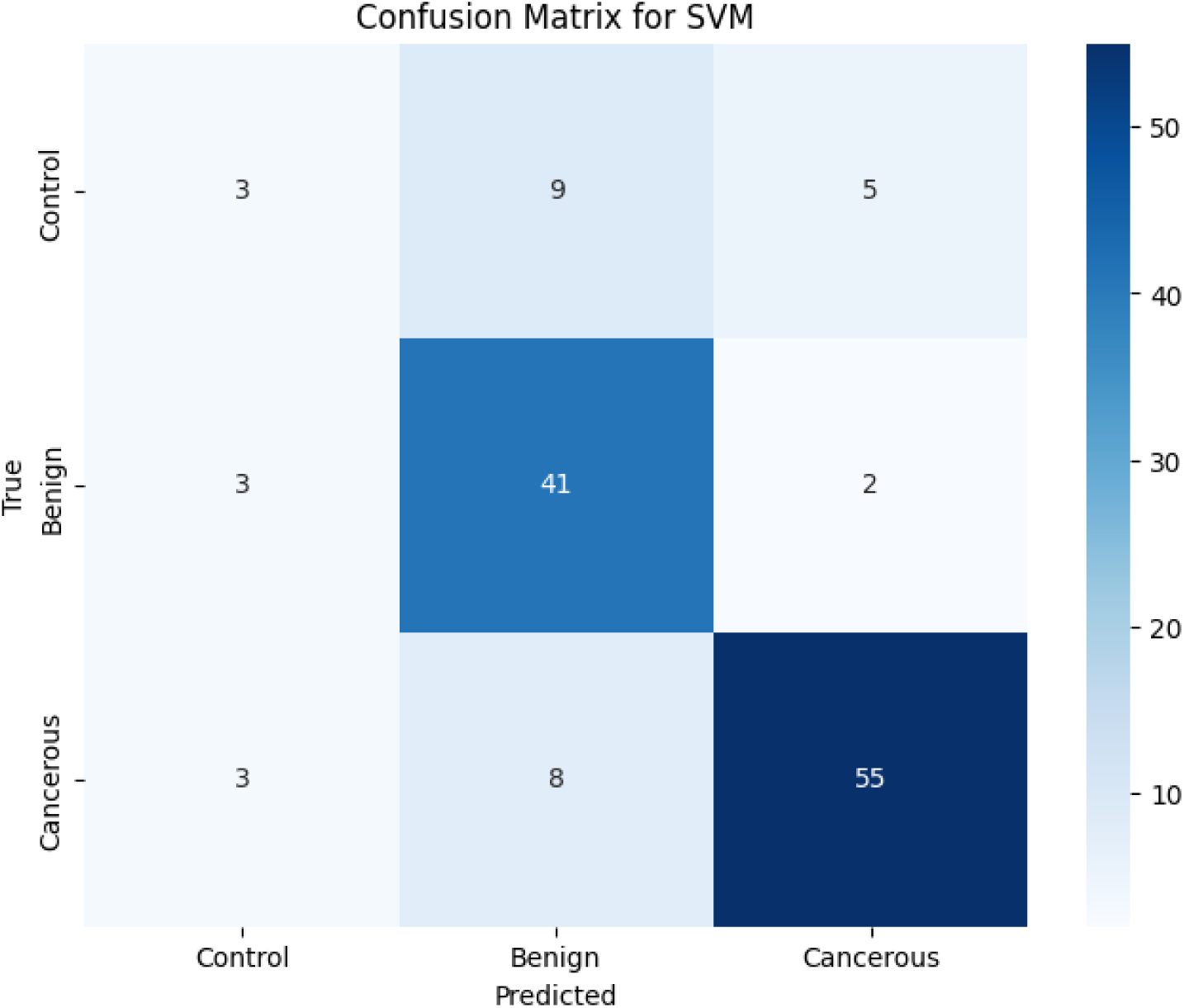
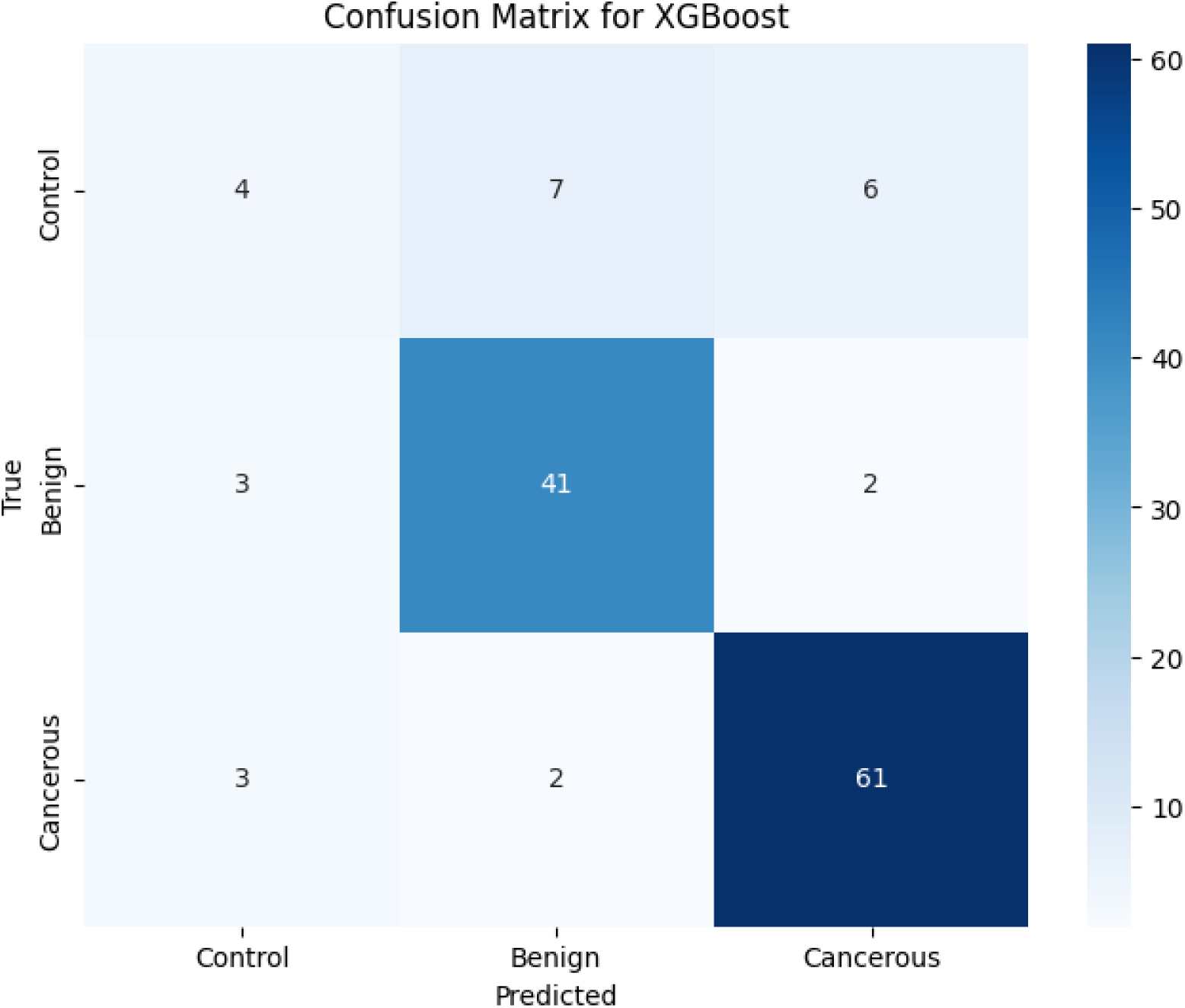
Case 4 (Multiclass) Confusion Matrices.

**Figure 3.2.**
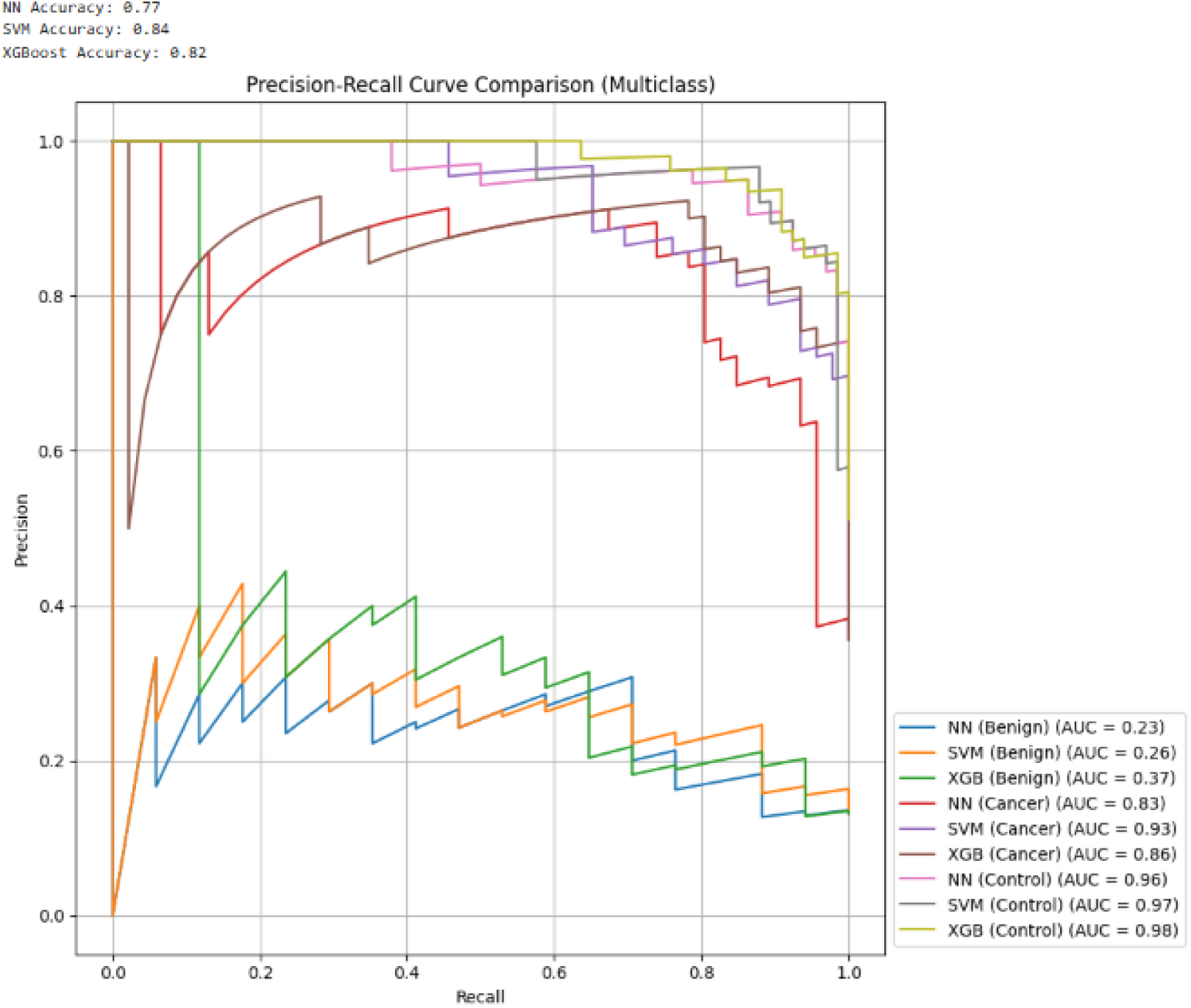
Multiclass PR Curve (70/30).

**Figure 3.3.**
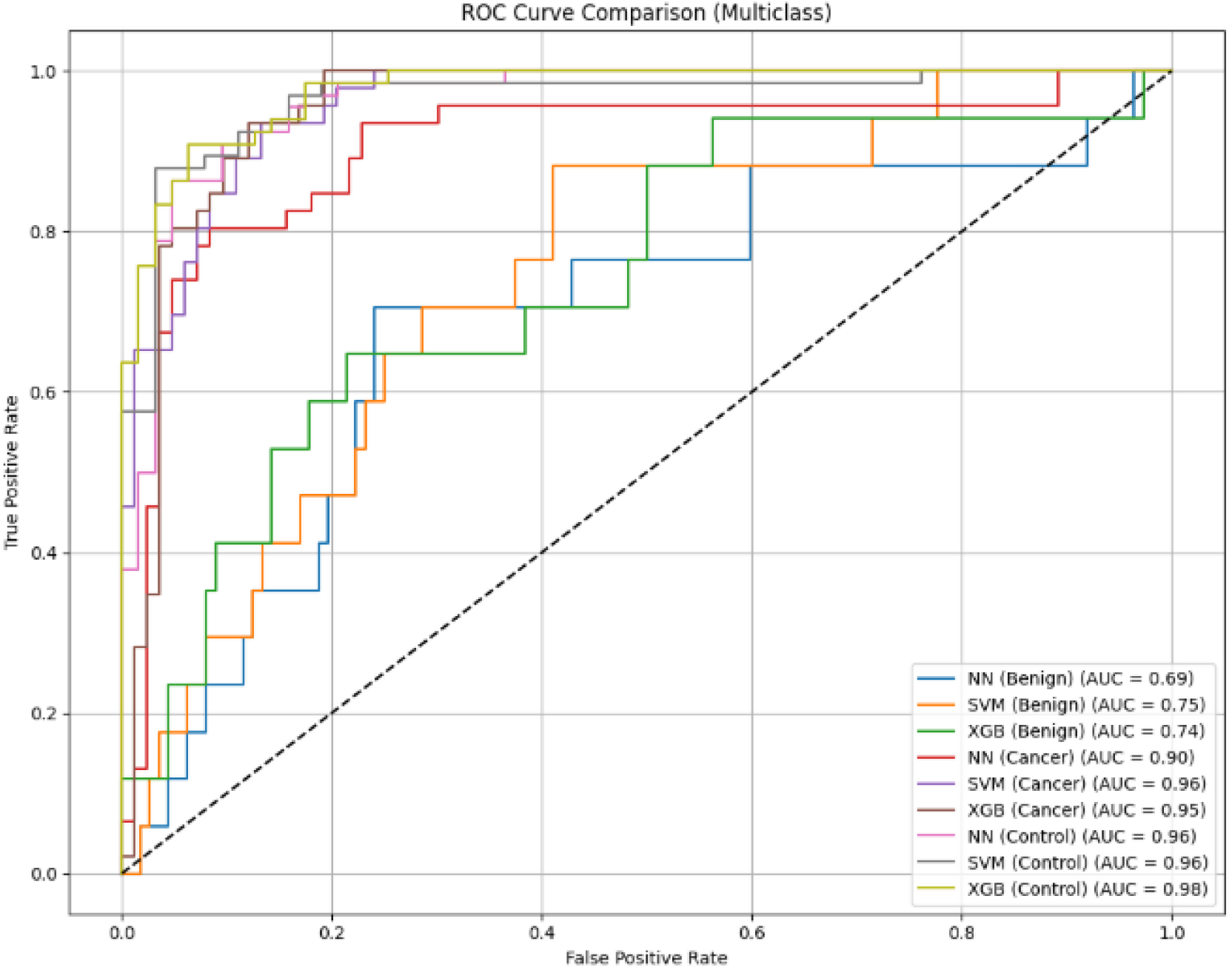
Multiclass ROC Curve (70/30).

**Figure 3.4.**
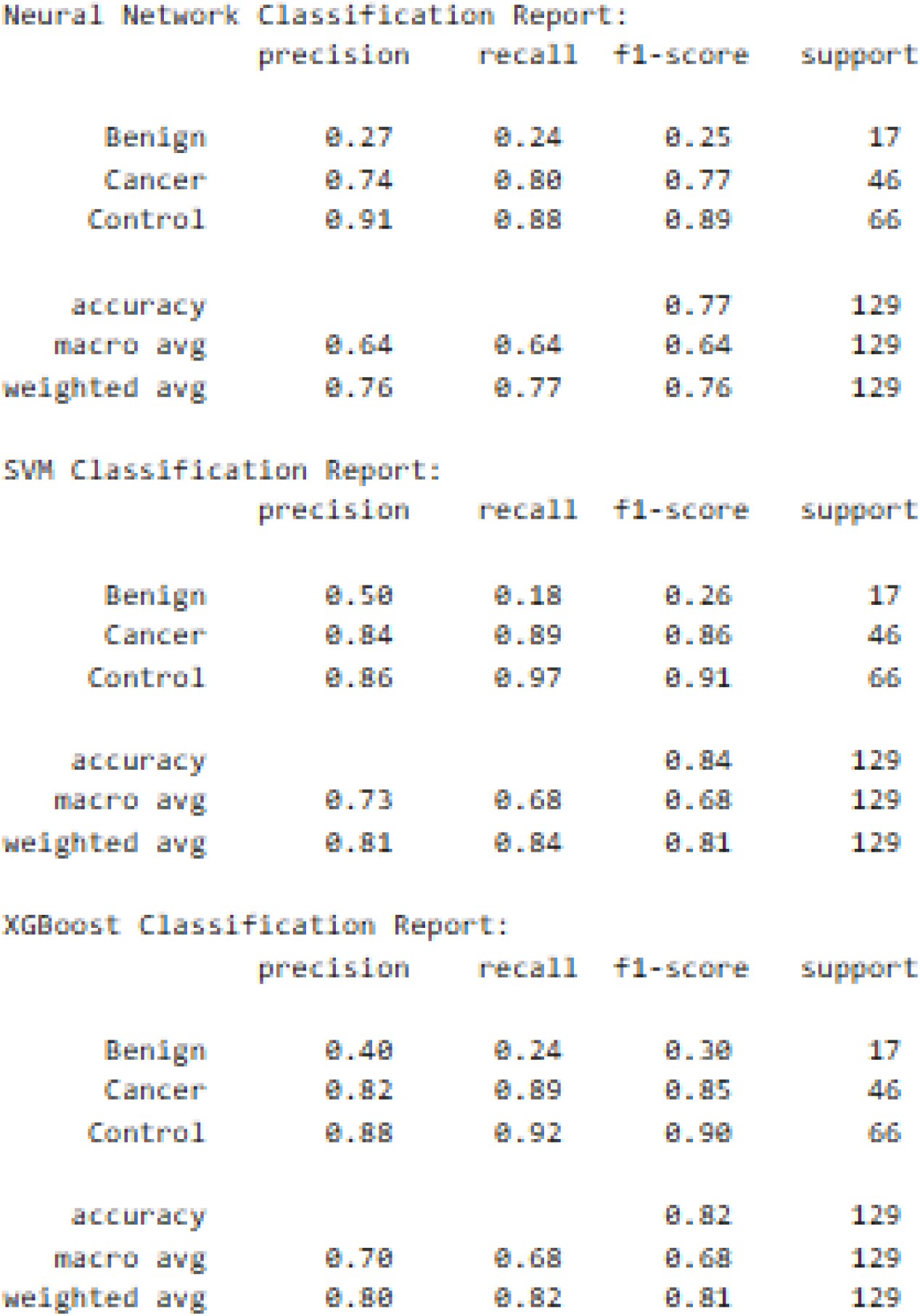
Multiclass Classification Report (70/30).

**Figure 3.5.**
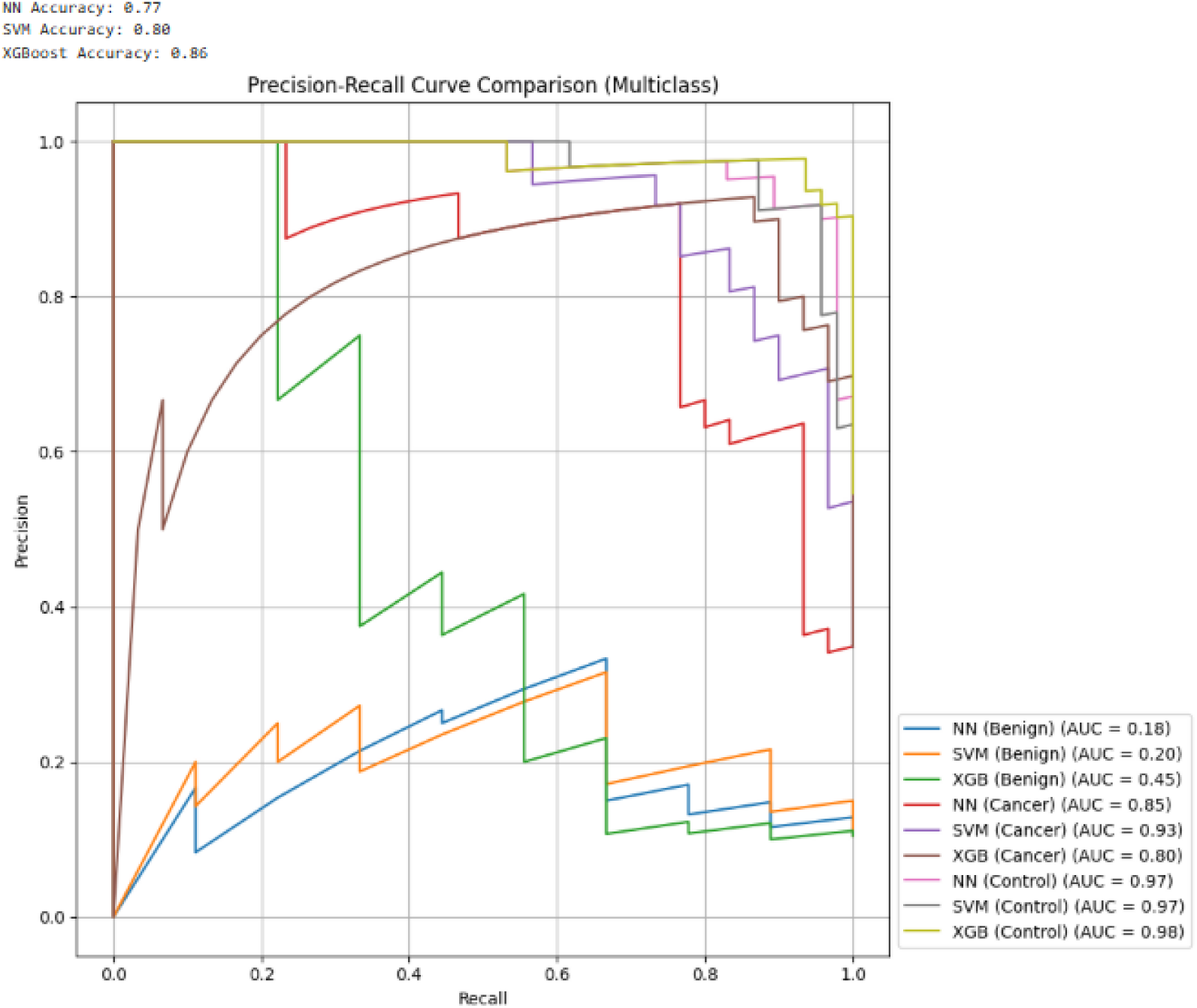
(80/20) Multiclass PR Curve (80/20).

**Figure 3.6.**
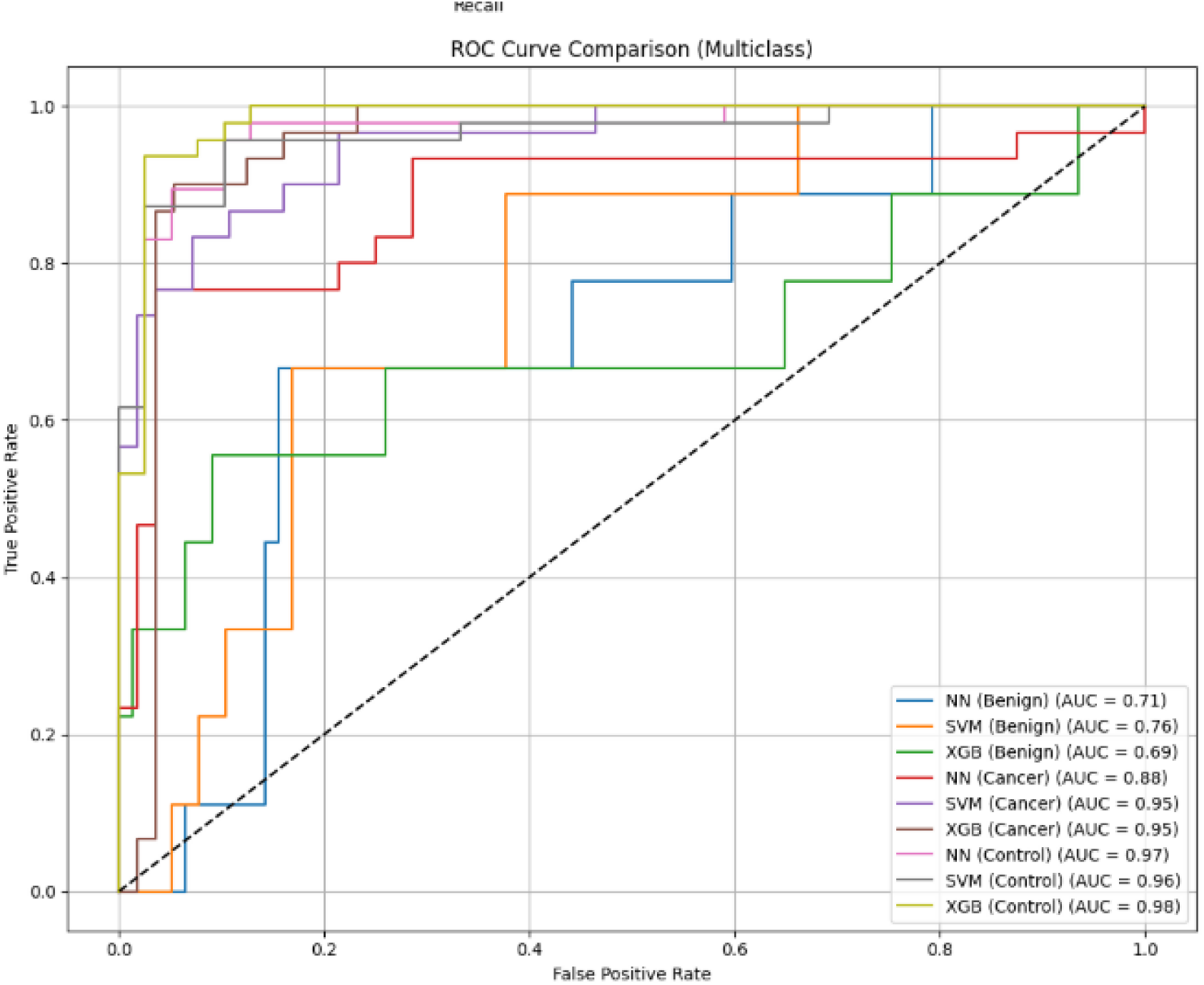
Multiclass ROC Curve (80/20).

**Figure 3.7.**
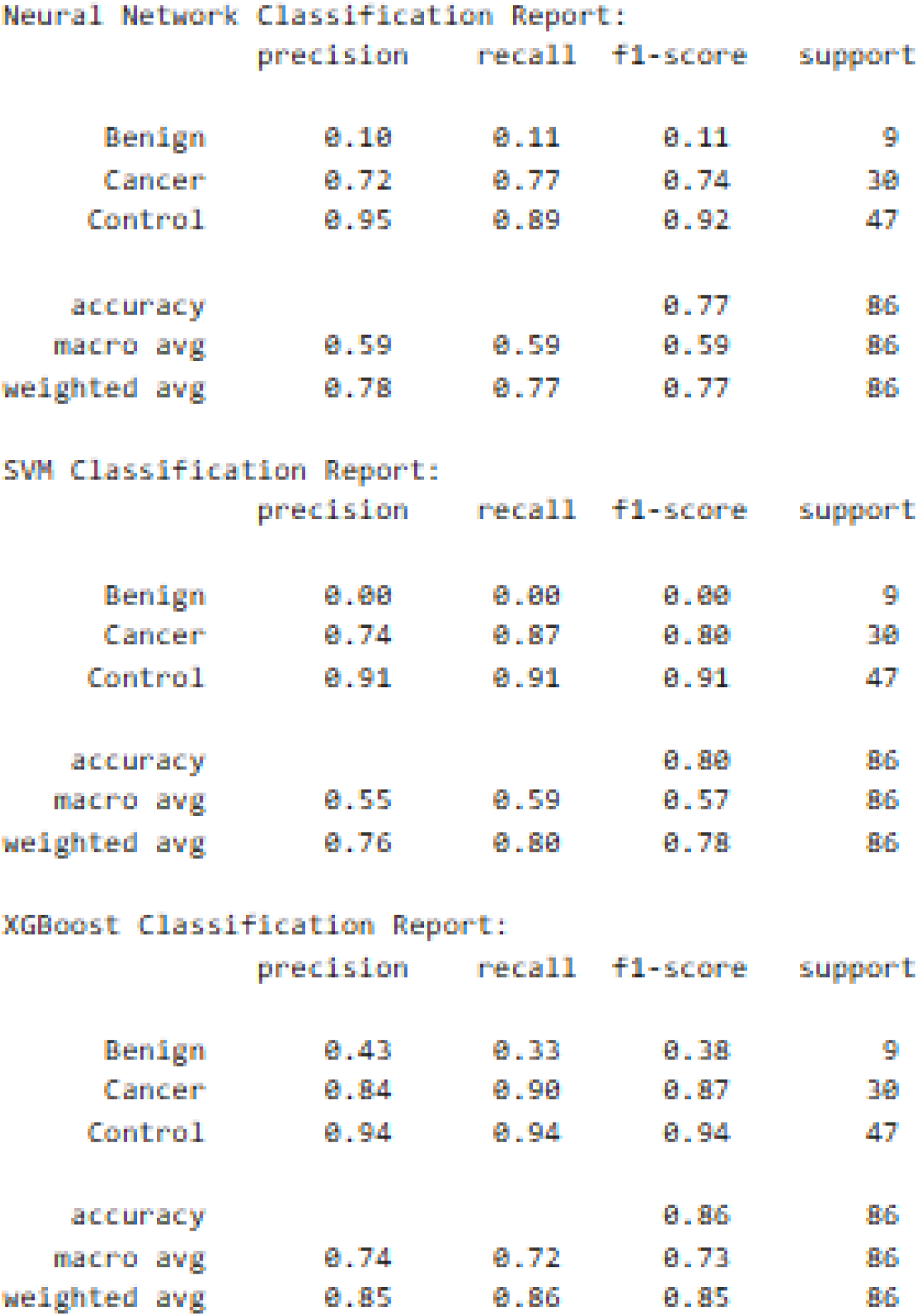
Multiclass Classification Report (80/20).

**Figure 4.1.**
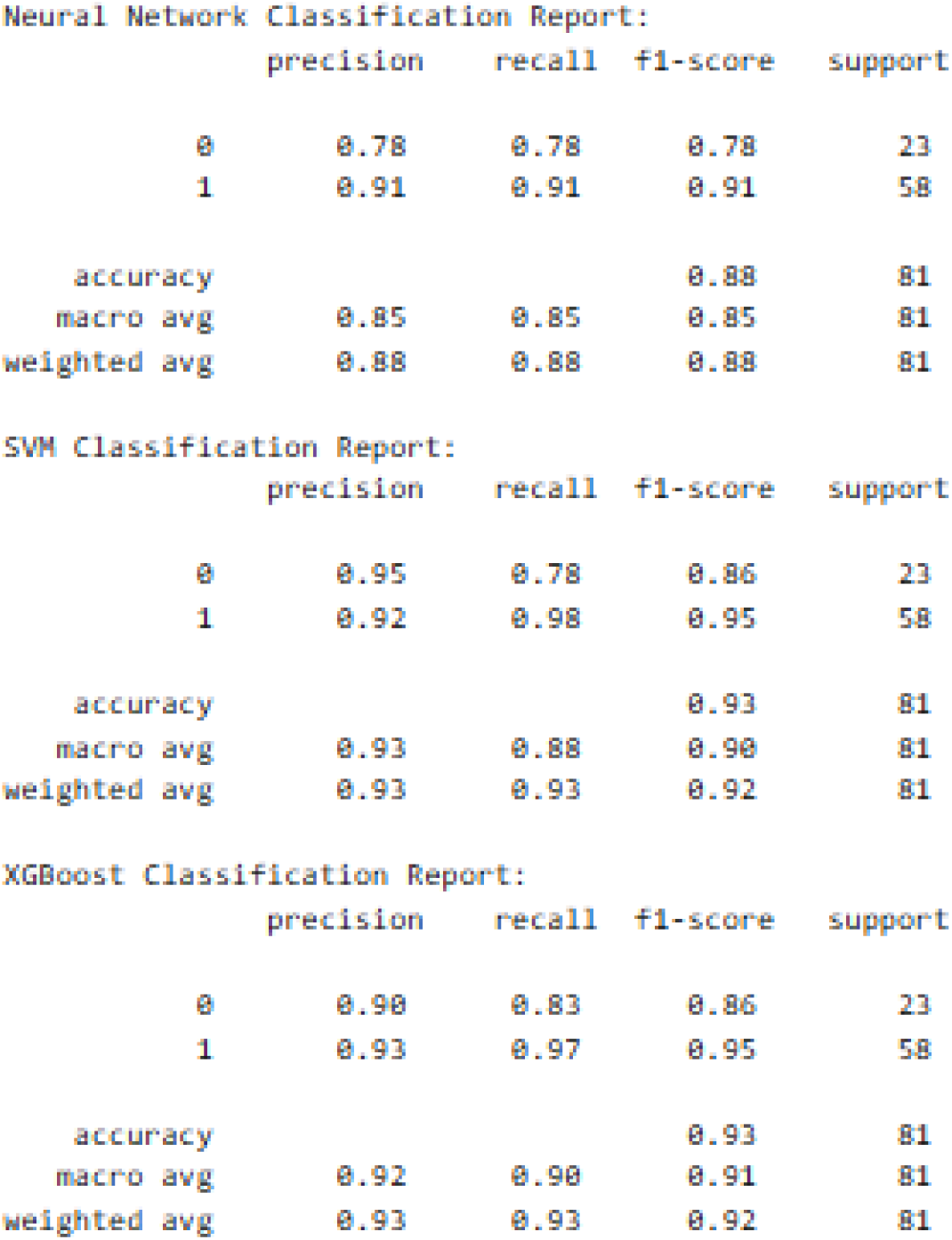
Benign versus Control Classification Report (80/20).

**Figure 4.2.**
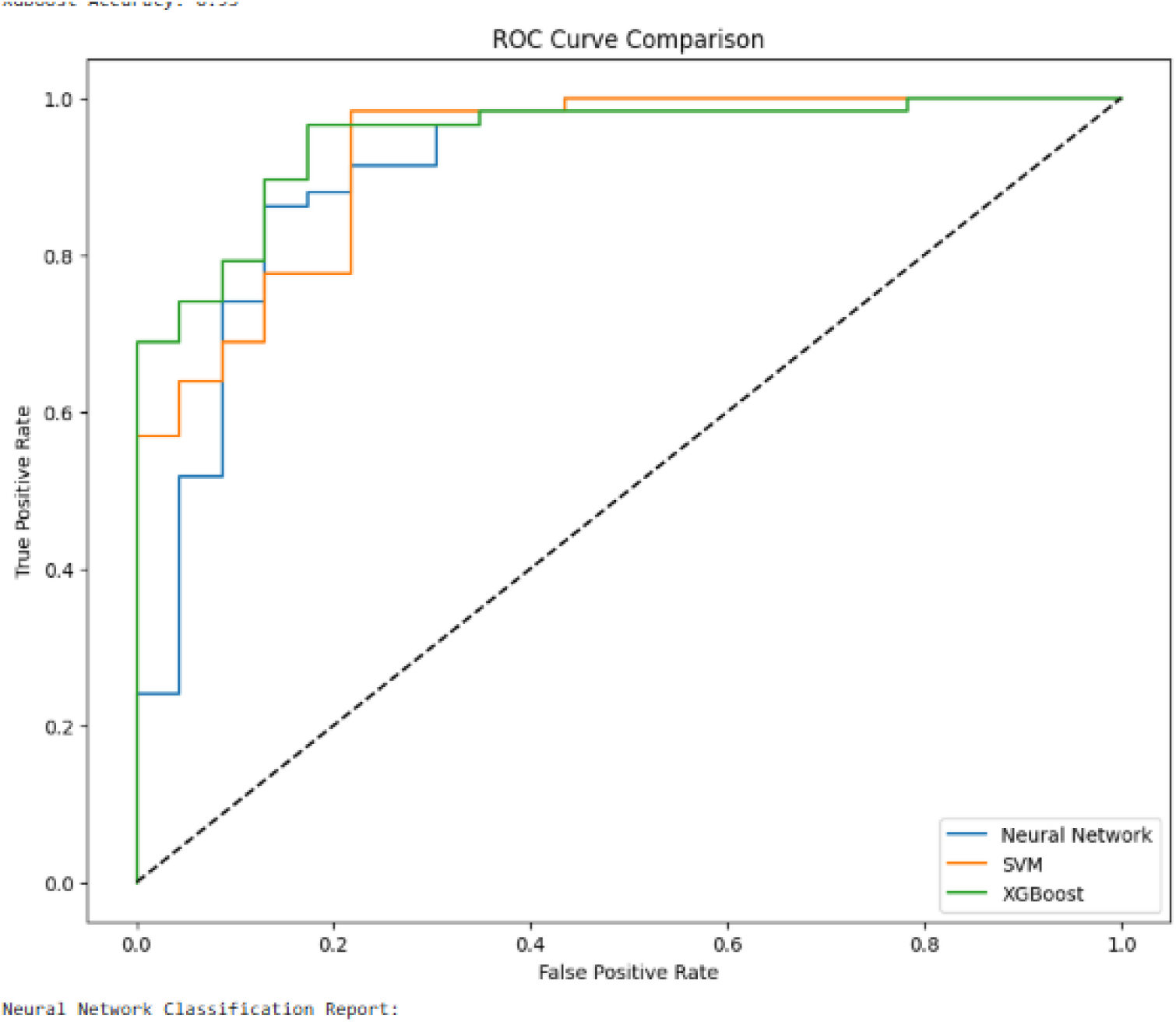
Benign versus Control ROC Curve (80/20).

**Figure 4.3.**
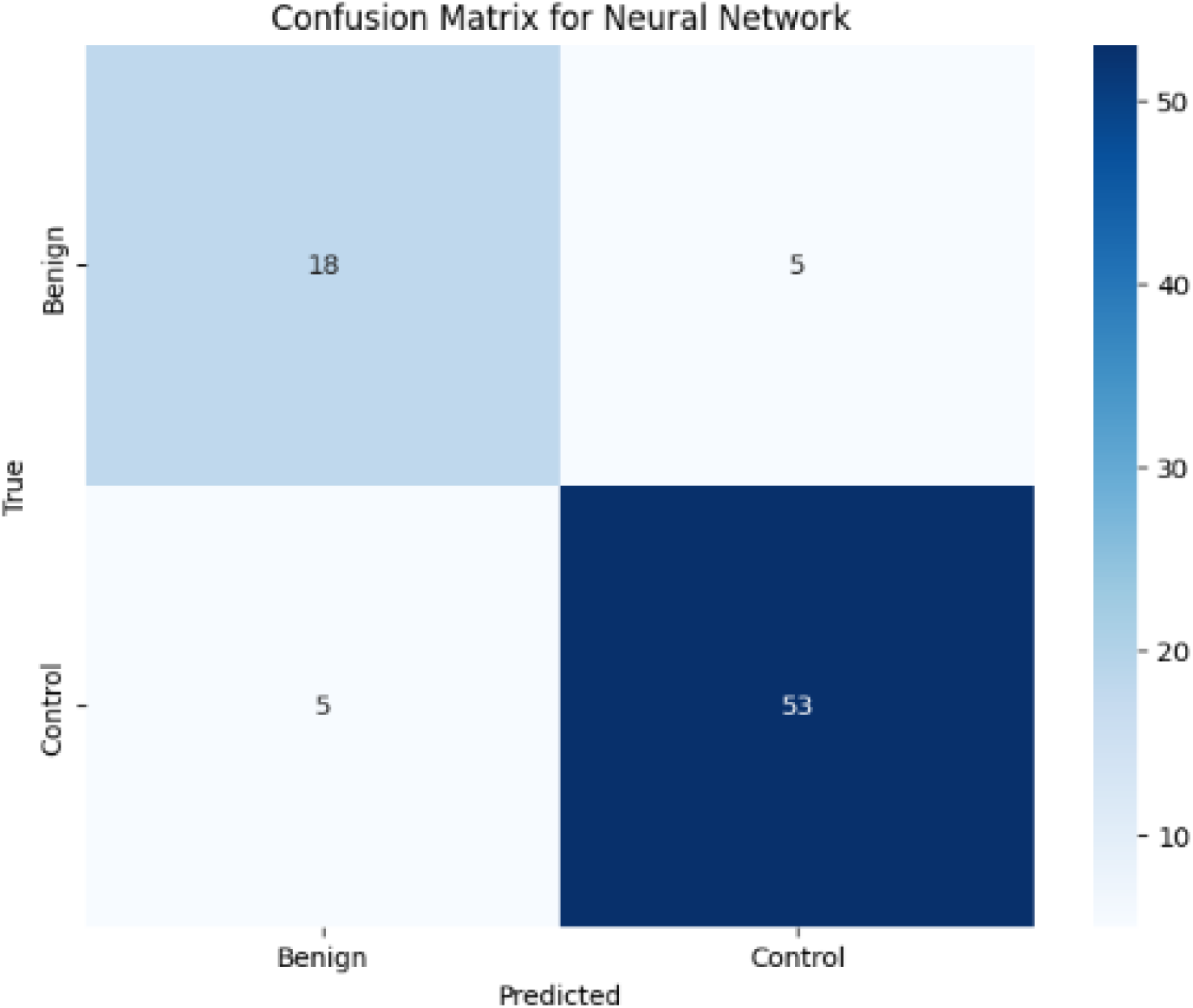
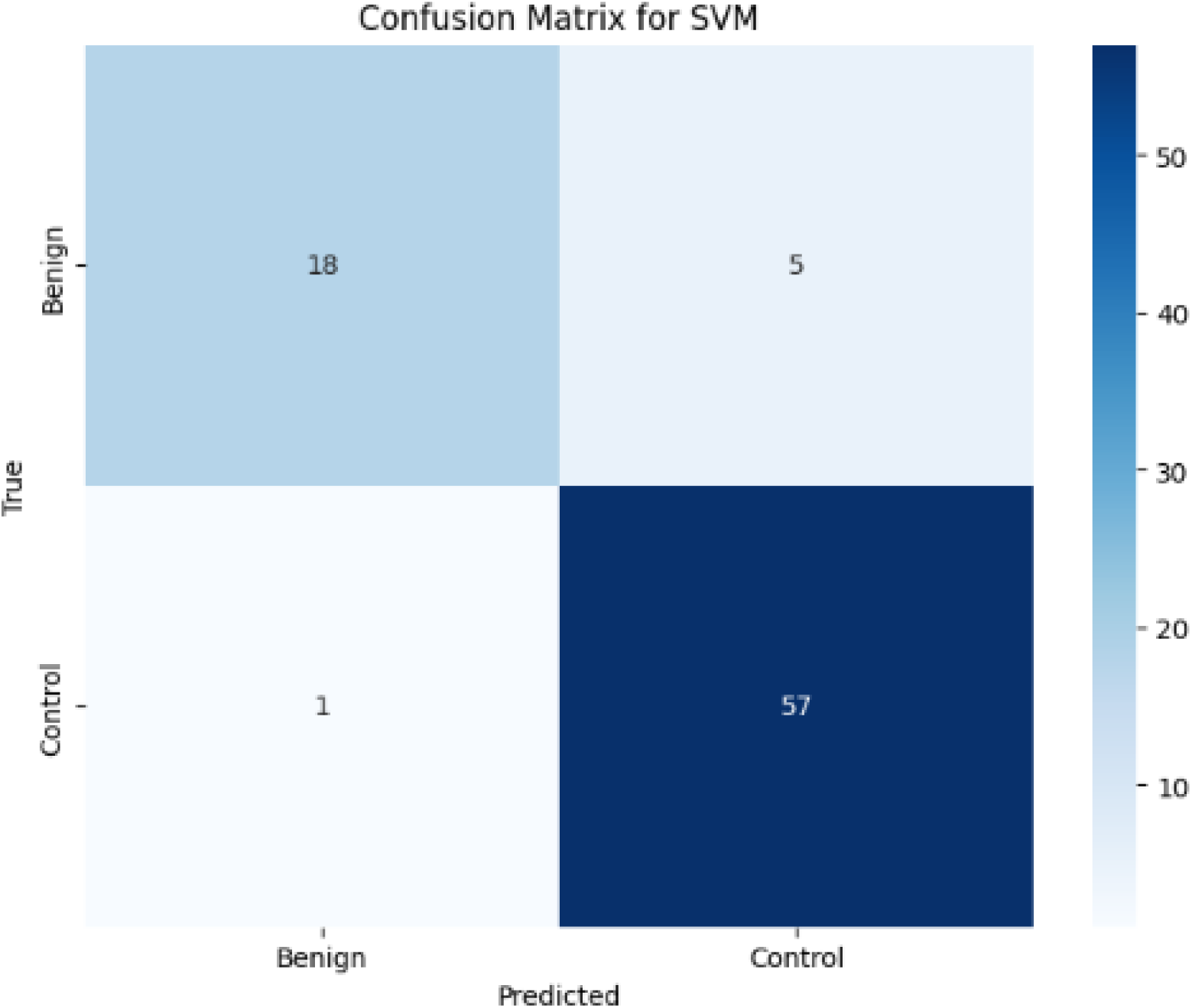
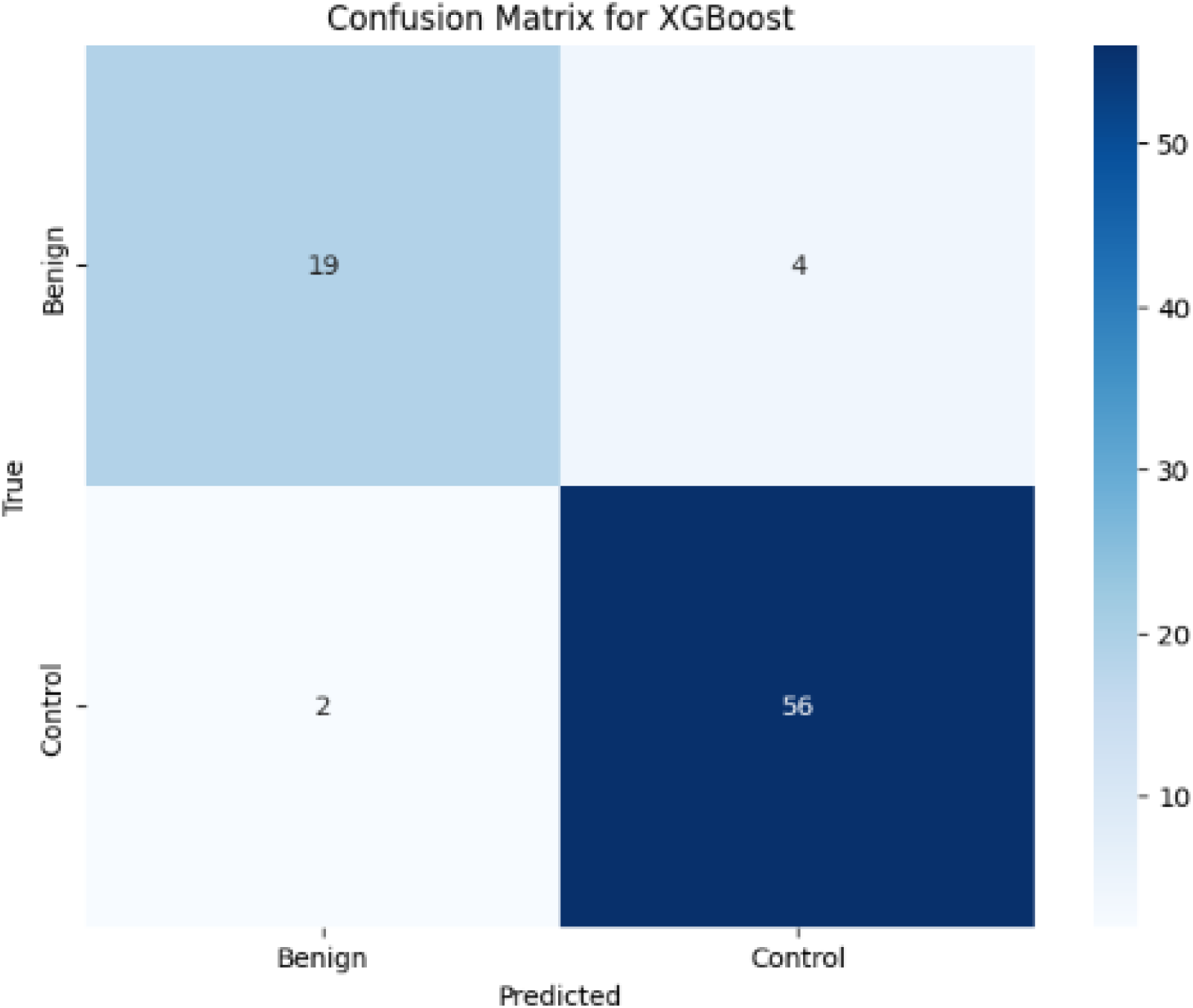
Benign versus Control Confusion Matrix (80/20).

**Figure 5.1.**
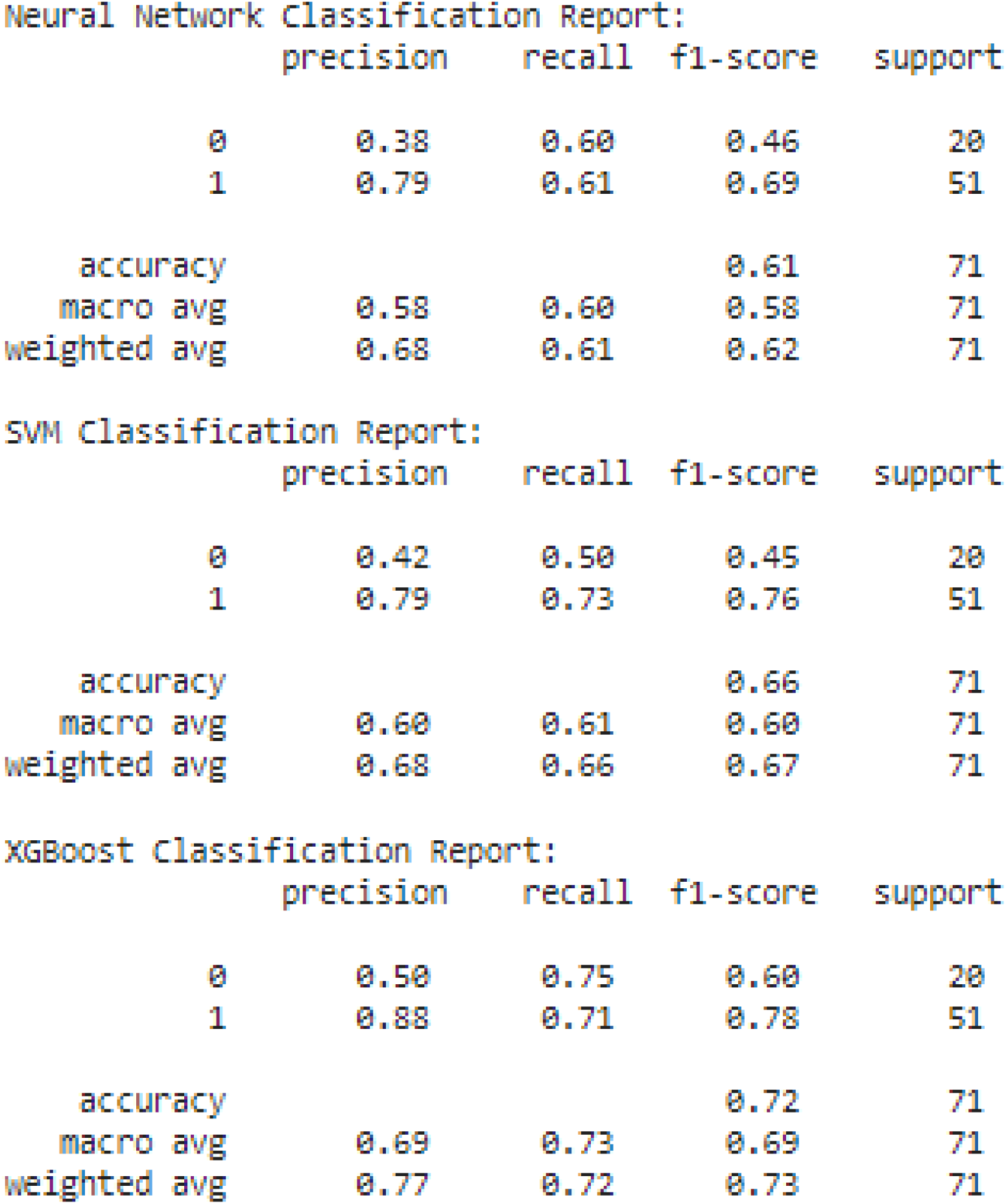
Cancer versus Benign Classification Report (80/20).

**Figure 5.2.**
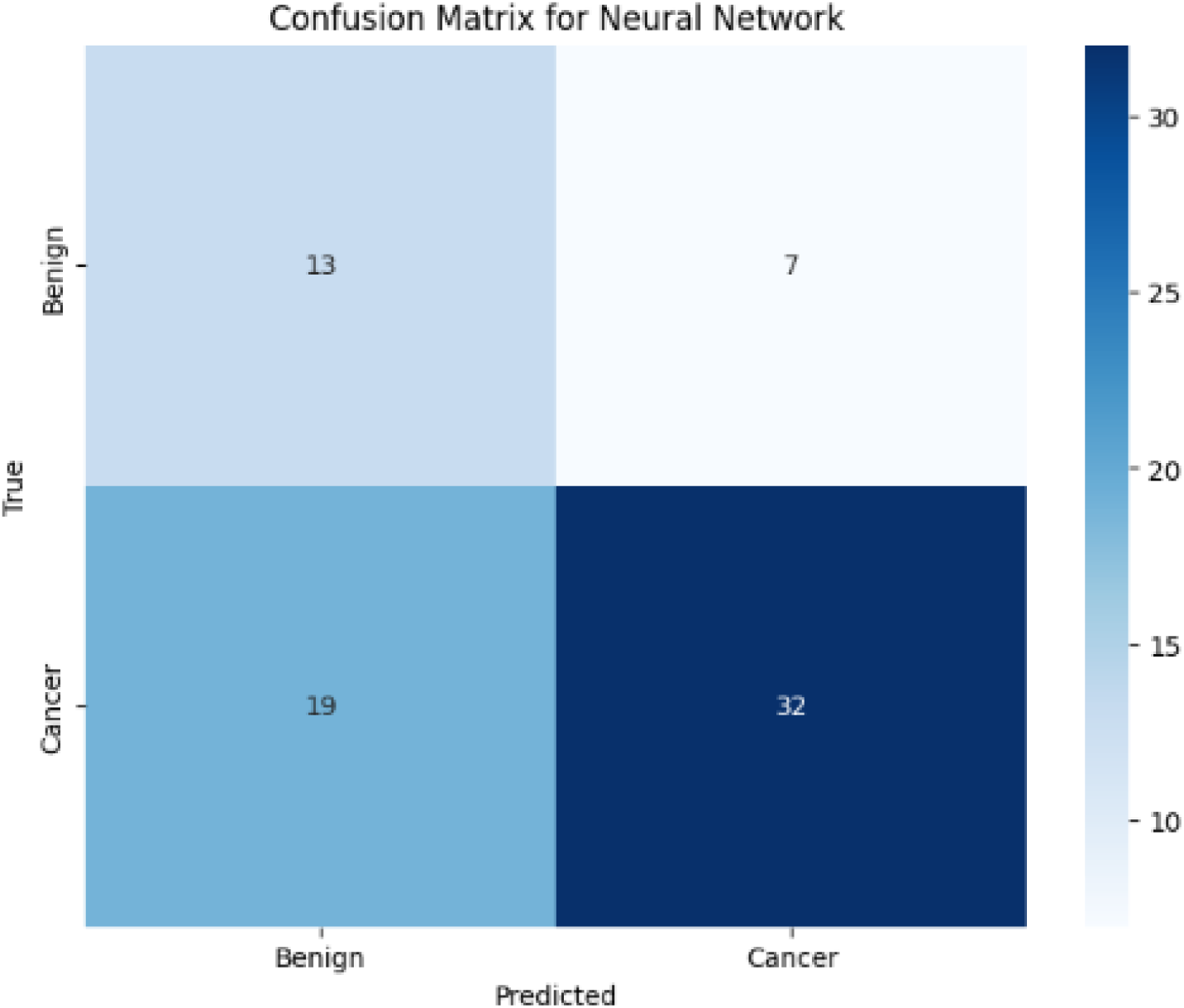
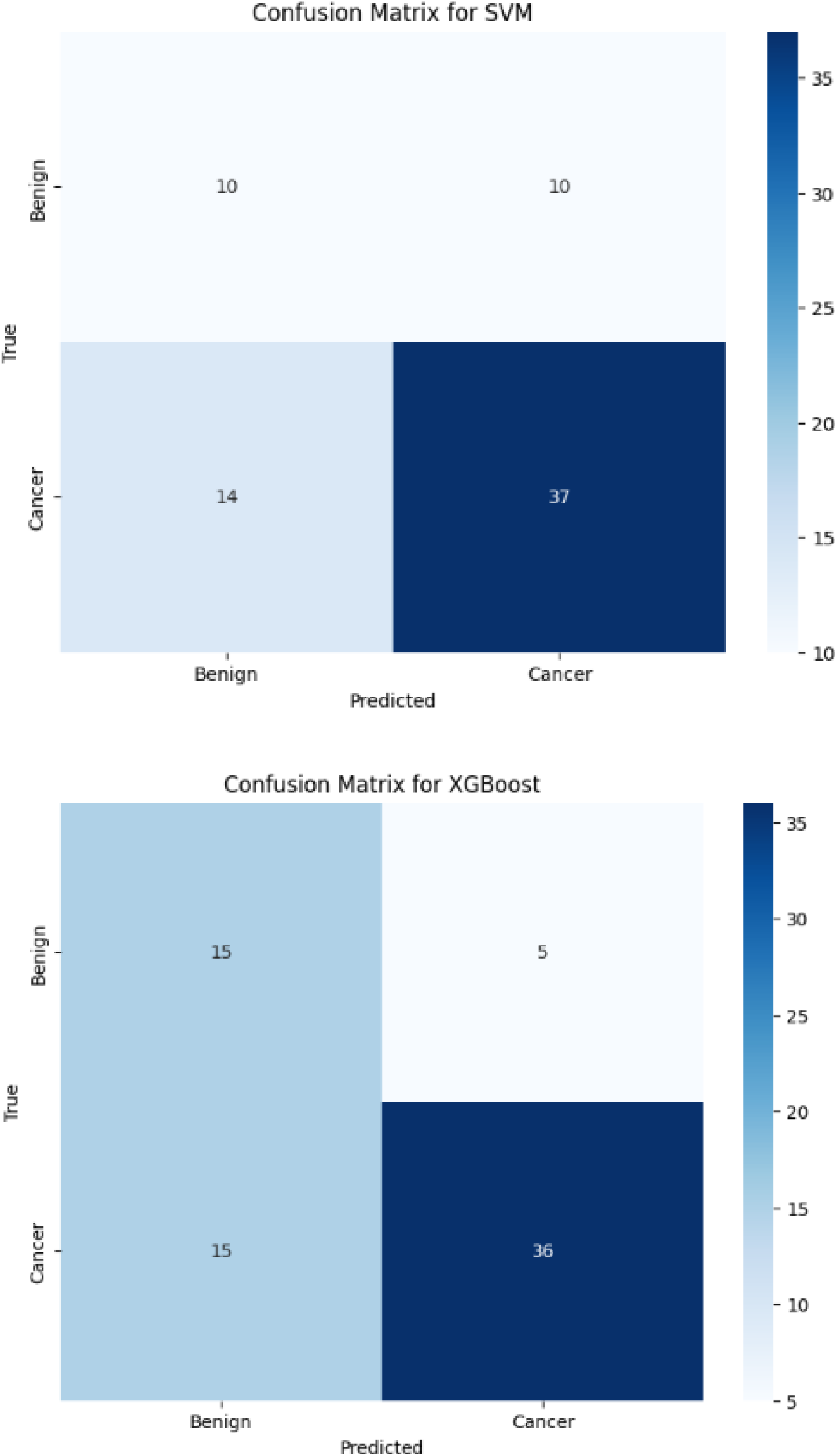
Cancer versus Benign Confusion Matrix (80/20).

**Figure 5.3.**
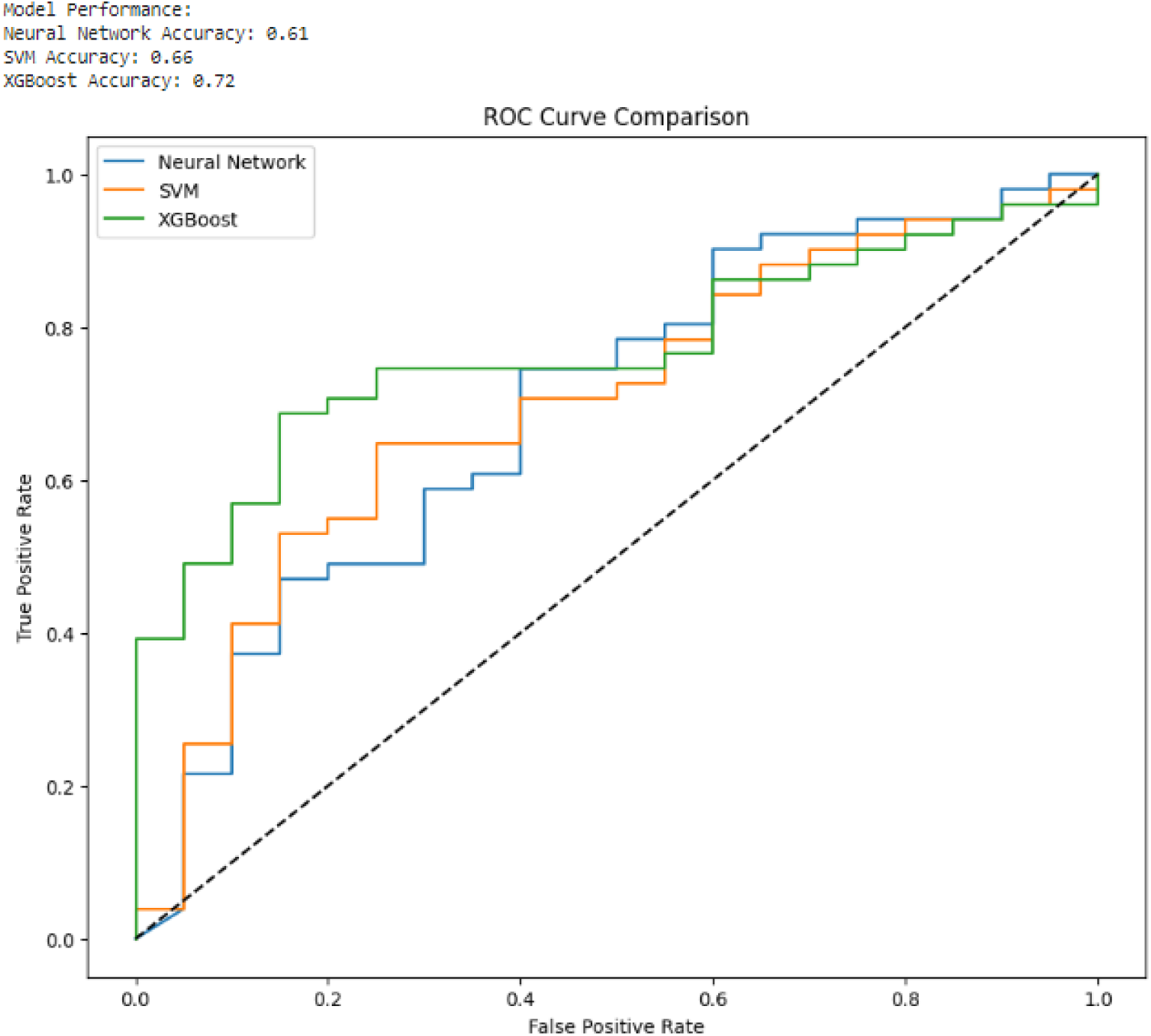
Cancer versus Benign ROC Curve (80/20).

**Figure 5.4.**
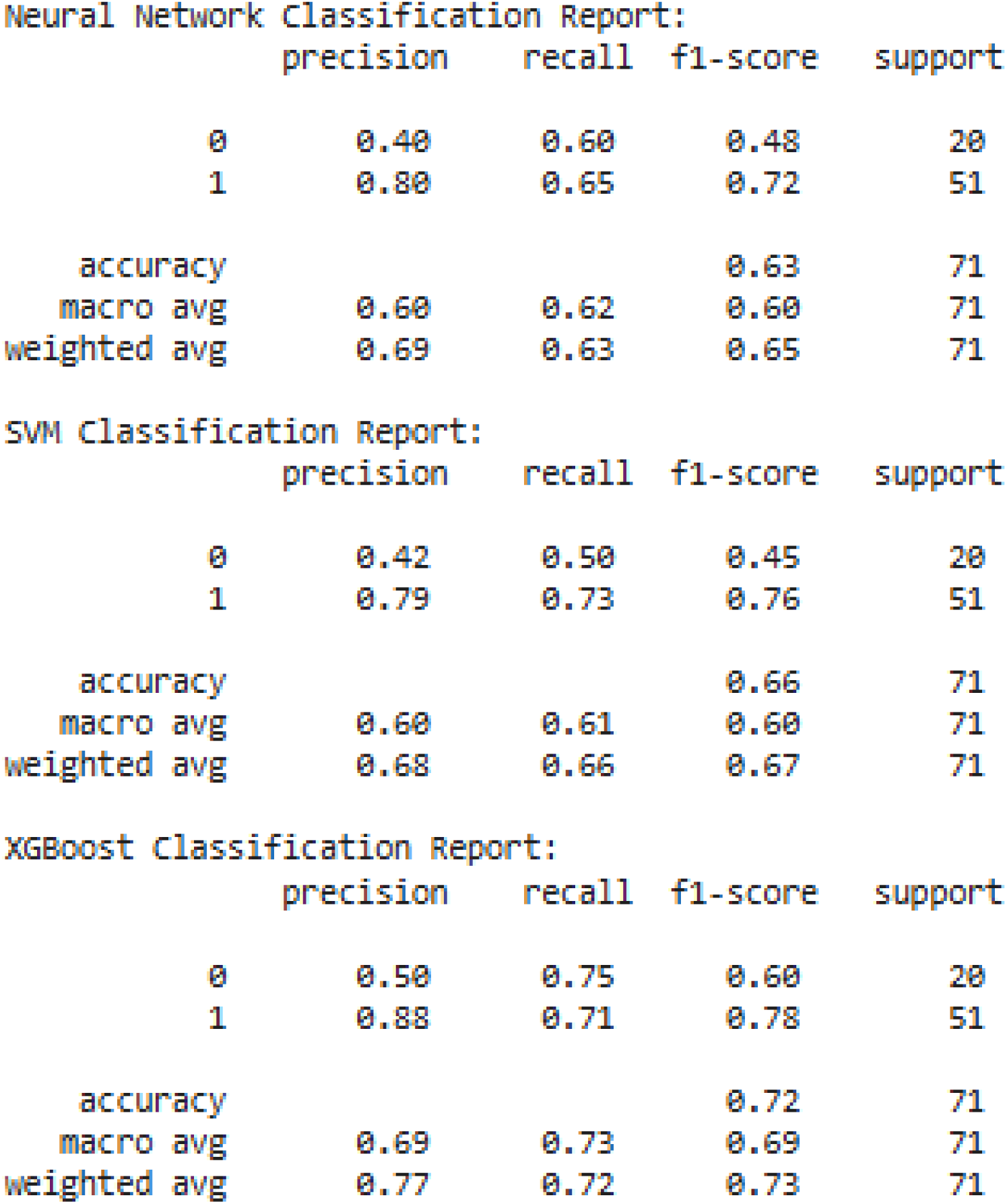
Cancer versus Benign Classification Report (80/20).

**Figure 6.1.**
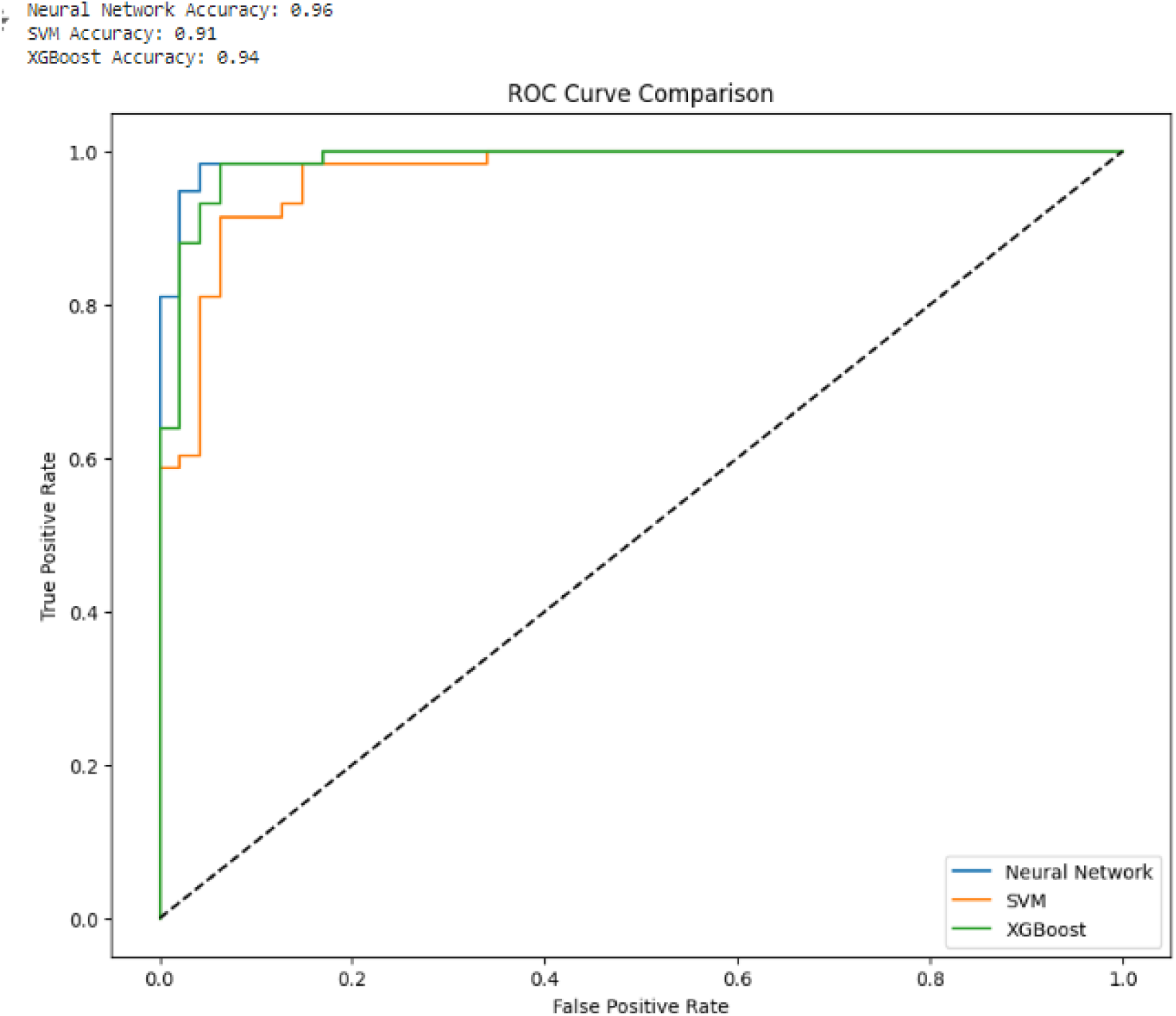
Cancer versus Control ROC Curve (70/30).

**Figure 6.2.**
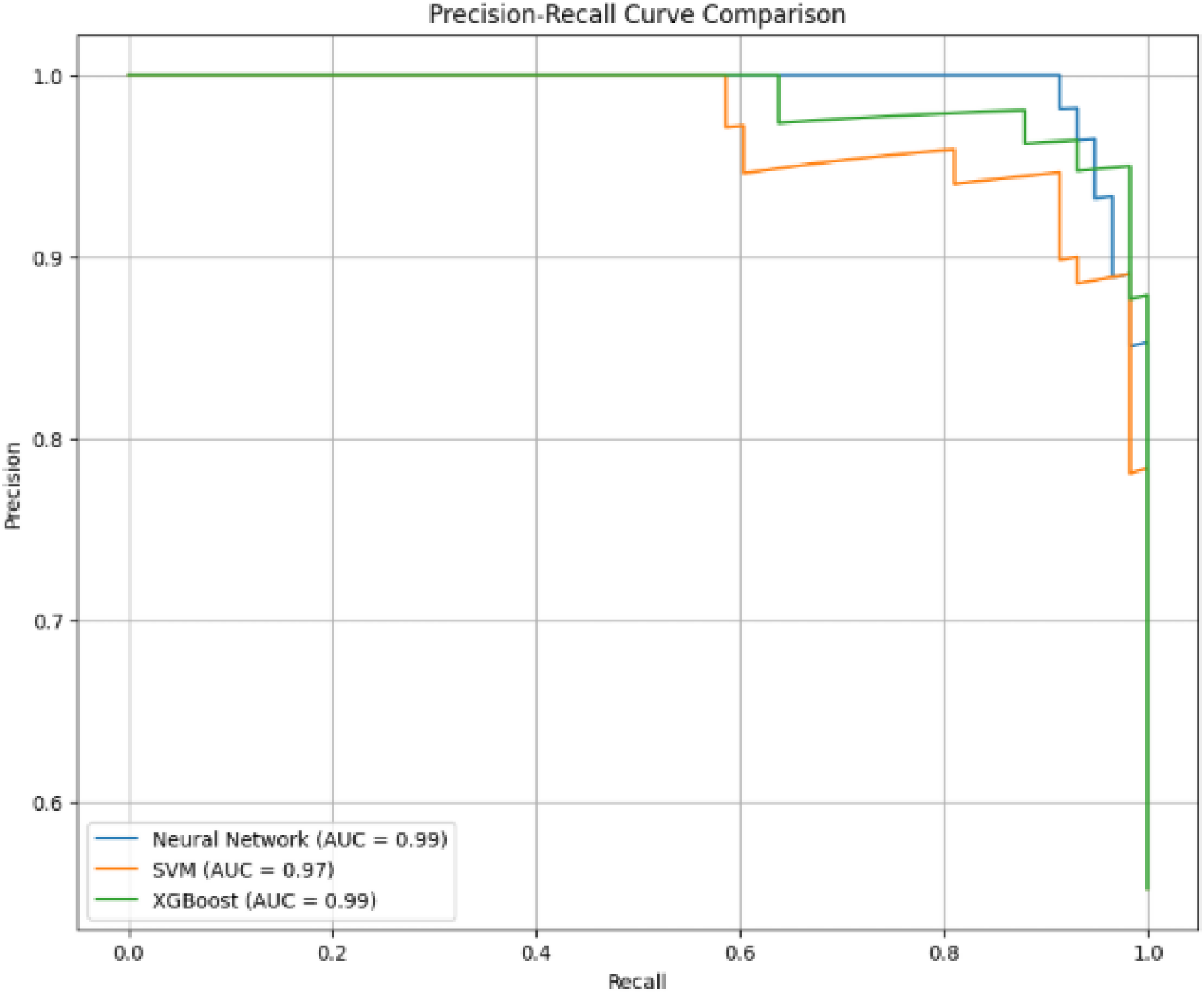
Cancer versus Control PR Curve (70/30).

**Figure 6.3.**
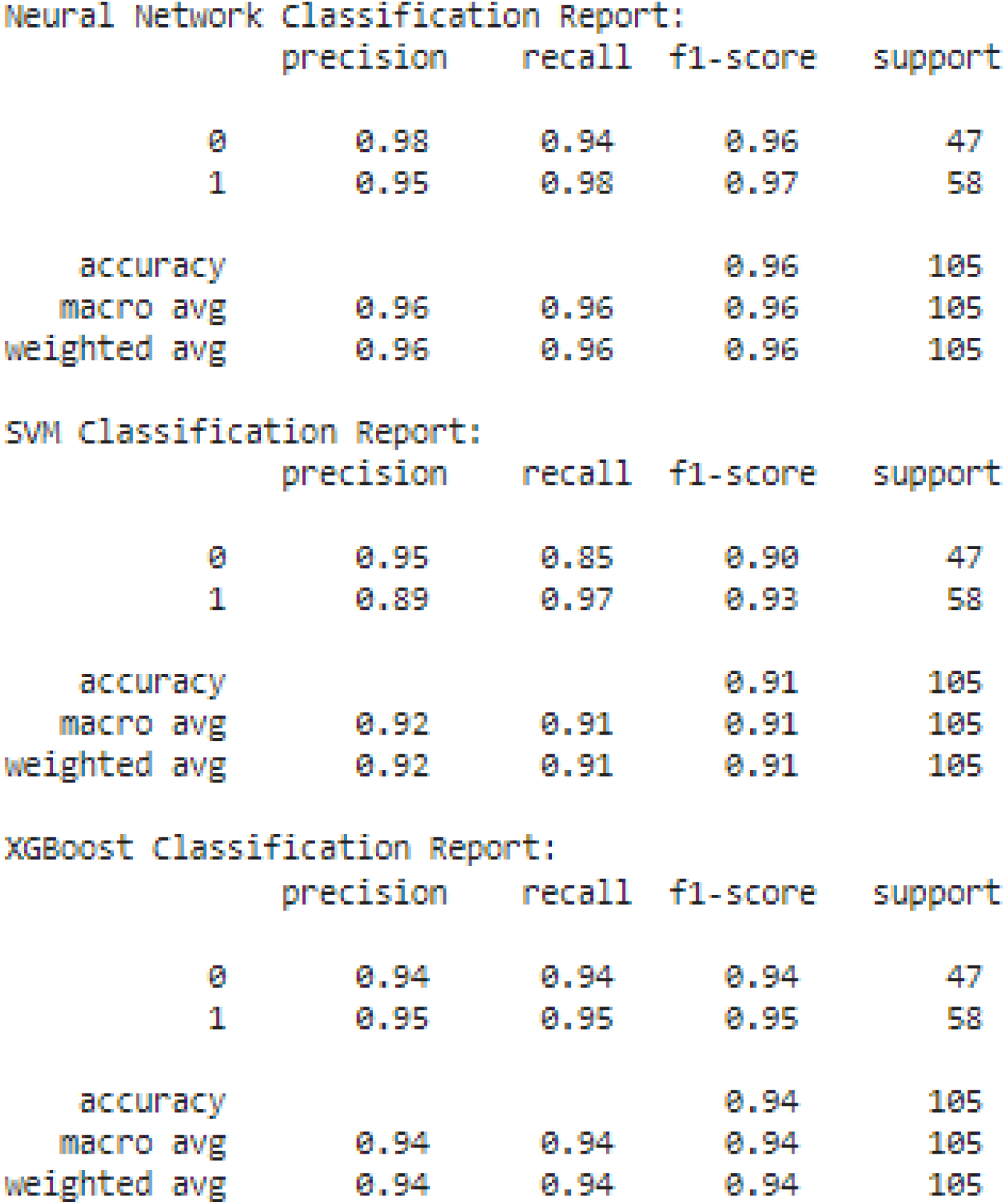
Cancer versus Control Classification Report (70/30).

**Figures 6.4.**
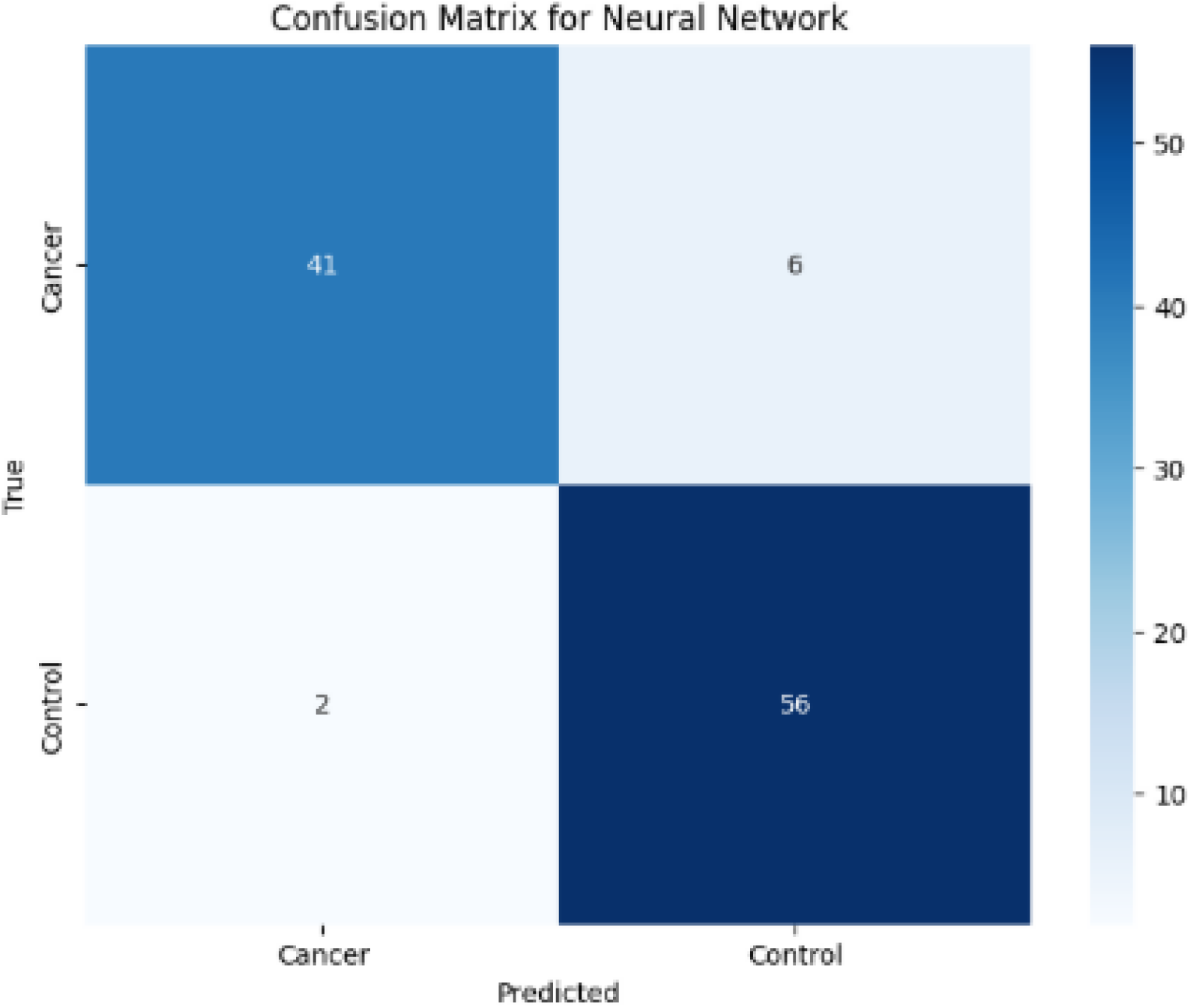
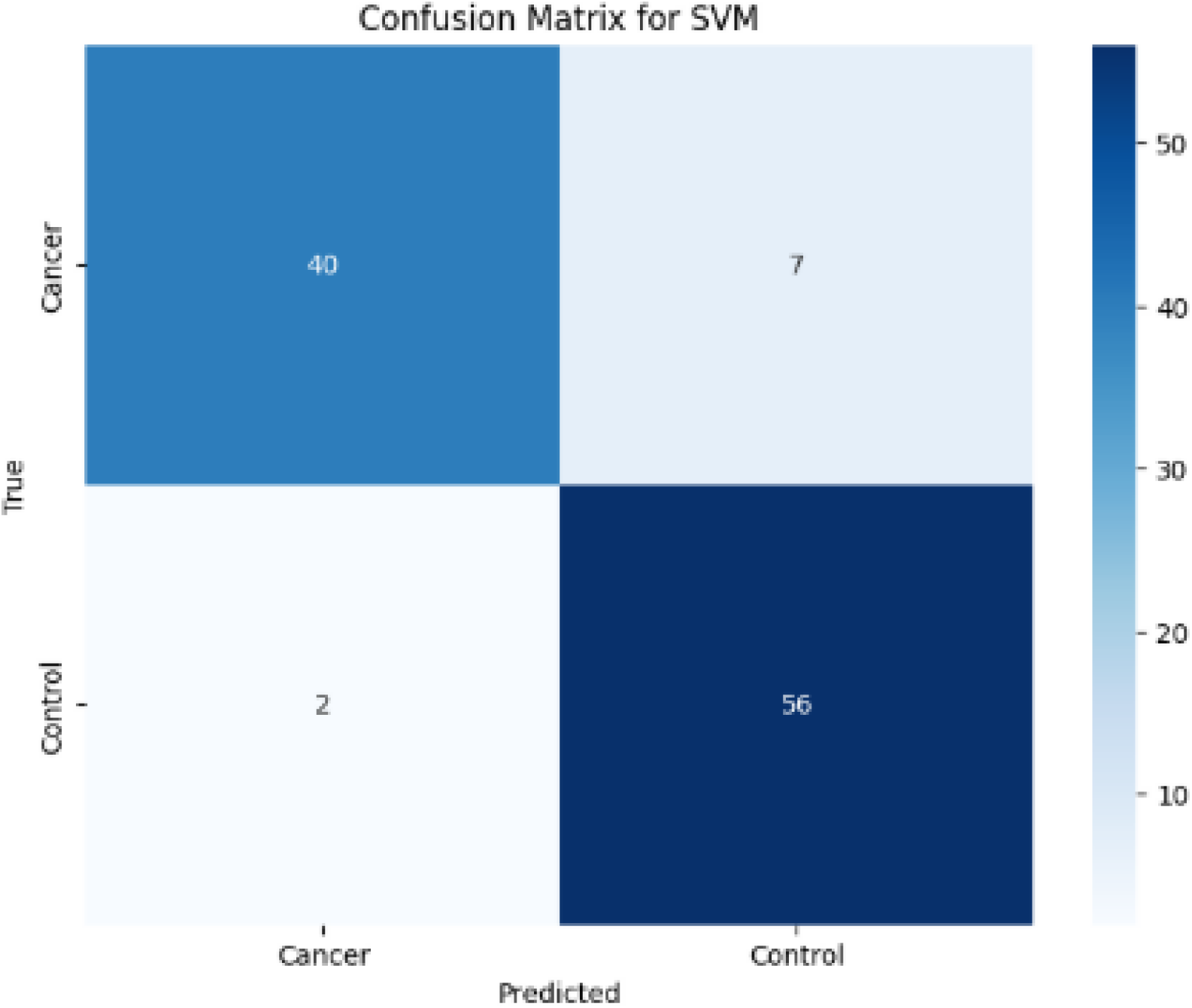
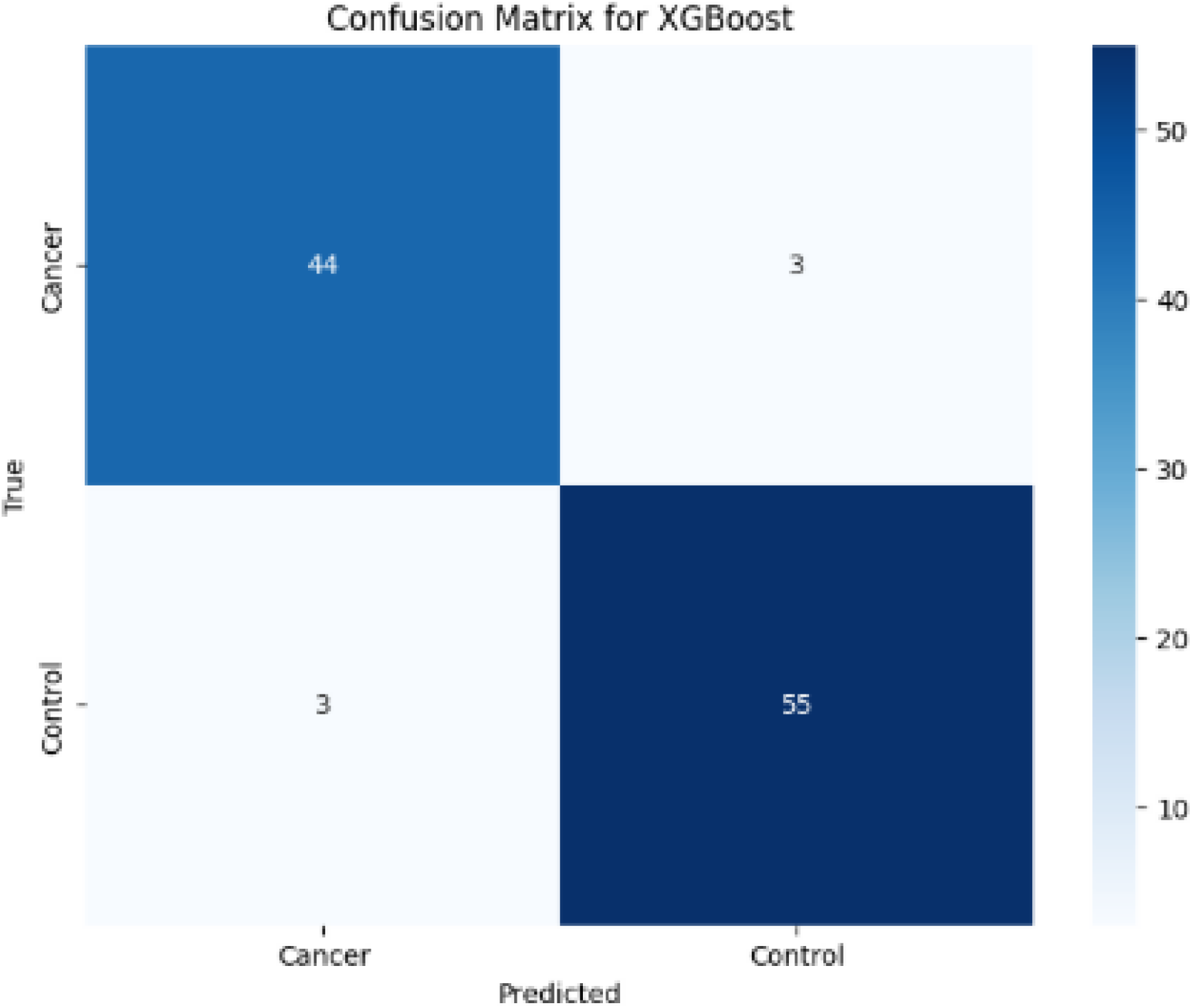
Cancer versus Control Confusion Matrix (70/30).

**Figure 7.1.**
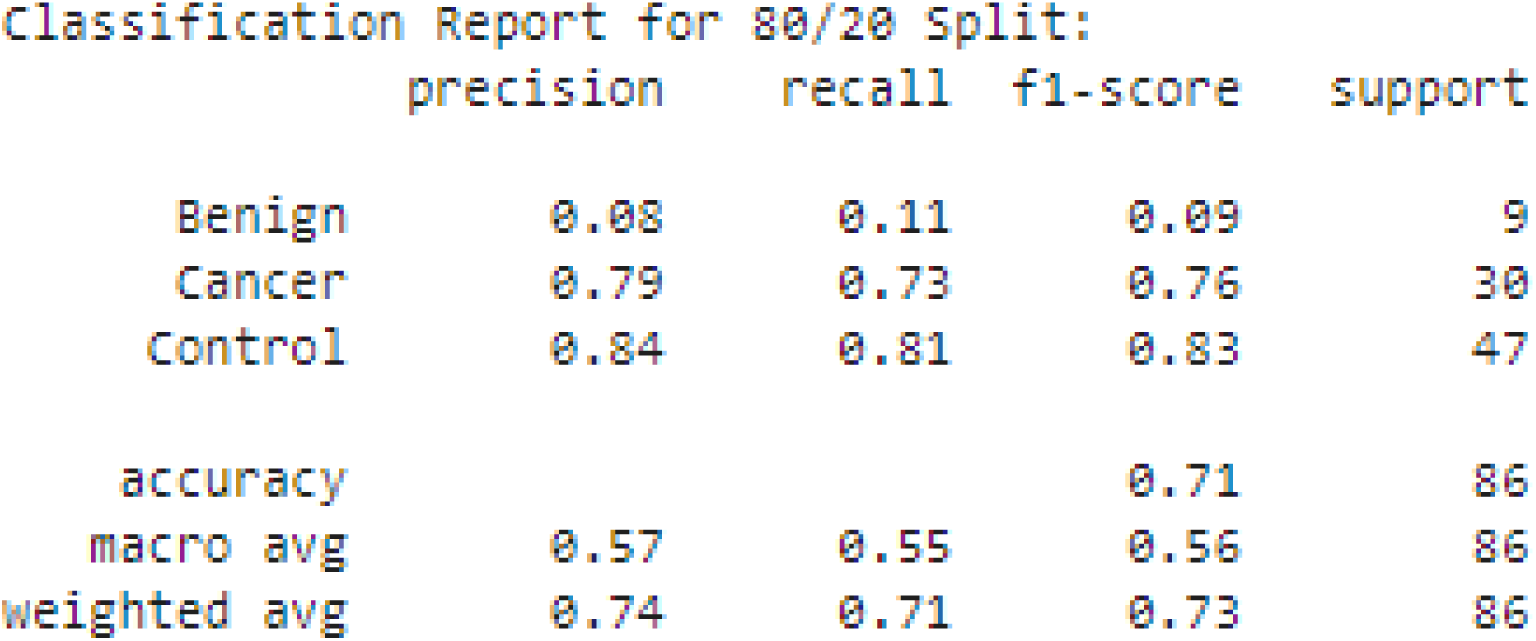
CNN Classification Report (80/20).

**Figure 7.2.**
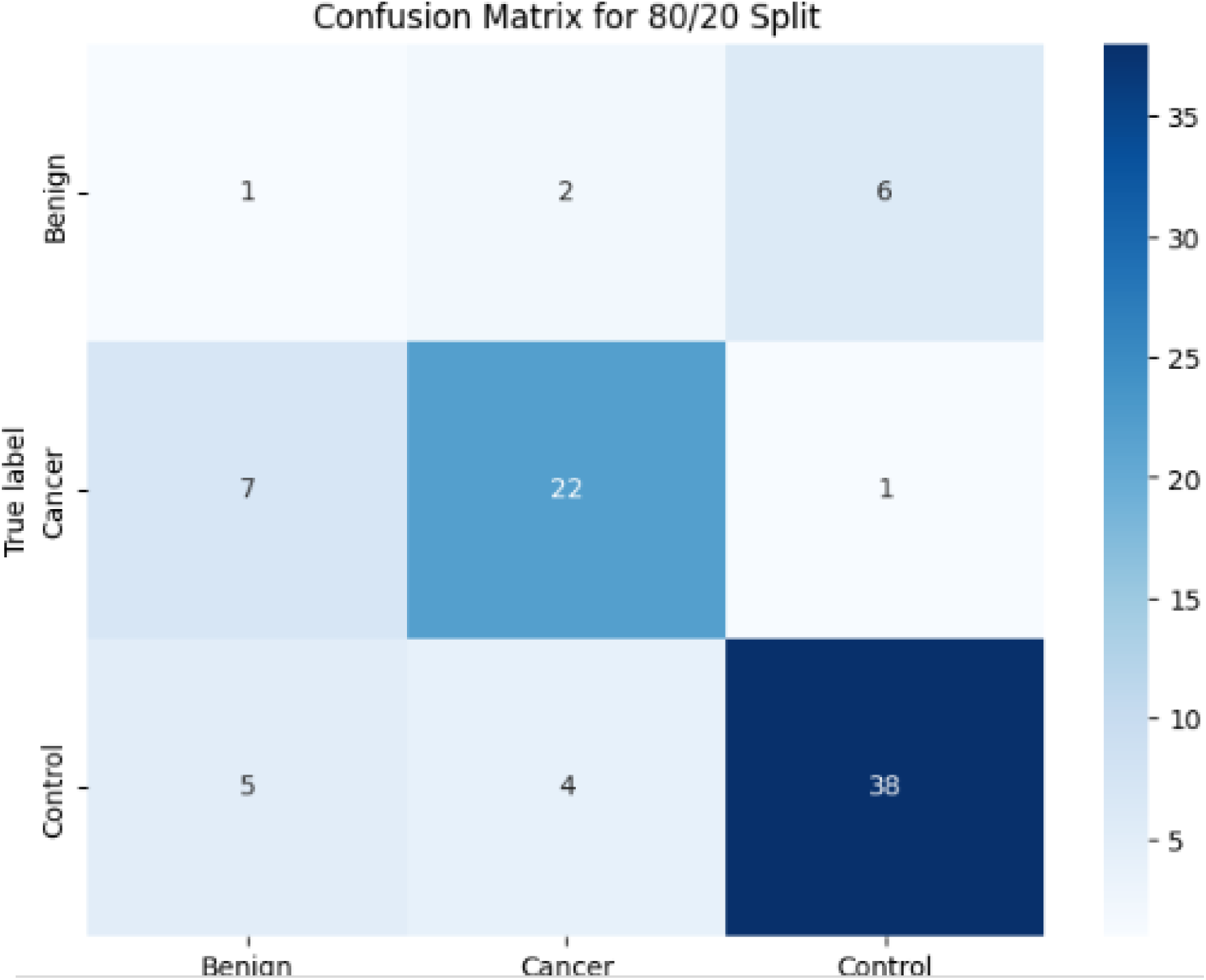
CNN Confusion Matrix (80/20).

**Figure 7.3.**
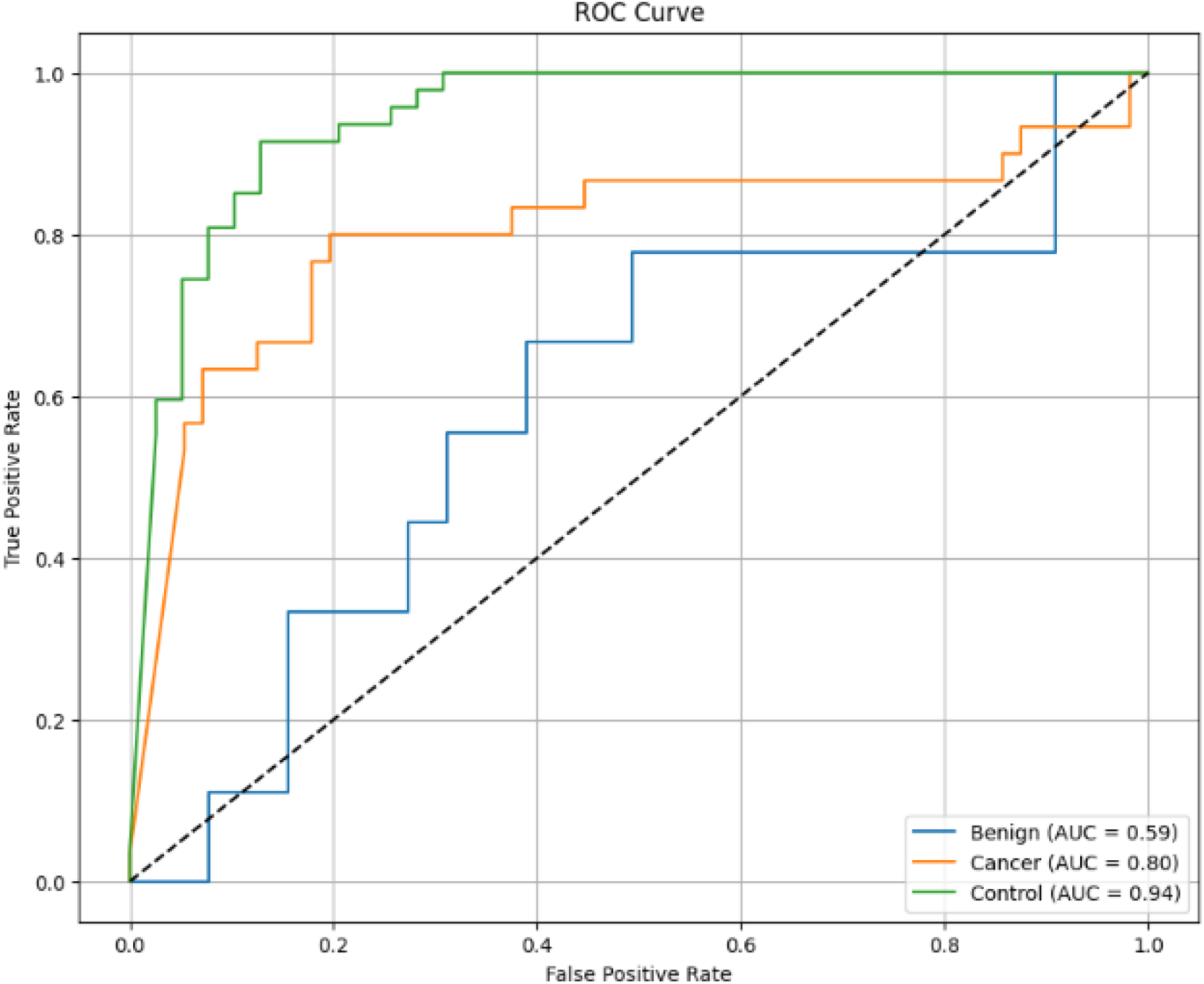
CNN ROC Curve (80/20).

**Figure 7.4.**
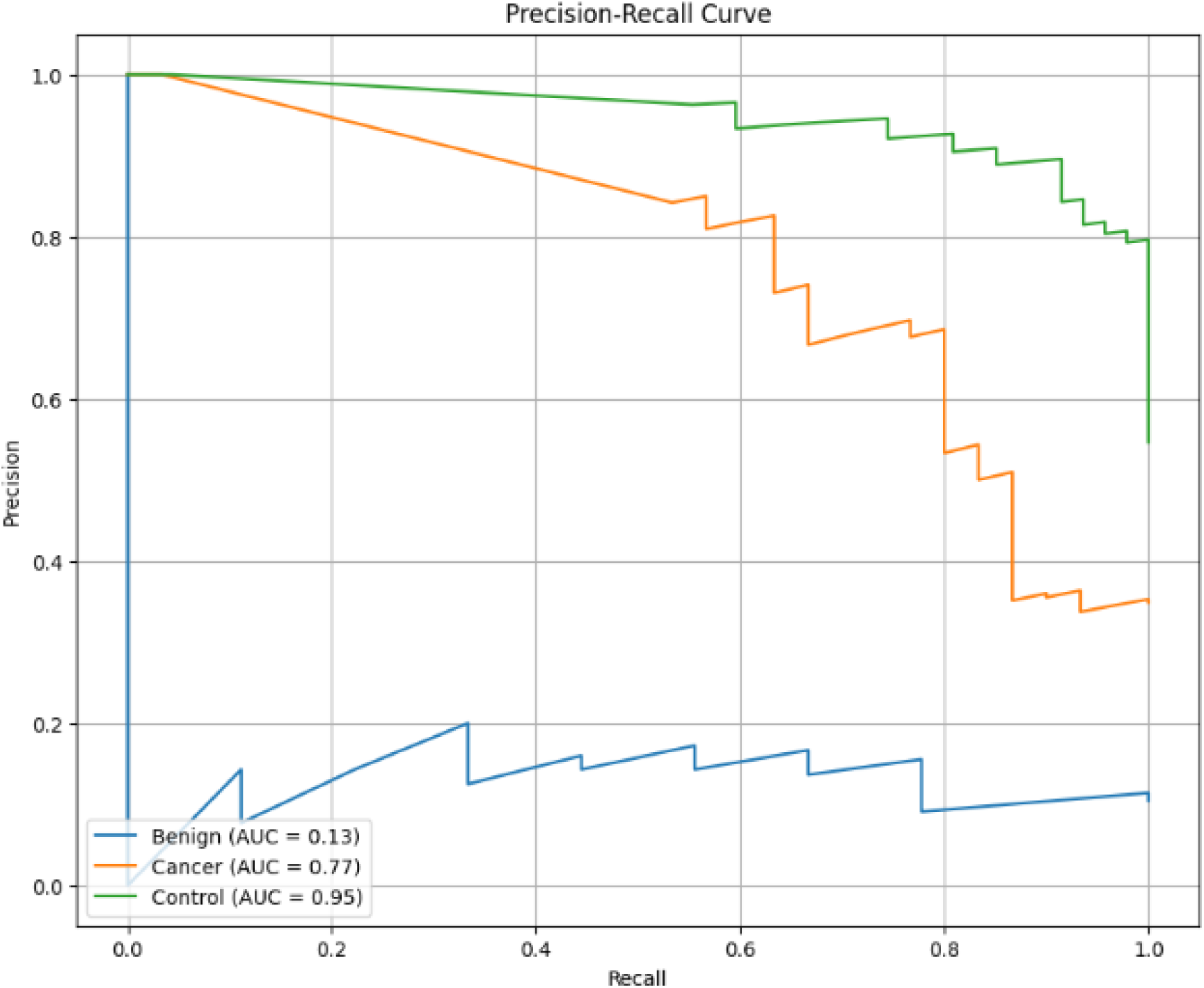
CNN PR Curve (80/20).

**Figure 7.5.**
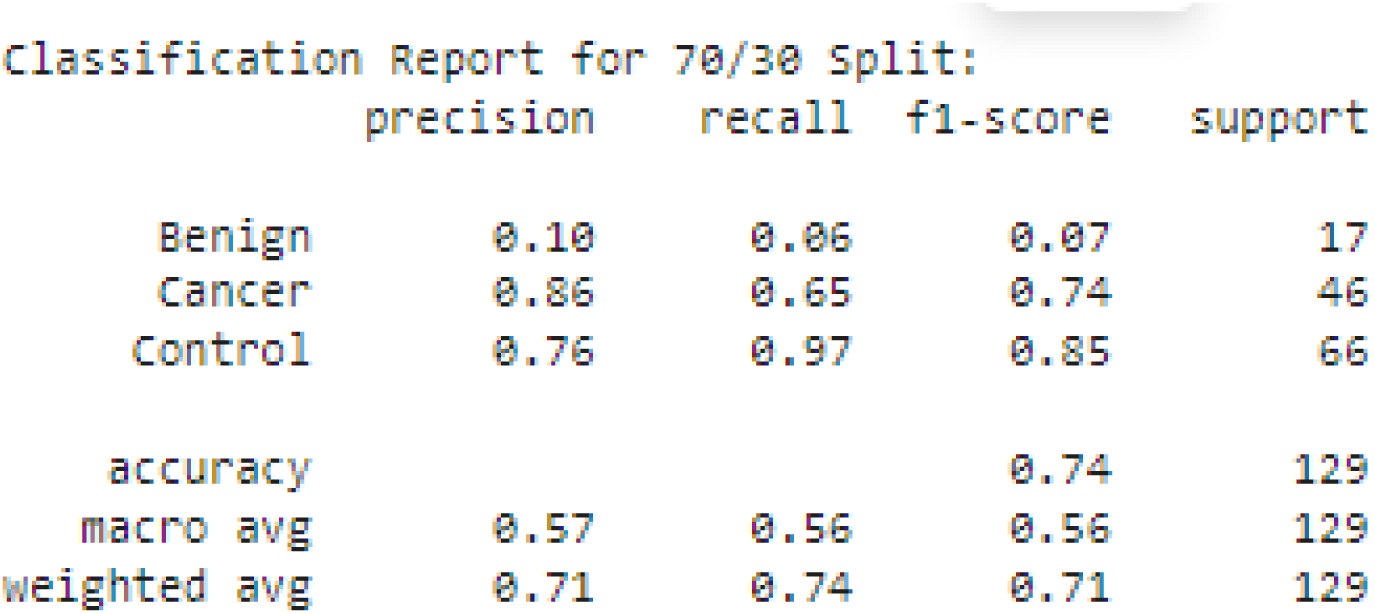
CNN Classification Report (70/30).

**Figure 7.6.**
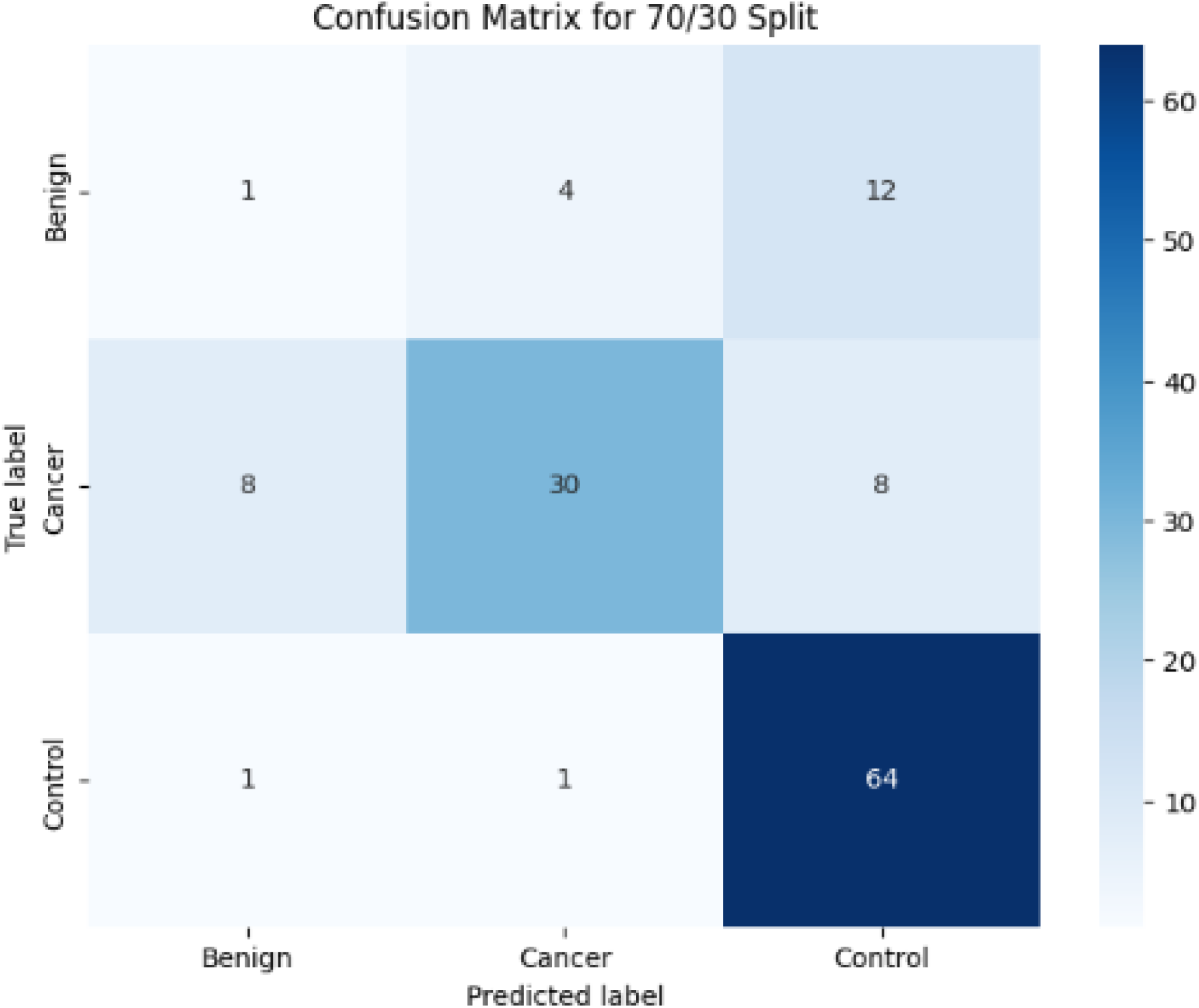
CNN Confusion Matrix (70/30).

**Figure 7.7.**
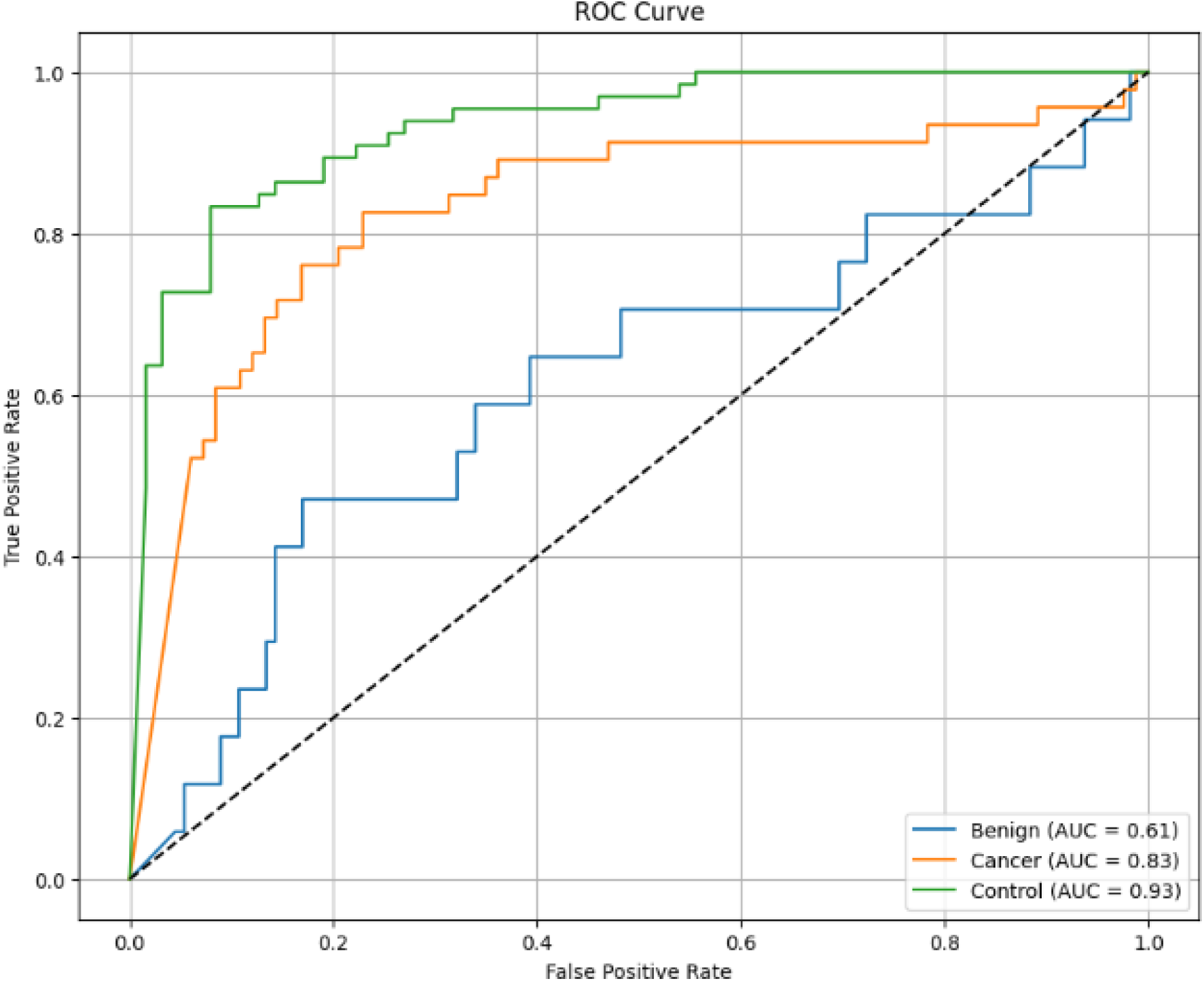
CNN ROC Curve (70/30).

**Figure 7.8.**
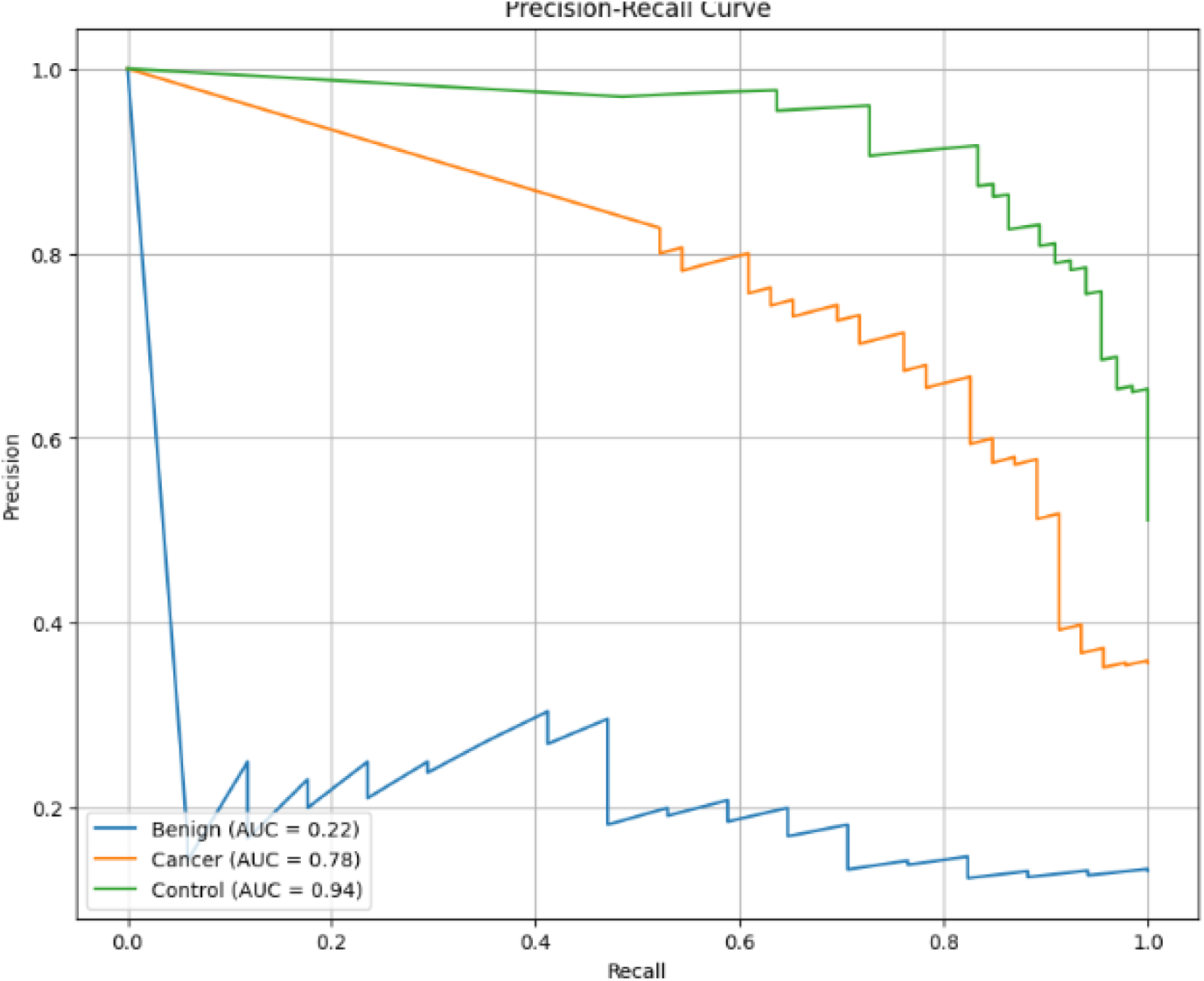
CNN PR Curve (70/30).

**Figure 7.9.**
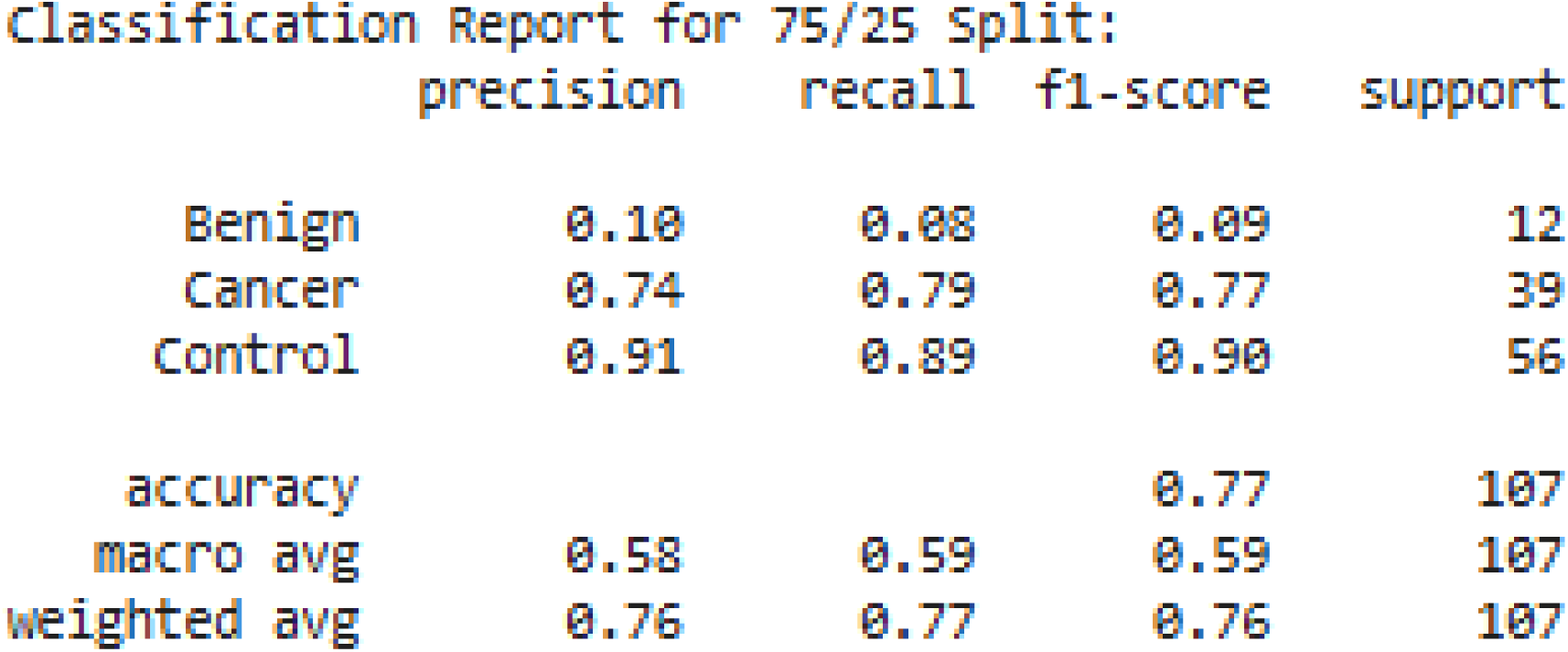
CNN Classification Report (75/25).

**Figure 7.10.**
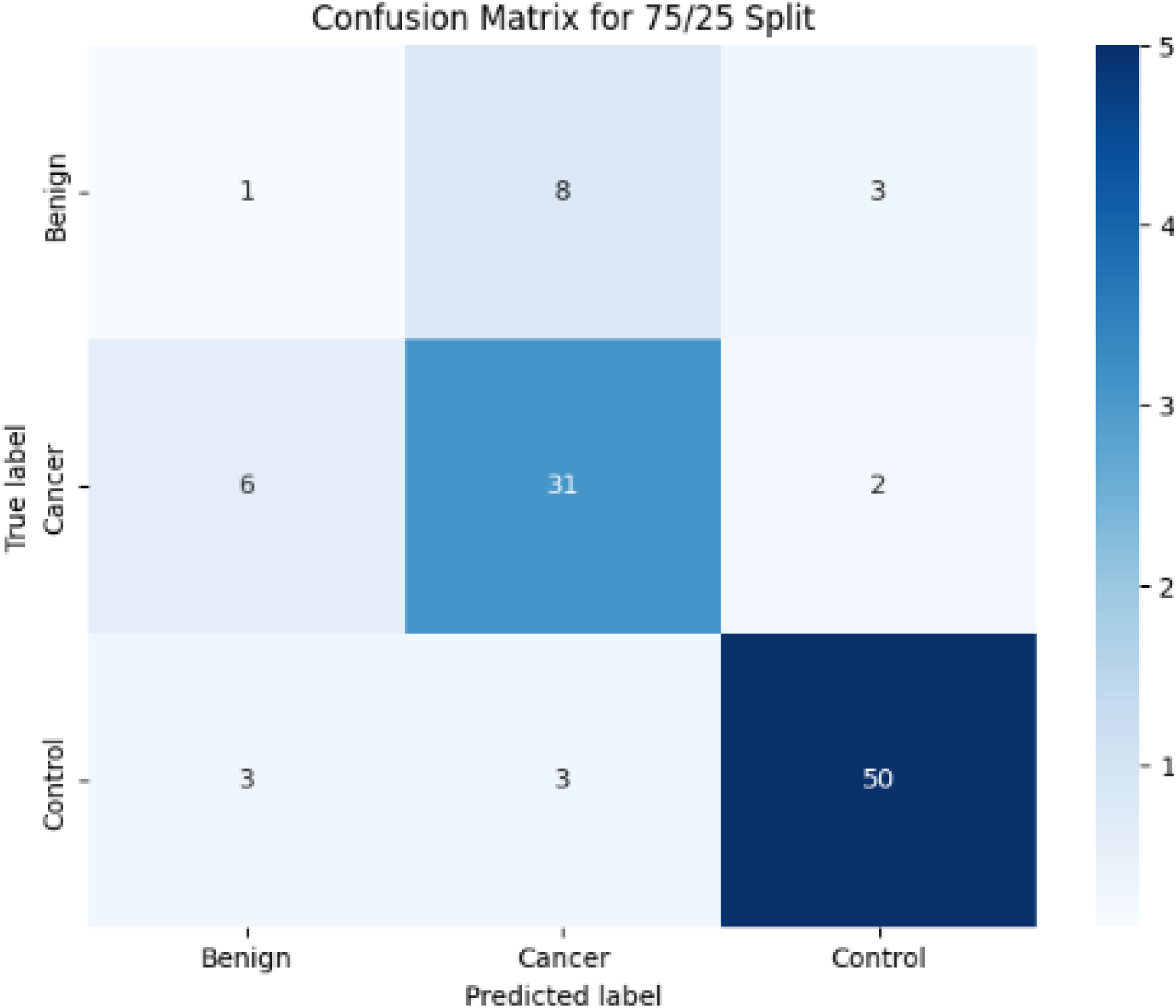
CNN Confusion Matrix (75/25).

**Figure 7.11.**
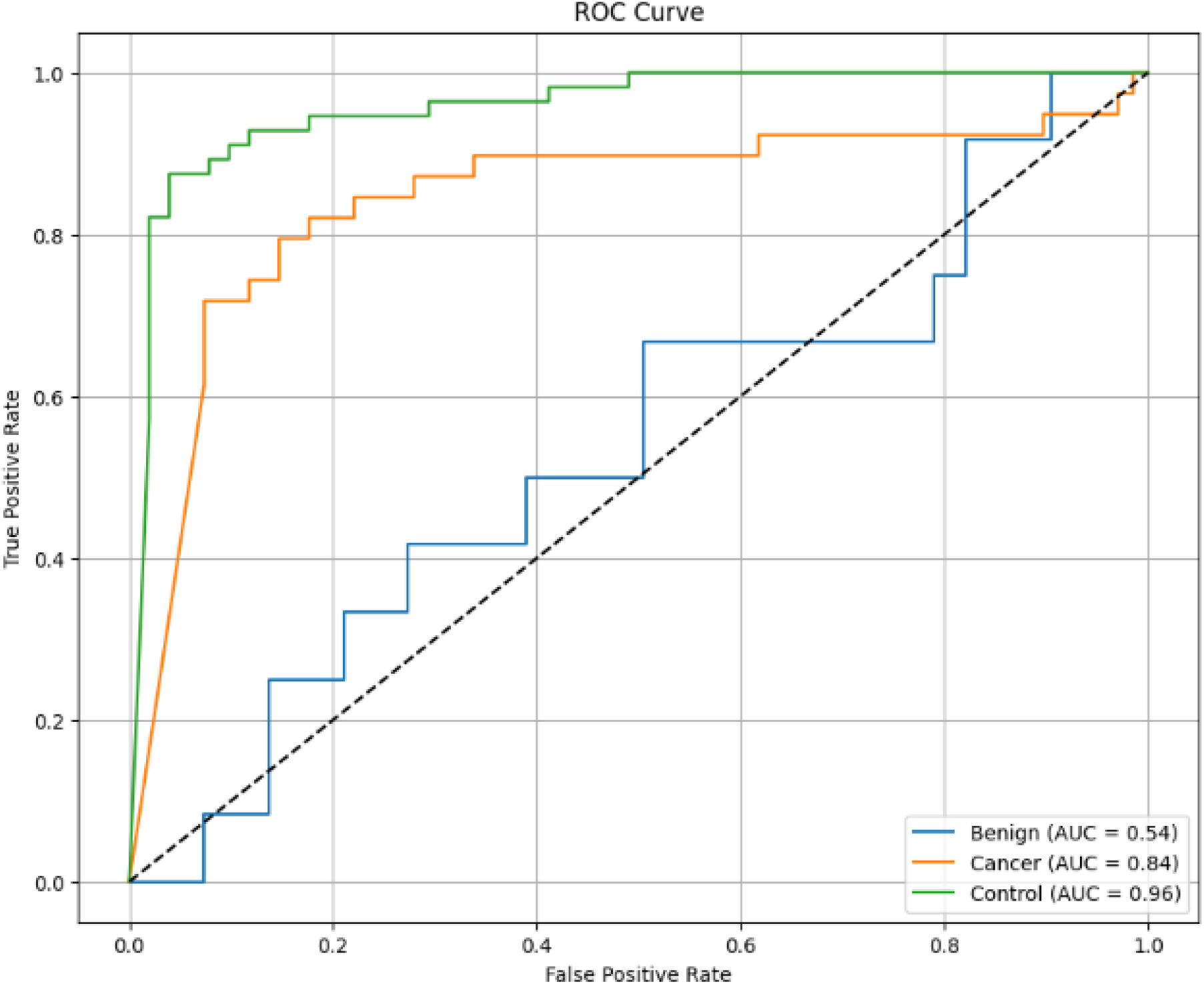
CNN ROC Curve (75/25).

**Figure 7.12.**
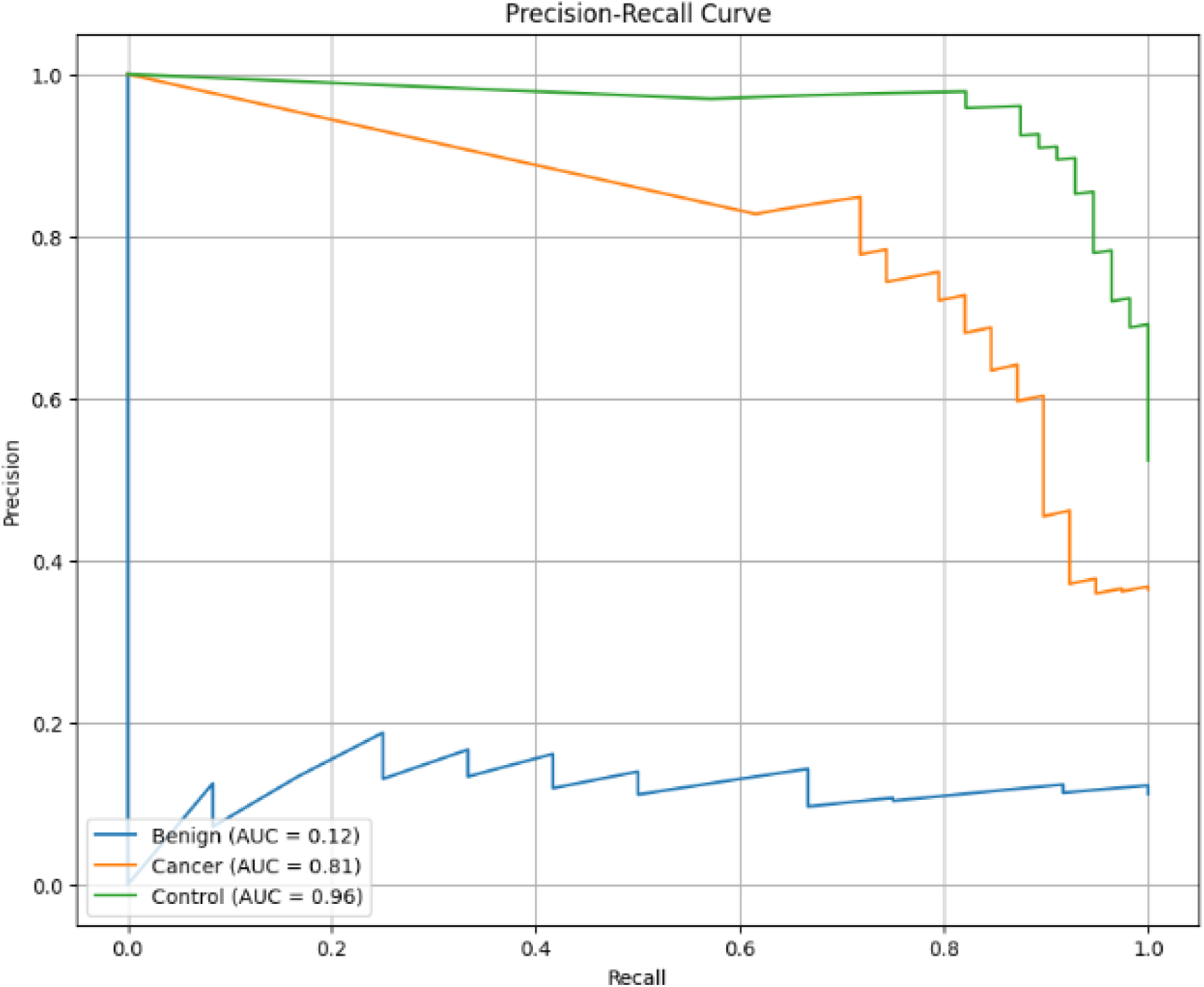
CNN PR Curve (75/25).

**Figure 7.13.**
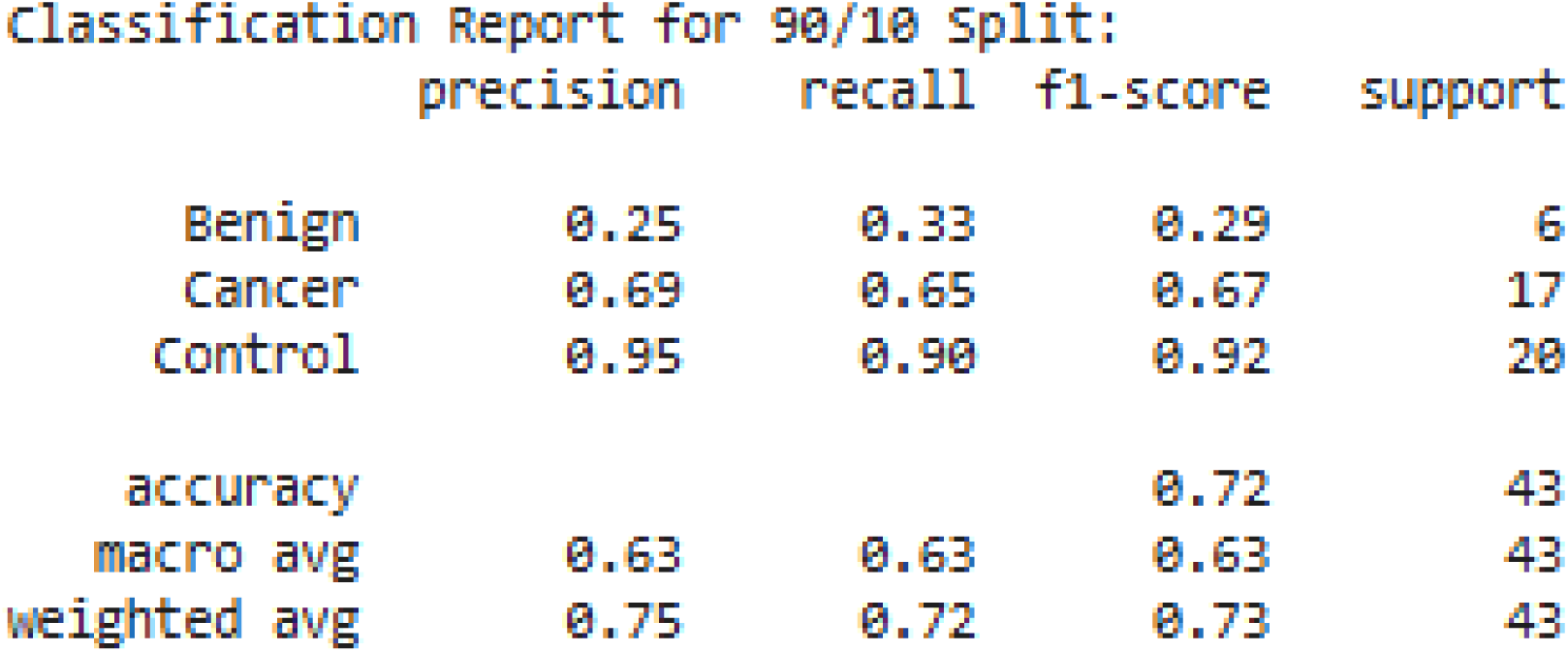
CNN Classification Report (90/10).

**Figure 7.14.**
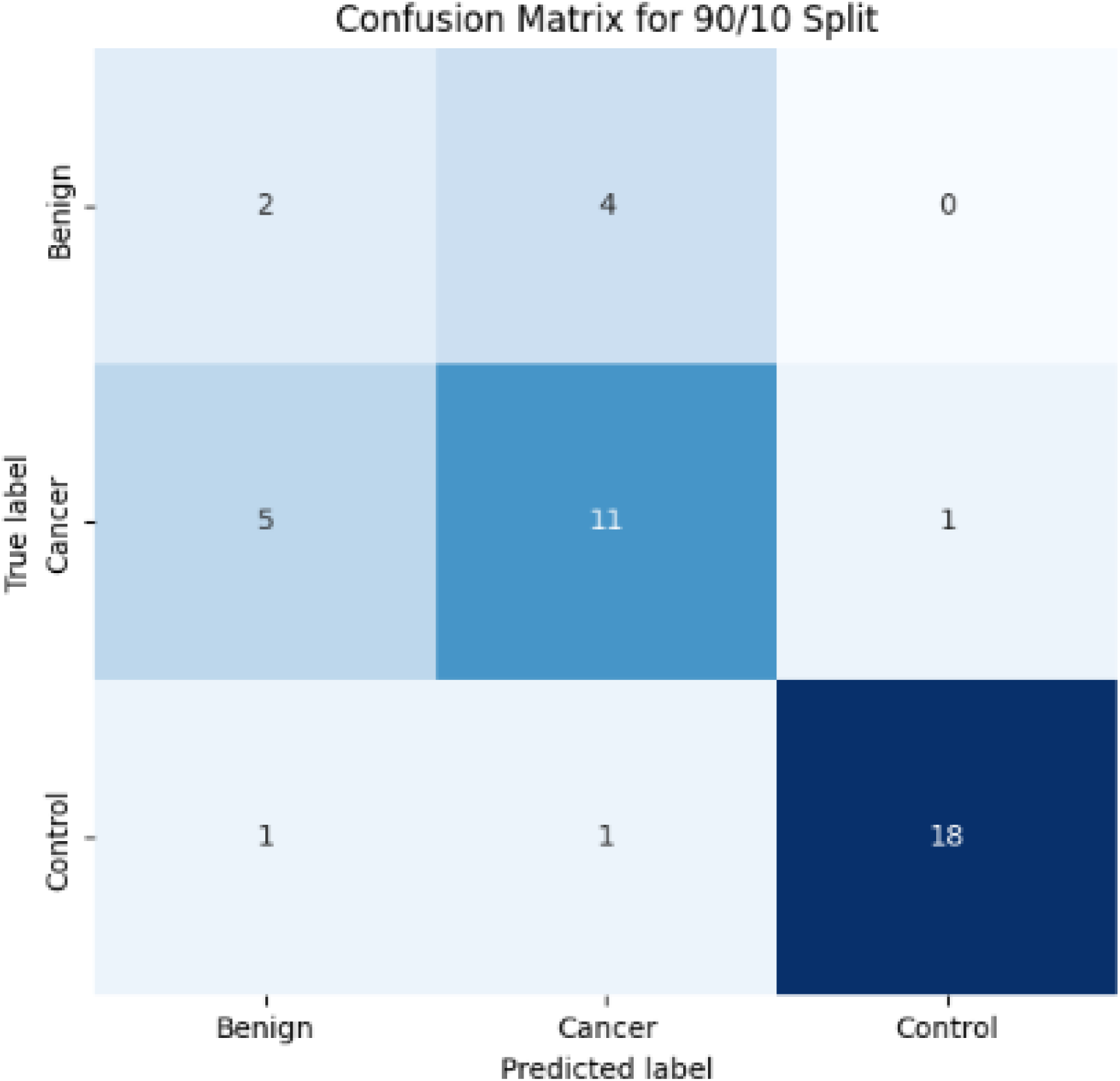
CNN Confusion Matrix (90/10).

**Figure 7.15.**
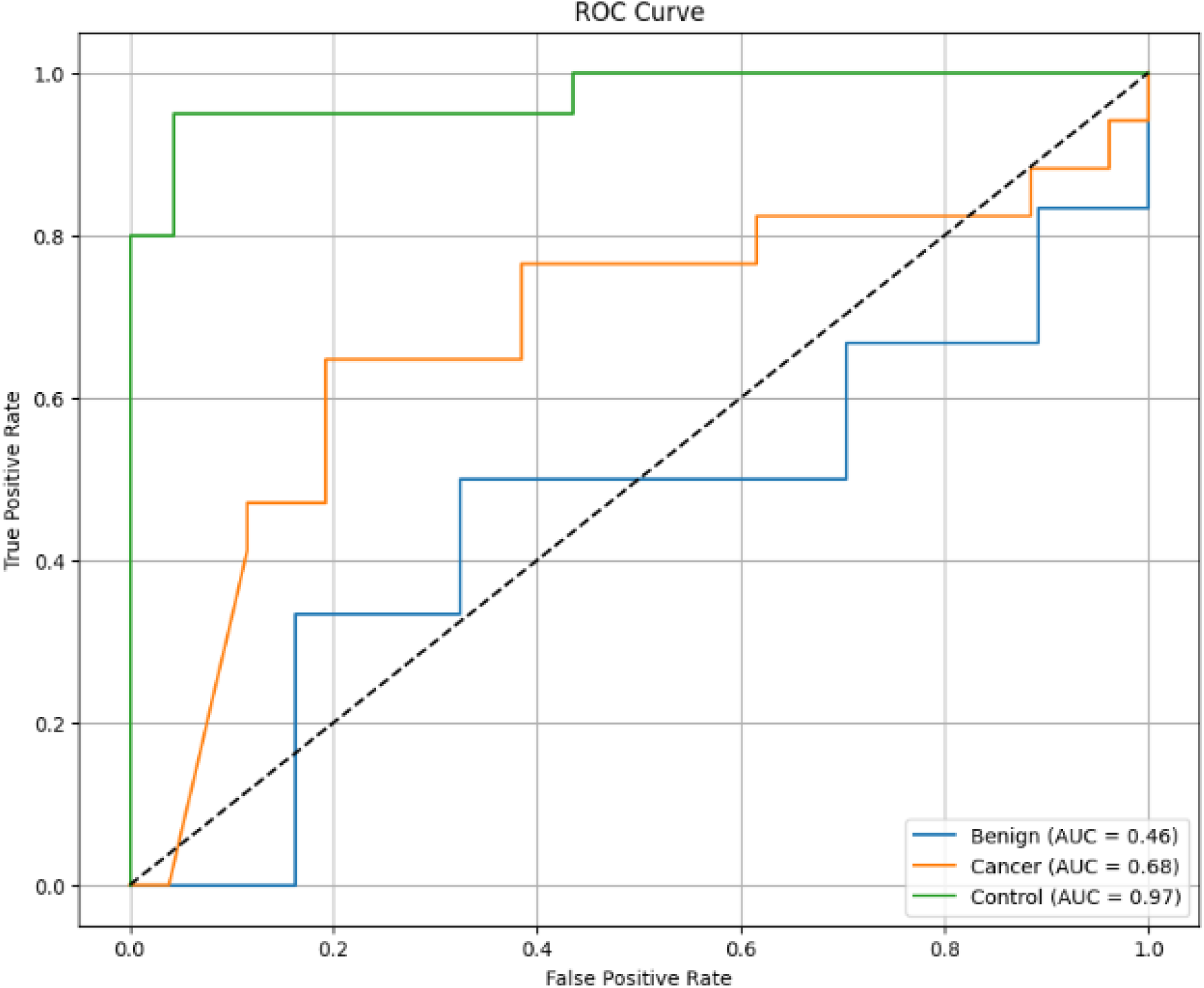
CNN ROC Curve (90/10).

**Figure 7.16.**
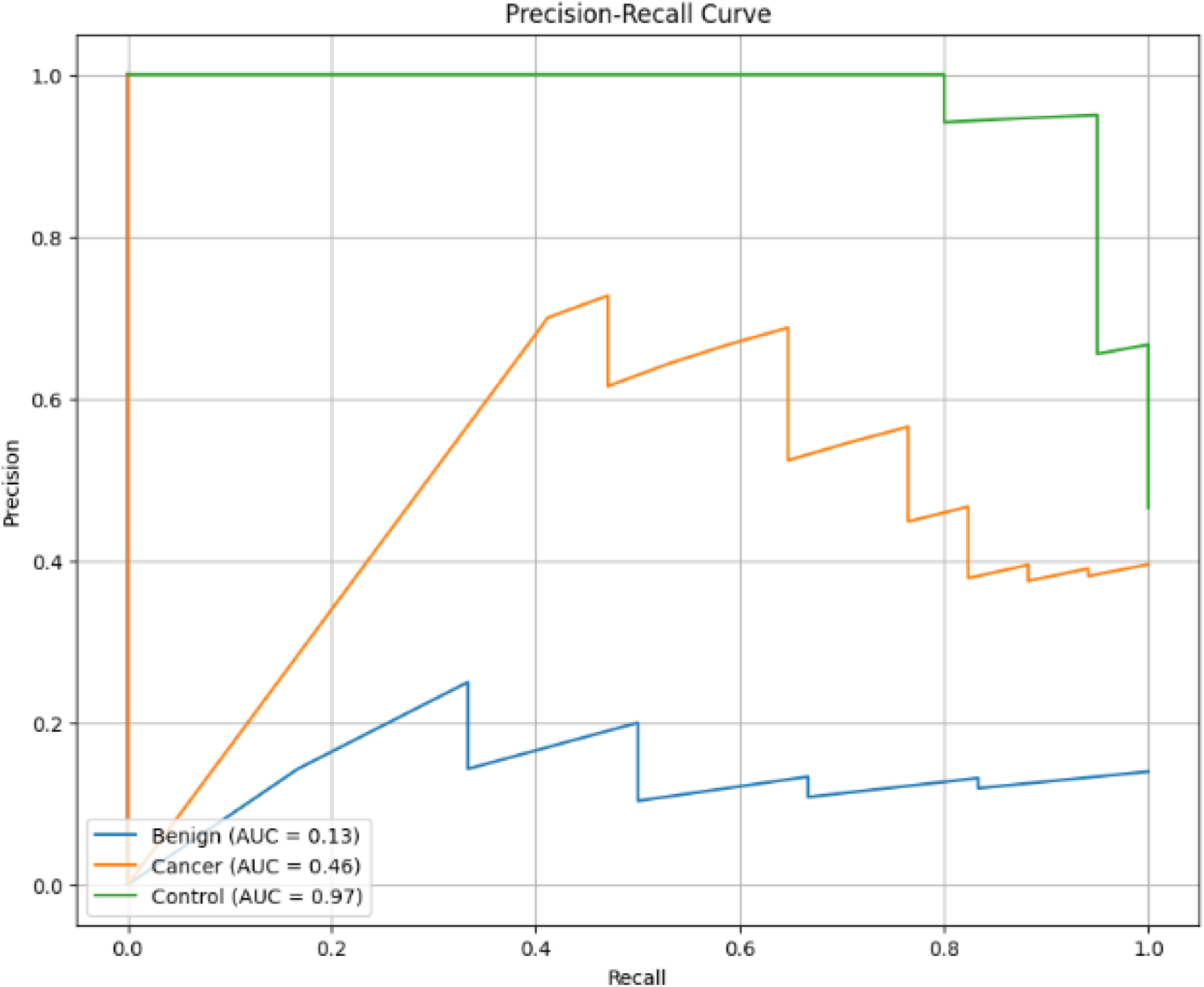
CNN PR Curve (90/10).

**8.1.**
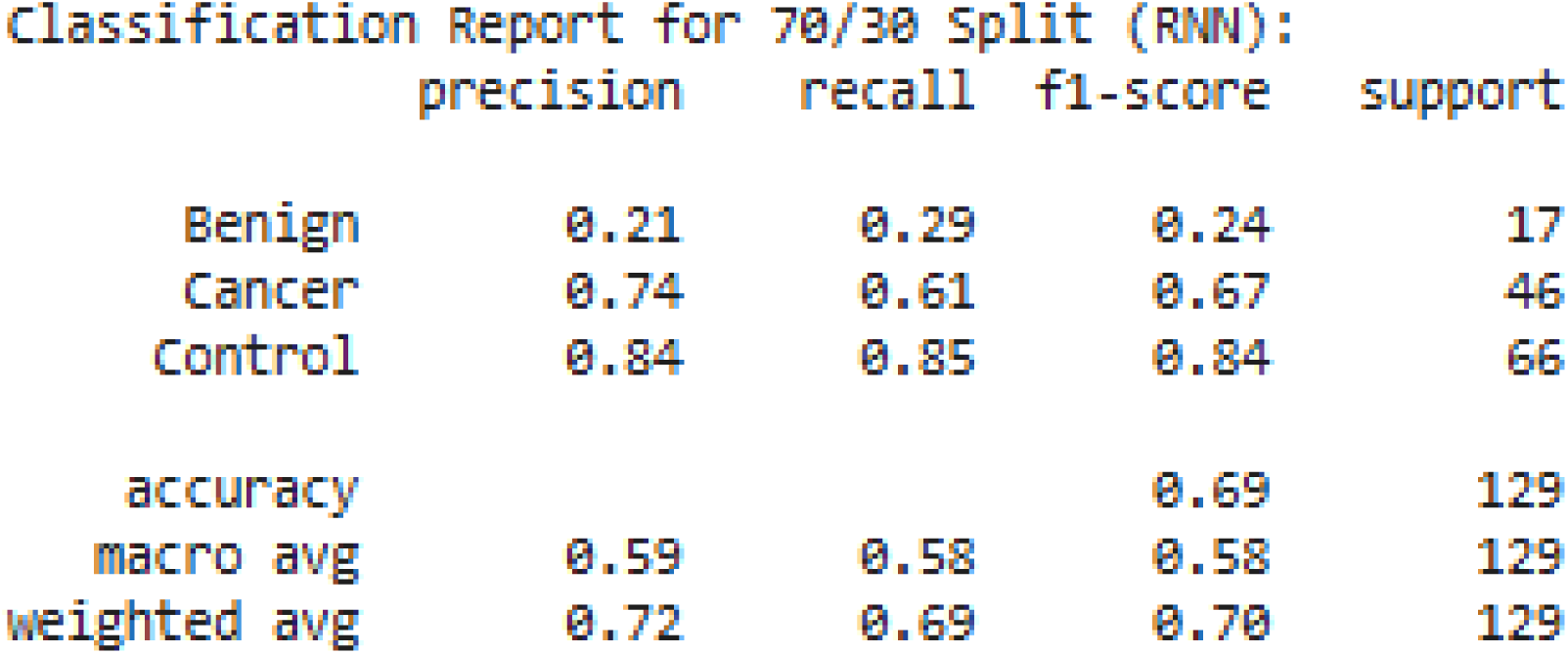
RNN Classification Matrix (70/30).

**8.2.**
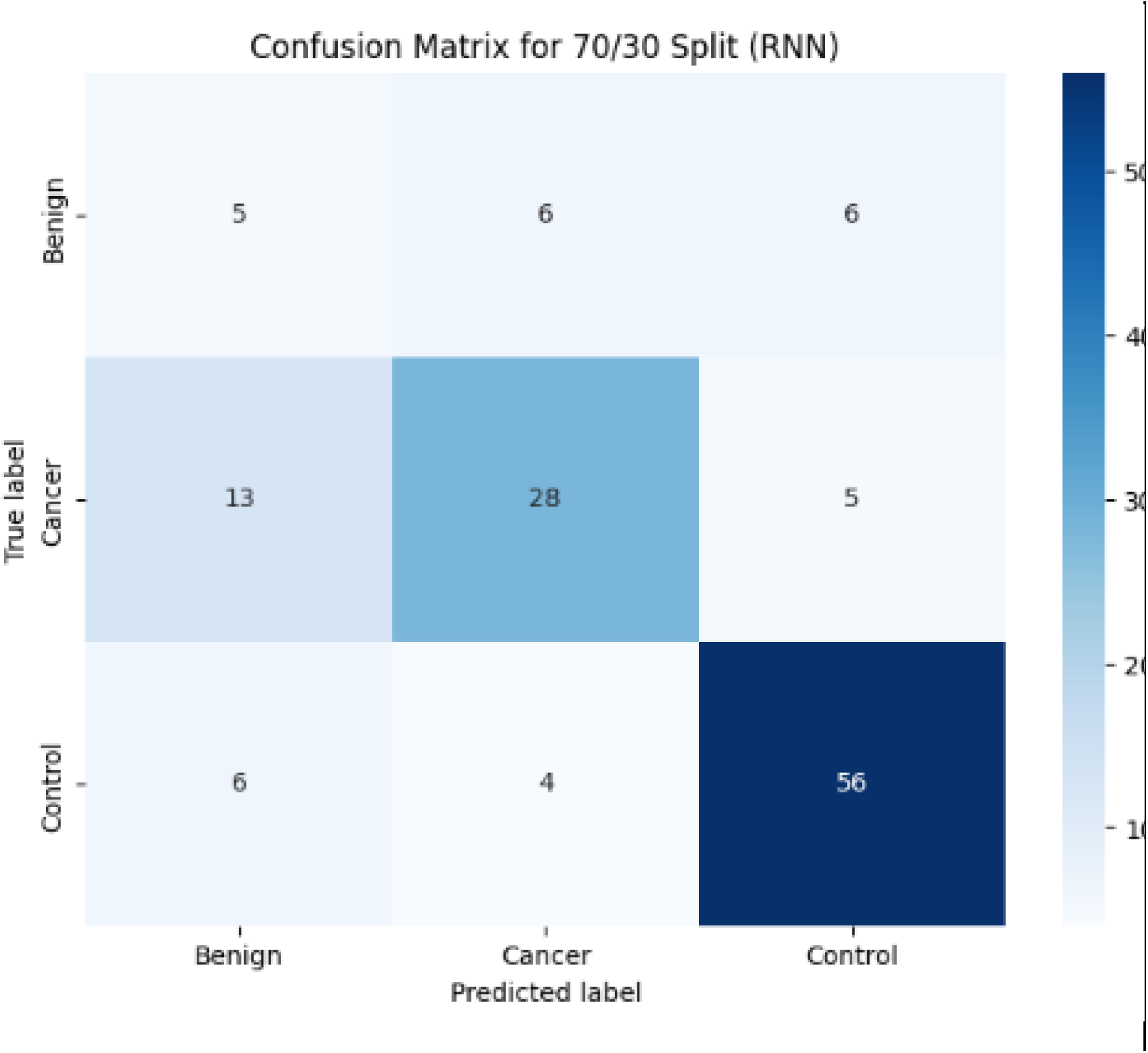
RNN Confusion Matrix (70/30).

**8.3.**
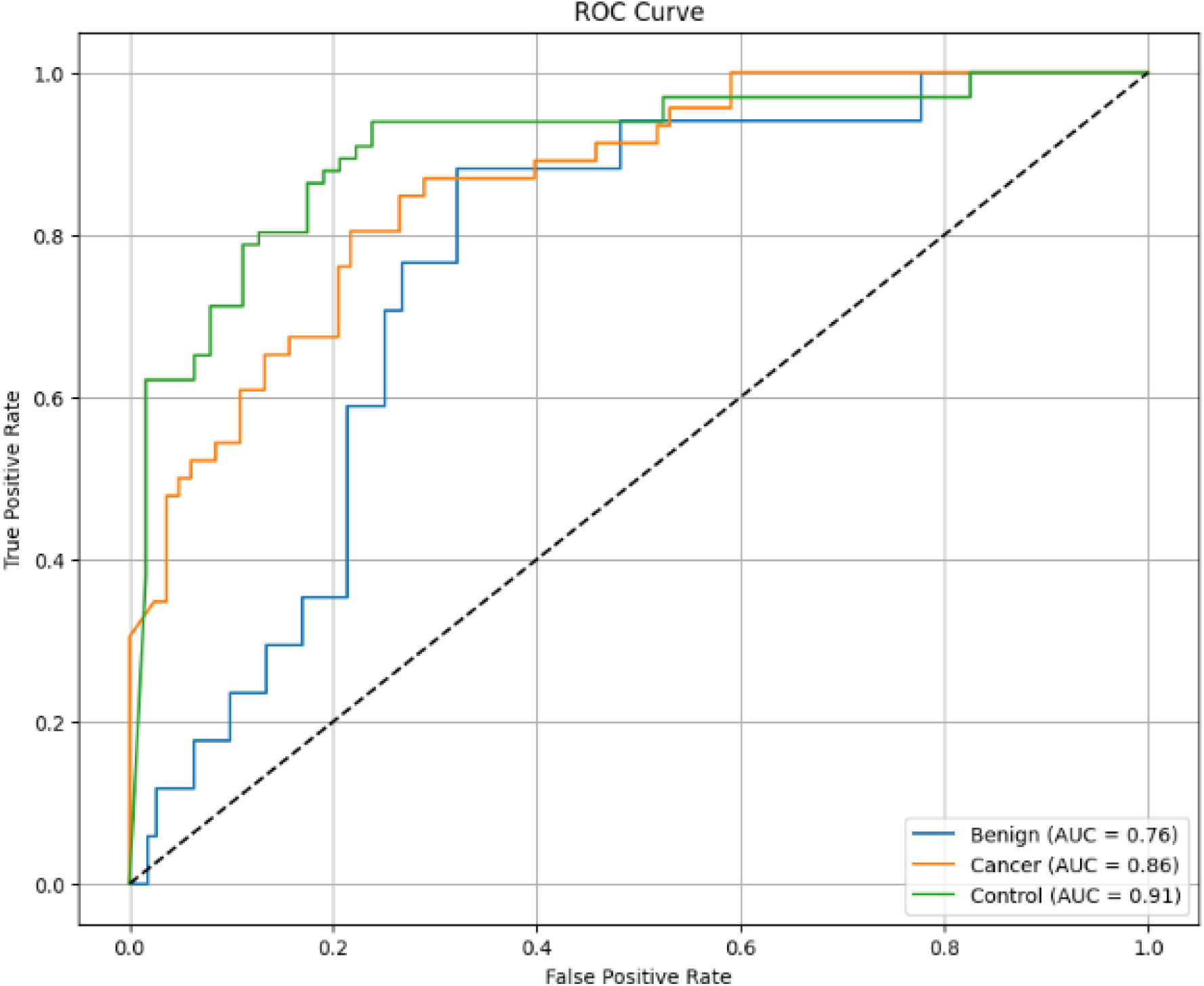
RNN ROC Curve (70/30).

**8.4.**
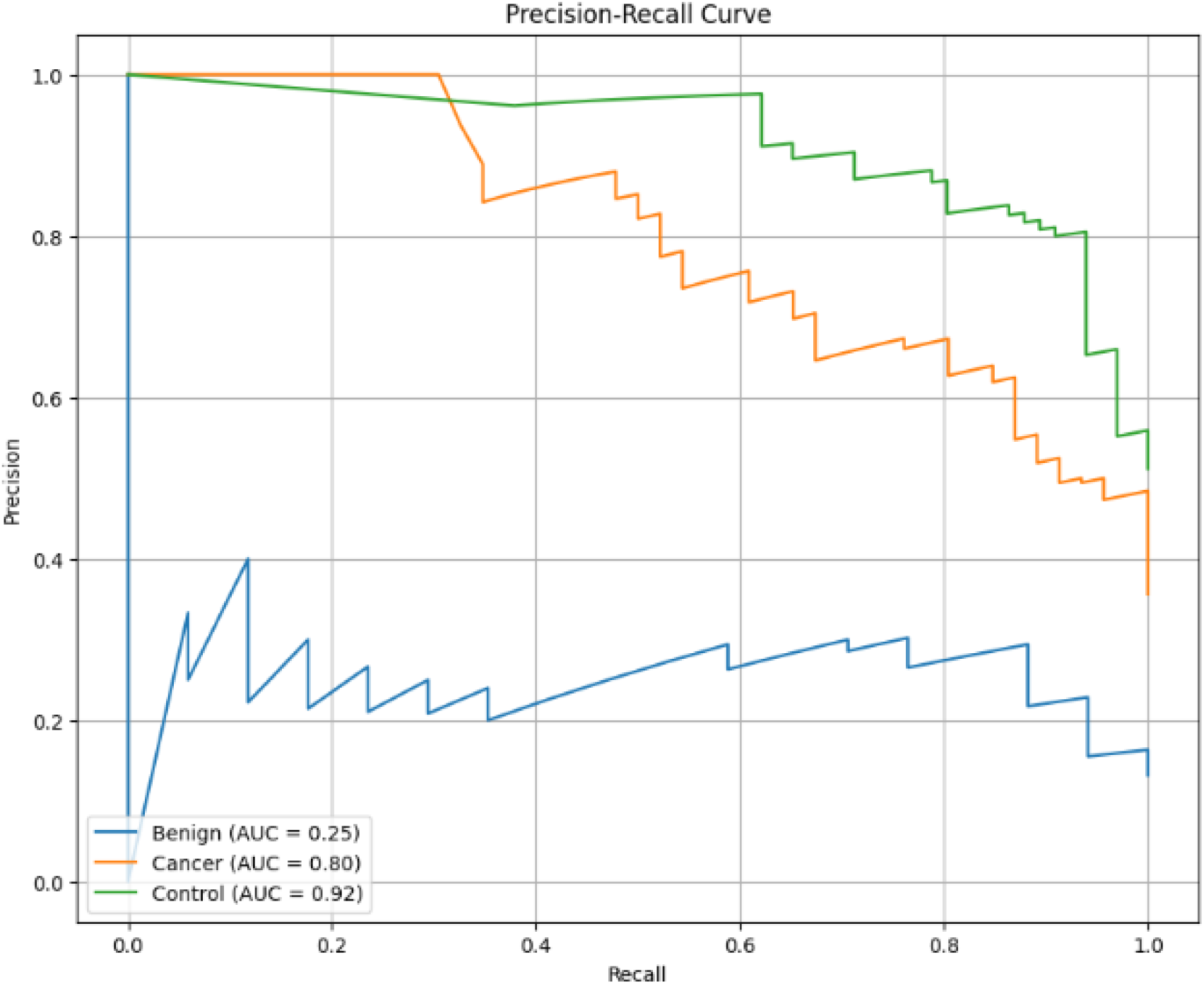
RNN PR Curve (70/30).

**8.5.**
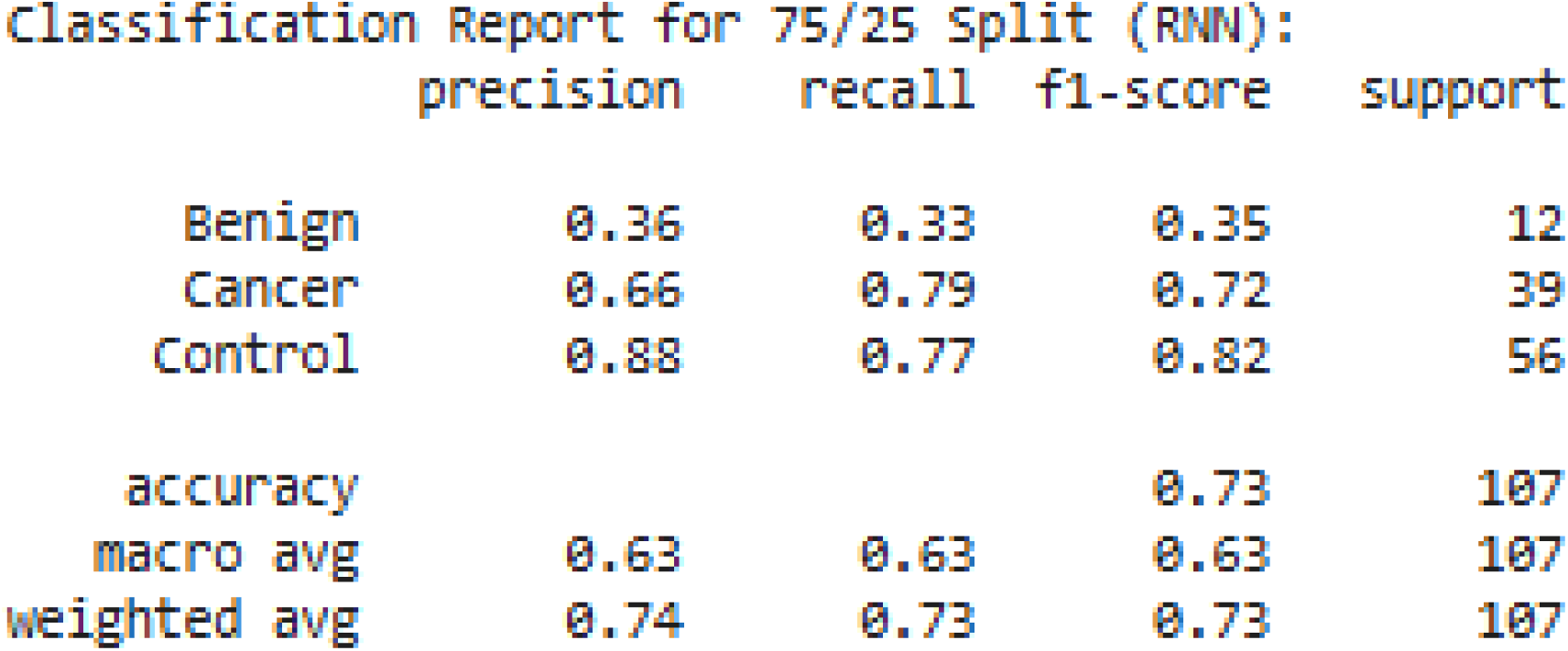
RNN Classification Report (75/25).

**8.6.**
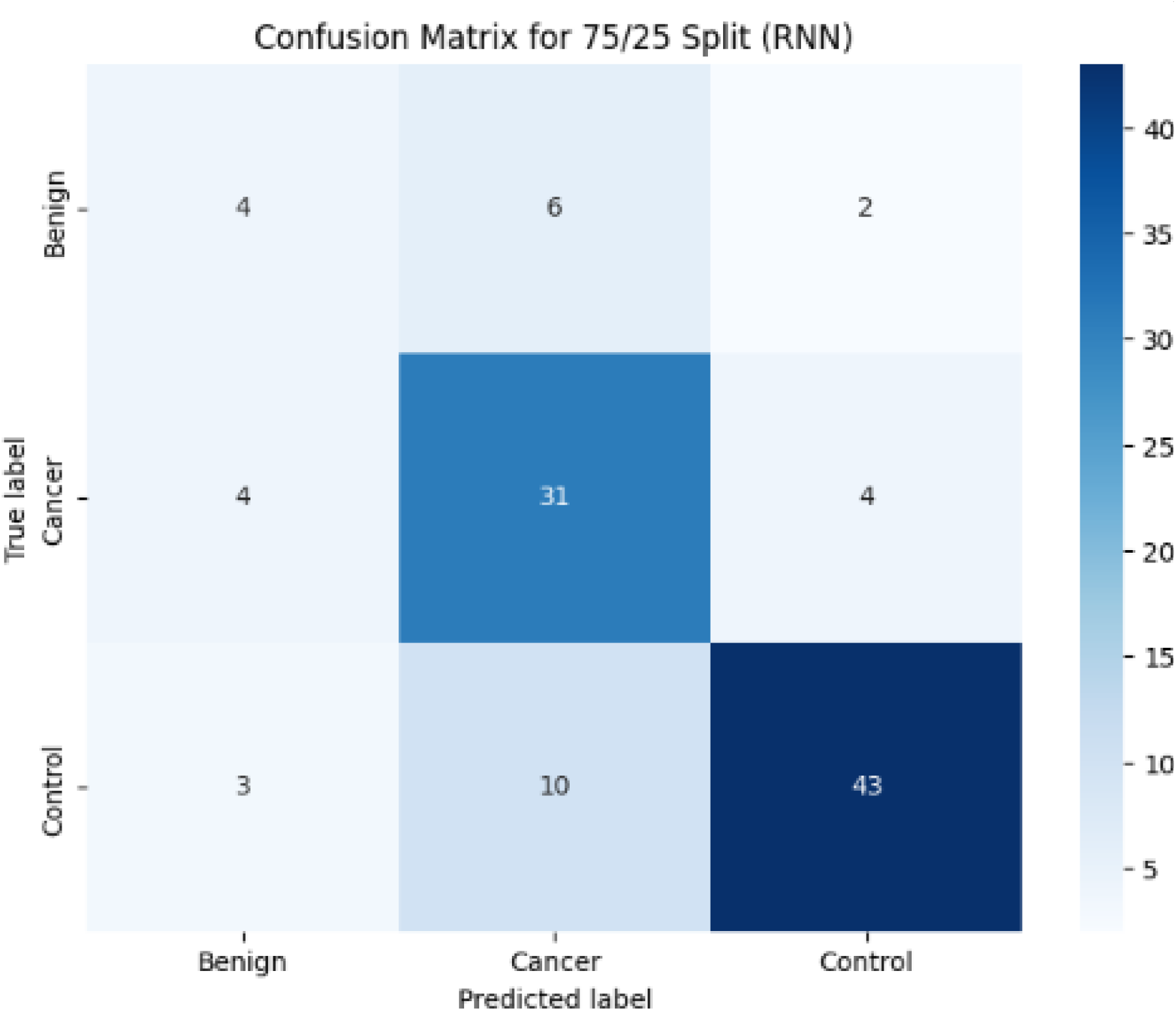
RNN Confusion Matrix (75/25).

**8.7.**
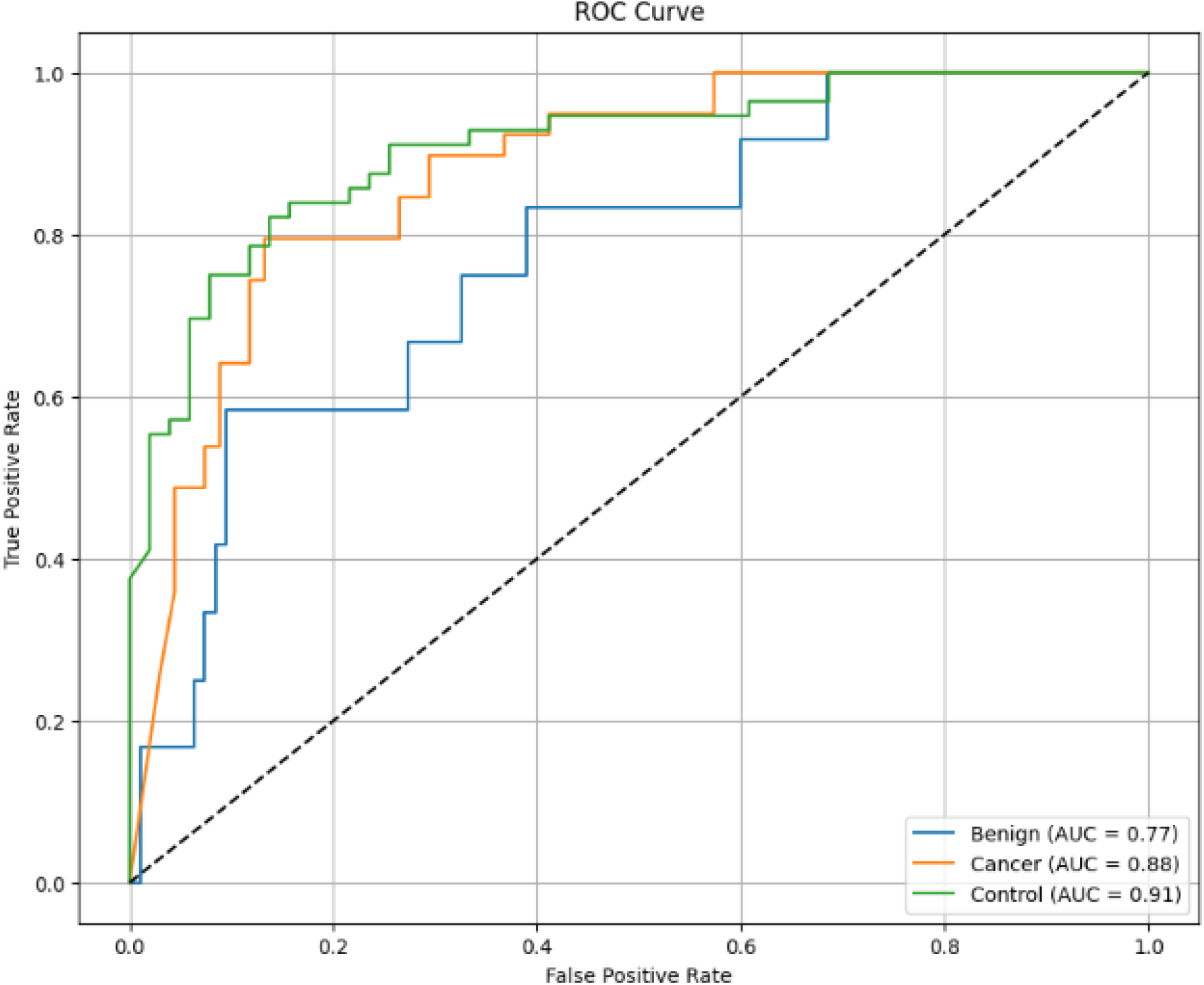
RNN ROC Curve (75/25).

**8.8.**
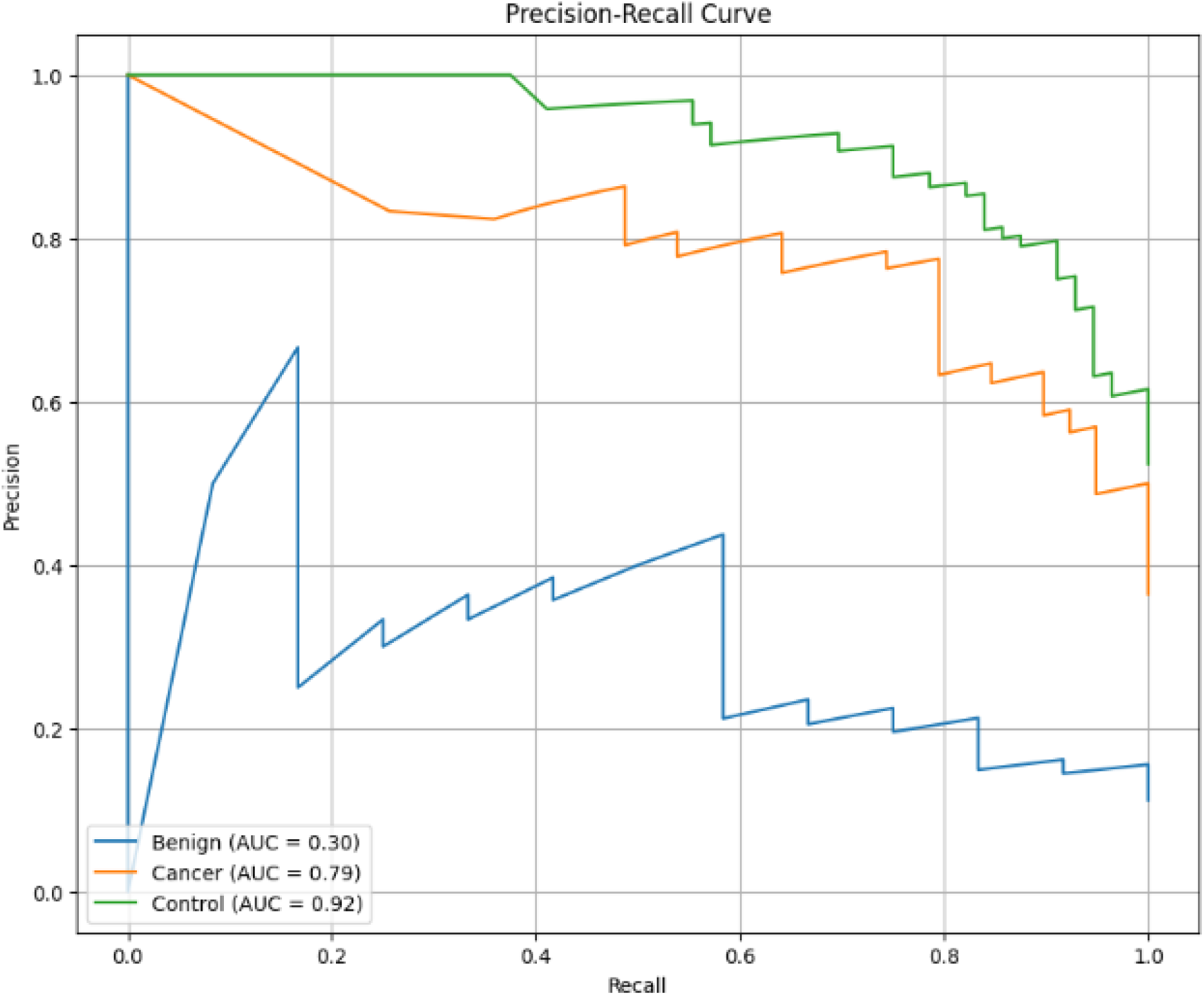
RNN PR Curve (75/25).

**8.9.**
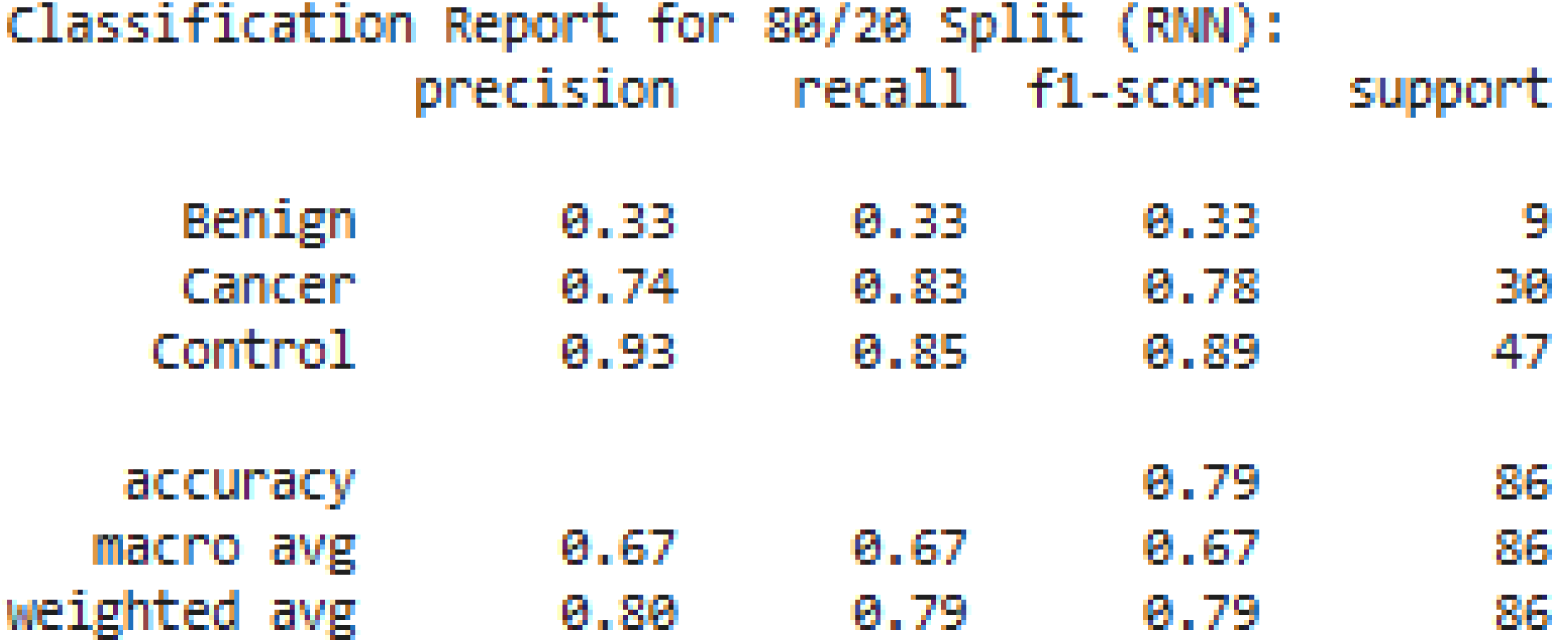
RNN Classification Report (80/20).

**8.10.**
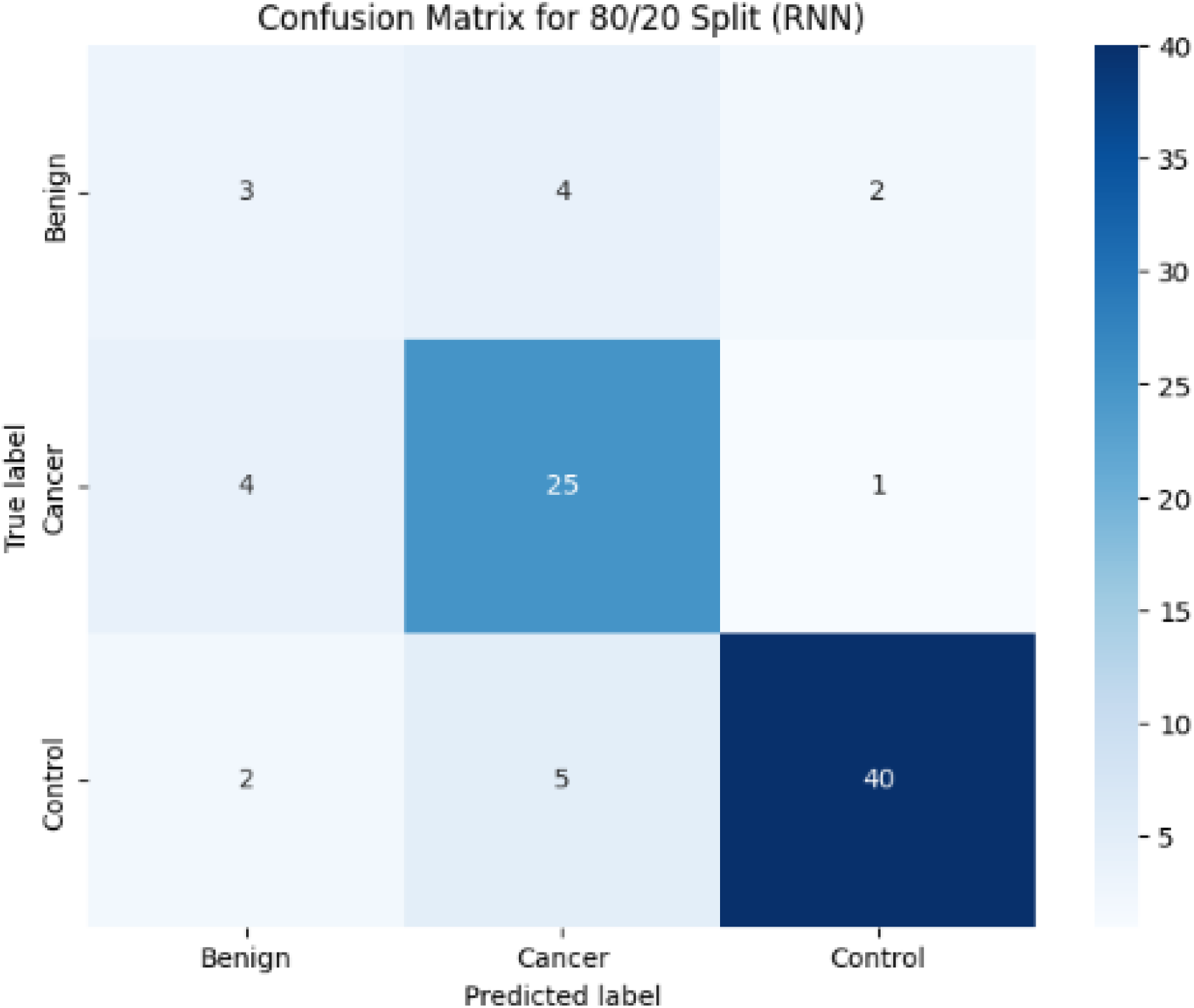
RNN Confusion Matrix (80/20).

**8.11.**
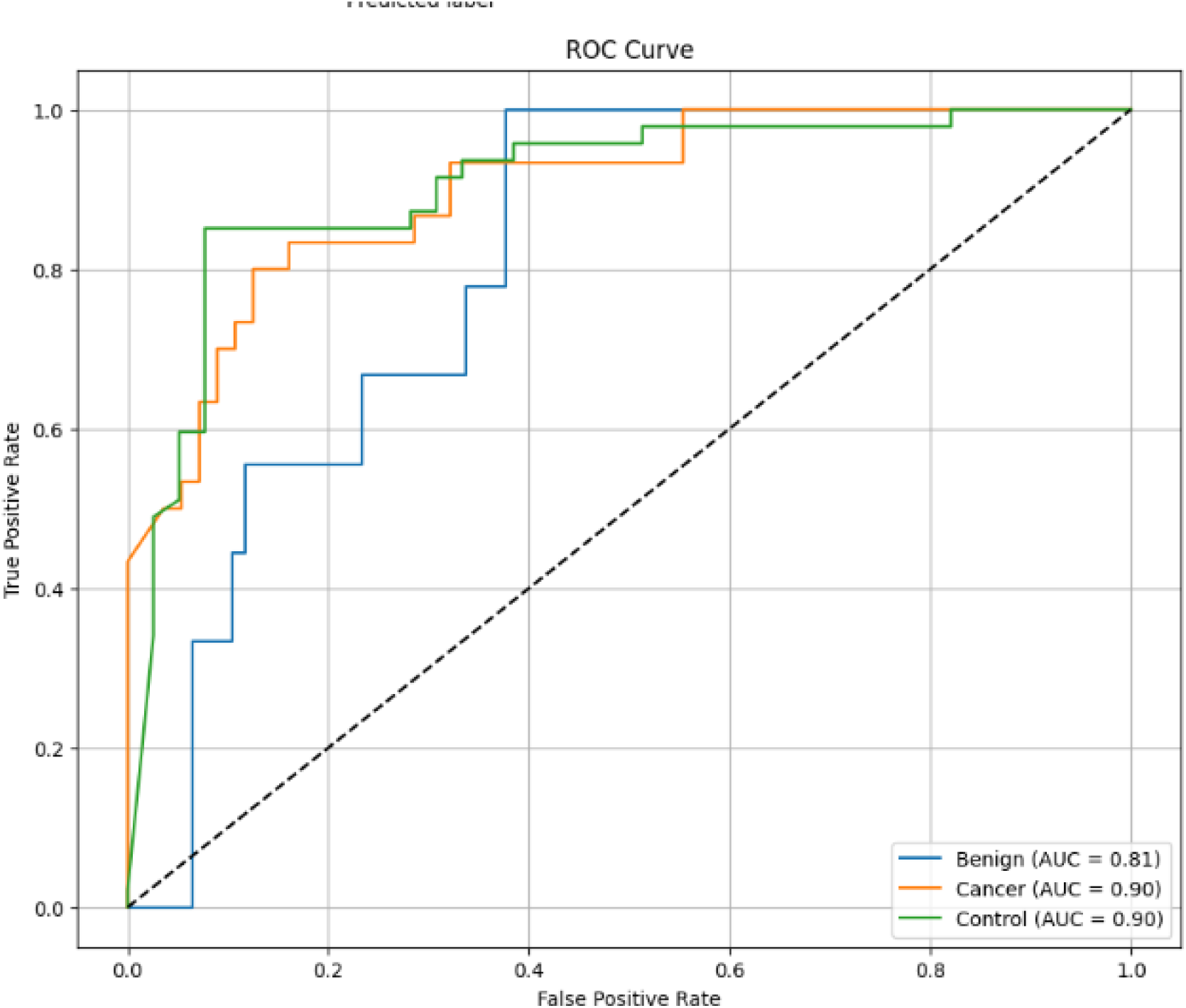
RNN ROC Curve (80/20).

**8.12.**
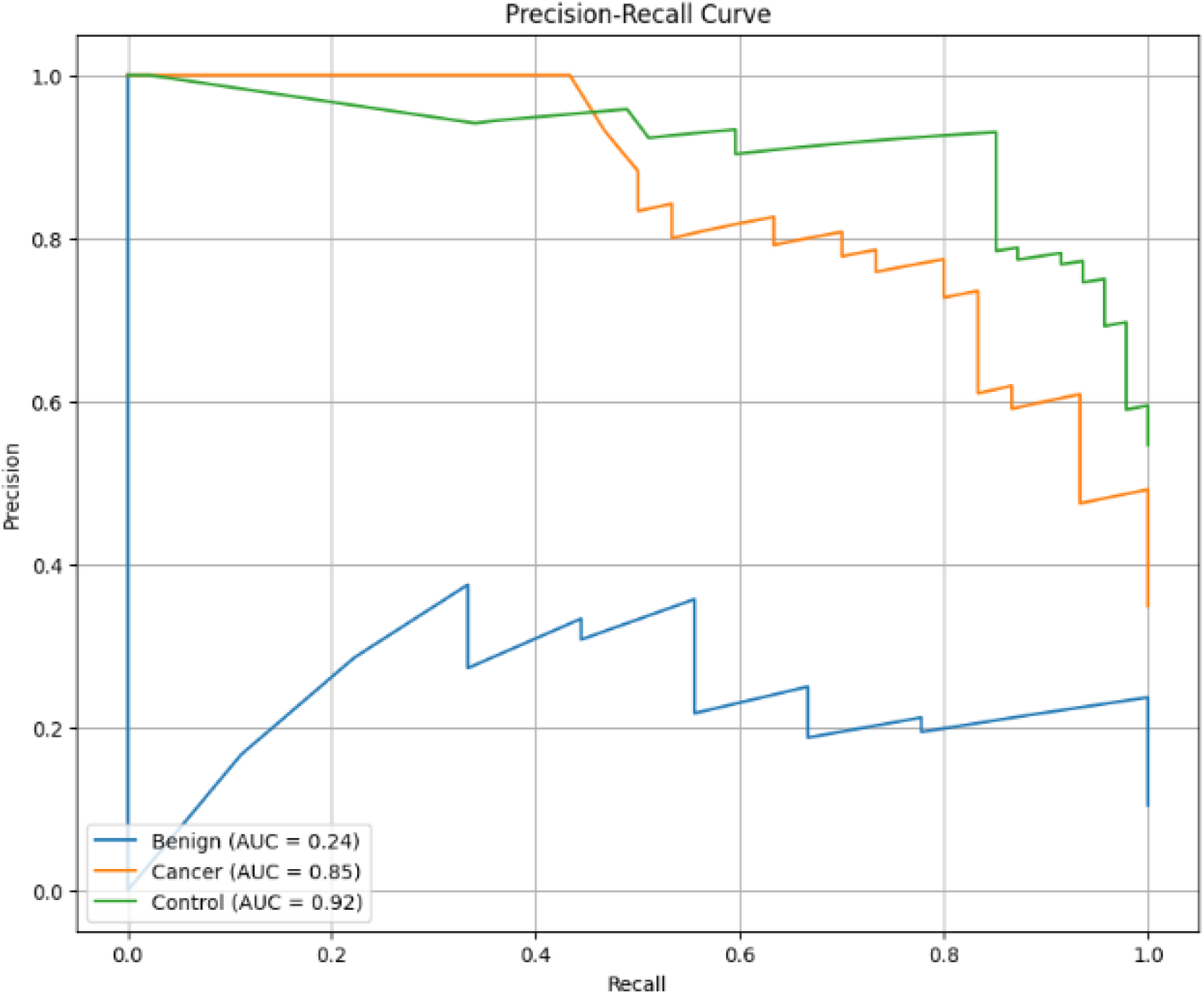
RNN PR Curve (80/20).

**8.13.**
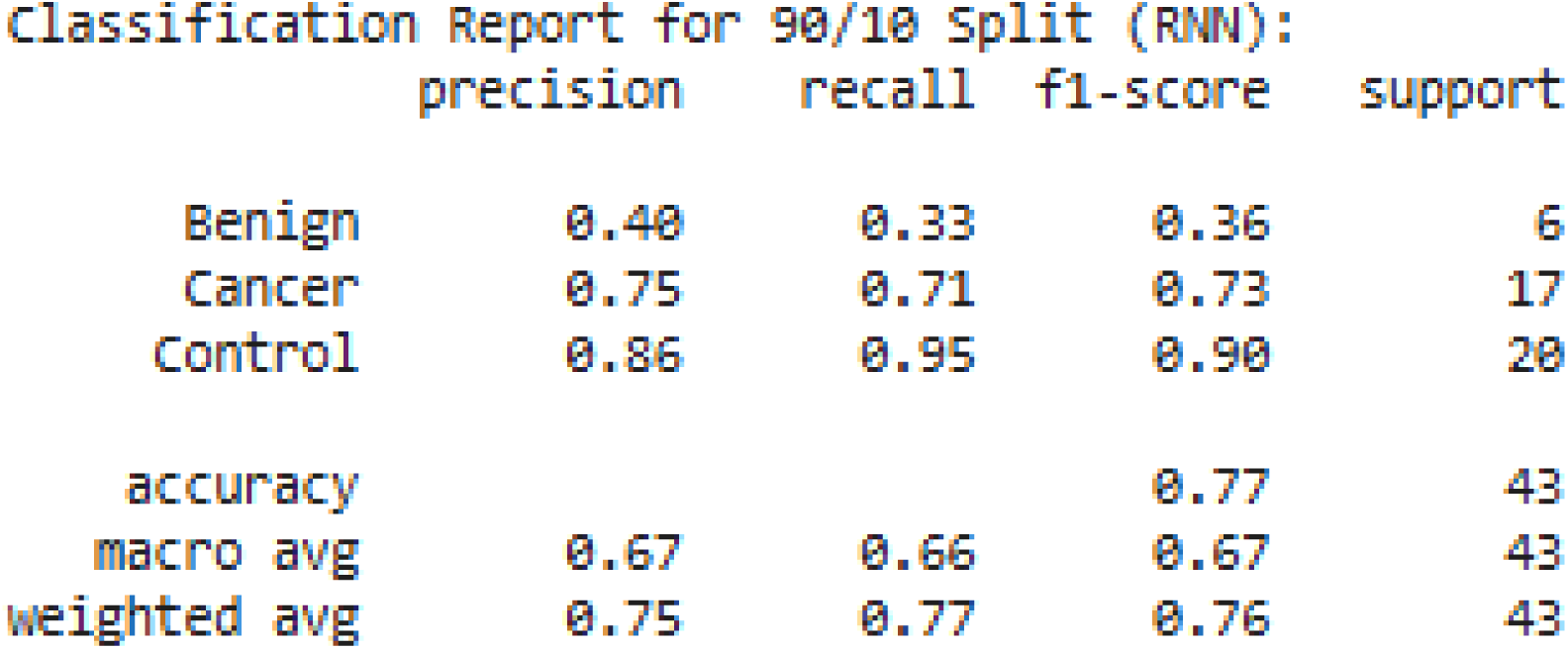
RNN Classification Report (90/10).

**8.14.**
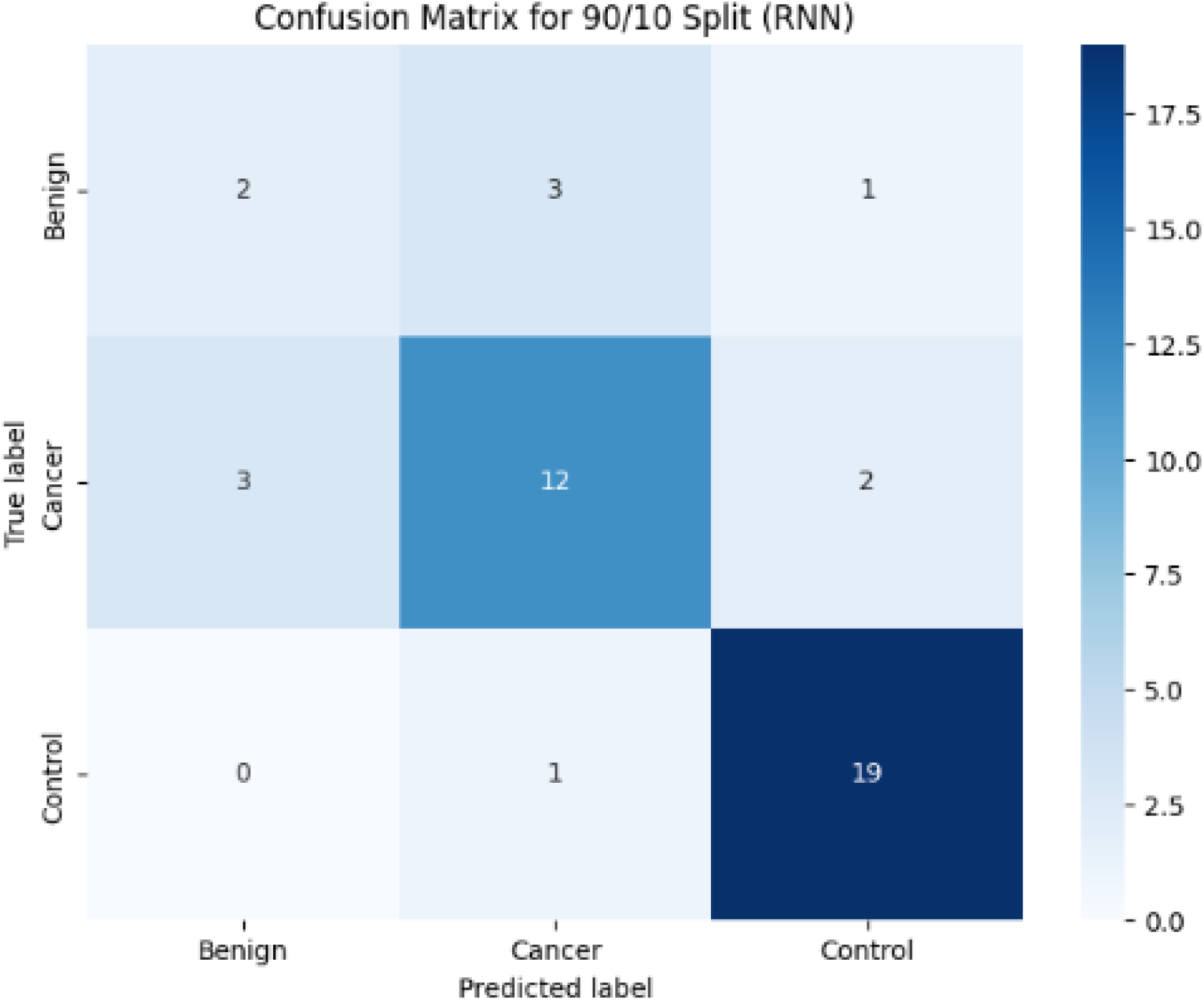
RNN Confusion Matrix (90/10).

**8.15.**
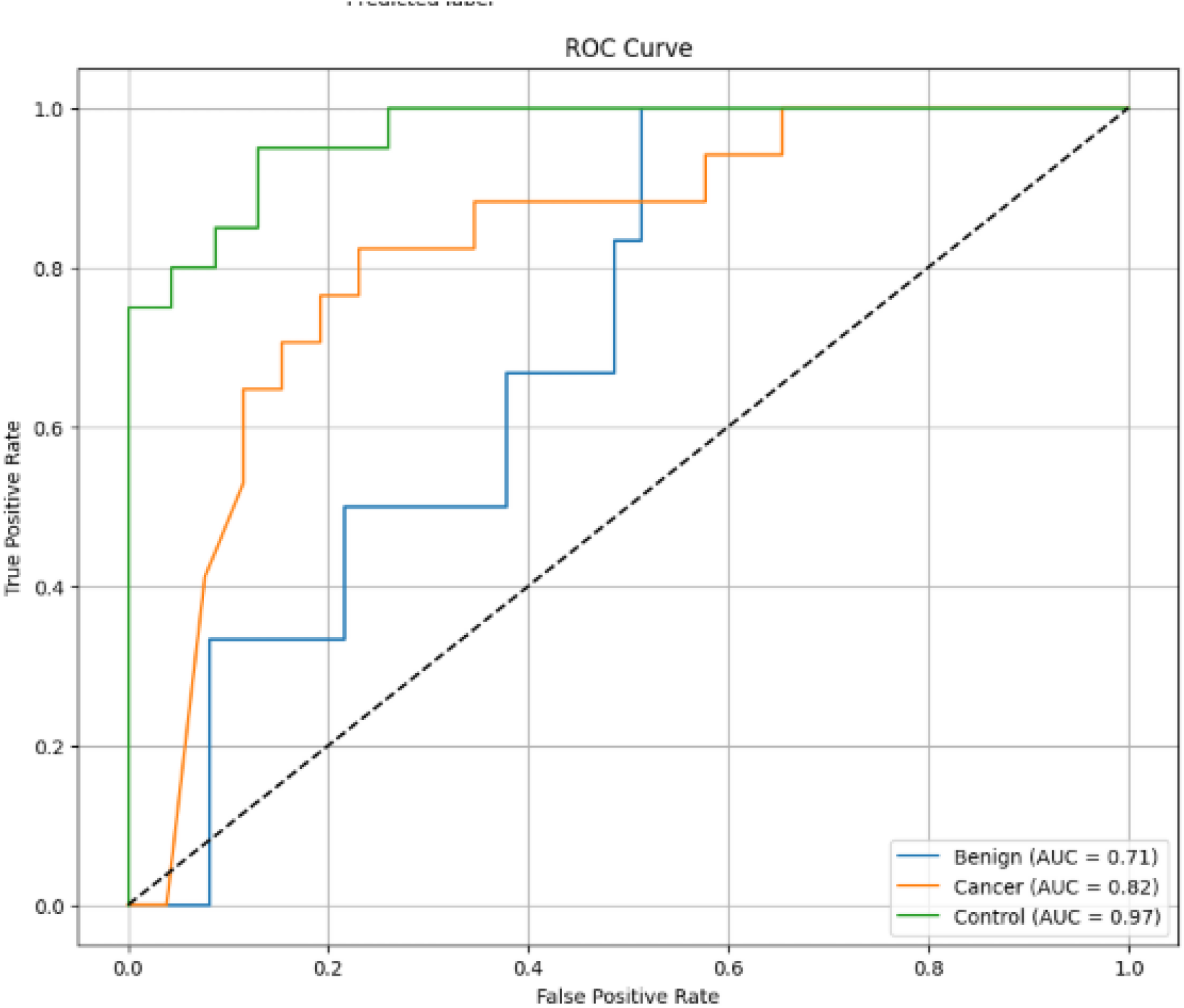
RNN ROC Curve (90/10).

**8.16.**
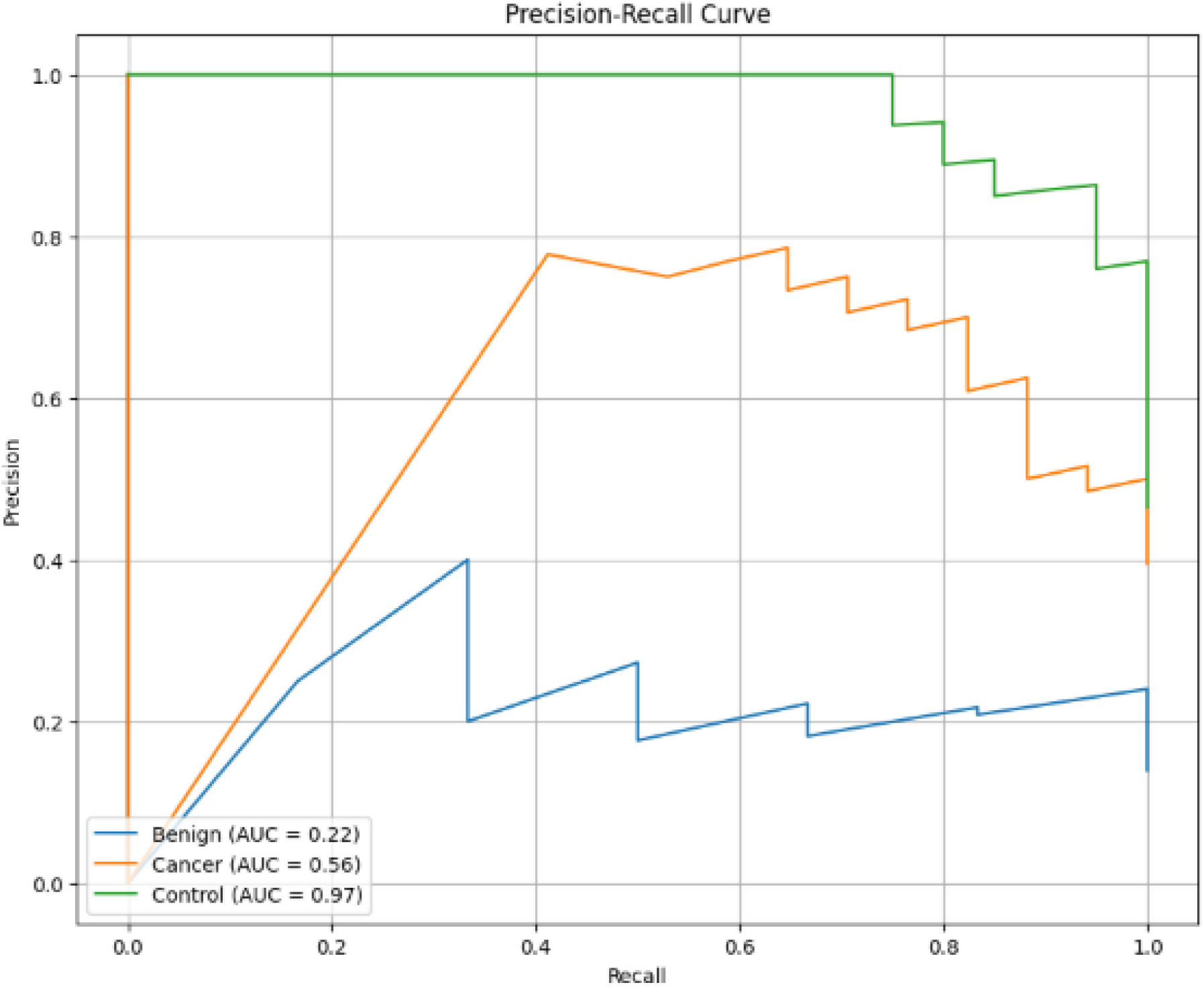
RNN PR Curve (90/10).

